# Healthcare presentations with self-harm and the association with COVID-19: an e-cohort whole-population-based study using individual-level linked routine electronic health records in Wales, UK, 2016 - March 2021

**DOI:** 10.1101/2021.08.13.21261861

**Authors:** Marcos DelPozo-Banos, Sze Chim Lee, Yasmin Friedmann, Ashley Akbari, Fatemeh Torabi, Keith Lloyd, Ronan A Lyons, Ann John

## Abstract

**Background:** Multi-setting population-based studies on healthcare service presentations with self-harm covering the first 12 months of the COVID-19 pandemic are yet to be published.

**Aims:** Ascertain changes across settings in healthcare service presentations with self-harm during Waves 1 and 2 of the COVID-19 pandemic.

**Method:** E-cohort study using individual-level linked routine healthcare data from Wales, UK, 2016-March 2021. We measured weekly proportion of self-harm contacts and people who self-harmed in contact with general practice (GP), emergency department (ED) and hospital admissions. We modelled weekly trends using linear regression and generalised estimated equations, quantifying time differences using difference-in-difference (DiD).

**Results:** We included 3,552,210 Welsh residents aged ≥10 years. Counts of self-harm presentations across settings was at a minimum at the start of stay-at-home restrictions during both waves and recovered compared to previous years in 3-5 months. Those who self-harmed in April 2020 were more likely to be seen in GP compared to other settings and previous years – mean rate of OR=1.2, although actual numbers fell. The proportion of self-harm ED contacts admitted to hospital dropped from June 2020 (1.9 [1.5-2.3] pp/month). Self-harm and COVID-19 infection had a bidirectional effect – self-harm history had OR=1.4 [1.2-1.6] and incidence had DiD=1.1 [0.8-1.4].

**Conclusions:** Those that self-harmed and sought help during the COVID-19 pandemic potentially encountered stringent criteria for hospitalisation, particularly in Wave 2, while in Wave 1 they preferentially presented to GP. Reductions in contacts likely resulted in unmet healthcare needs which may later emerge placing further burden on individuals and healthcare services.

**Relevance statement:** This study provides novel findings on how the COVID-19 pandemic and the measures taken to curb its spread affected self-harm healthcare service presentations. To our knowledge no other population-based studies in the UK have linked routinely collected general practice (GP), emergency department (ED) and hospital admission data covering Waves 1 and 2 of the pandemic.

Reductions in presentations with self-harm during the pandemic may be the result of those not requiring ED care or hospitalisation avoiding seeking help during the pandemic as often as before. Those that did seek help potentially encountered more stringent criteria for hospitalisation, particularly during Wave 2. This likely resulted in unmet healthcare needs which may later emerge placing further burden on individuals and healthcare services. Measures should be put in place to ensure that those who self-harm receive appropriate assessment and intervention.

## Introduction

Concerns were raised early on in the COVID-19 pandemic that the consequences of the infection and the measures taken to curb its spread were likely to have a considerable impact on people’s mental health and suicidal behaviours.^1^ Most studies comparing service utilisation in the first months of the pandemic (March-August 2020) with similar periods in previous years reported a reduction in the number of presentations to emergency departments (EDs), psychiatric emergency services and trauma centres with suicidal thoughts, behaviours and self-harm compared to previous years.^2^ A 30% fall (compared to expected values) in consultations for self-harm in April to June 2020 in primary and secondary care in the UK has been reported.^3^ A small number of studies showed an increase in self-harm / suicide attempts.^4,5^ There was some evidence of a return to pre-lockdown levels by May 2020.^6^ The reduction in service utilisation for self-harm and suicidal behaviours was attributed to fear of catching the virus in healthcare settings, avoidance of being a burden to overstretched healthcare services, the conversion of mental health facilities to treat COVID-19 patients, and remote delivery of services.^7^ Some suggested COVID-19 infection was positively linked with self-harm and suicidal behaviours.^8,9^

Many of these studies were based on single healthcare settings and/or specific sites and sub-populations, hindering cross-setting analyses and the generalisation of findings. There is also scant evidence of service utilisation for self-harm and suicidal behaviours during Wave 2 of the pandemic into early 2021. In this study we aimed to quantify self-harm contacts across the population of Wales, UK, by age, sex, and deprivation status across healthcare settings (primary care, EDs and hospitals) during Waves 1 and 2 of the COVID-19 pandemic. We compare before and during COVID-19 periods from 1 January 2020 to 14 March 2021 with similar periods in counterfactual years 2016-2019. We also present preliminarily analyses on the association between history of self-harm service presentation, COVID-19 infection and changes in self-harm presentation following COVID-19 infection, similar to that previously conducted in psychiatric conditions.^10^

## Methods

### Design

An e-cohort study using individual-level linked routine electronic healthcare records in Wales, UK, 2016-2021.

### Data sources

Individual-level linkable data sources were accessed within the Secure Anonymised Information Linkage (SAIL) Databank. SAIL is a privacy-protecting trusted research environment which holds anonymised population-scale data, with an expanding repository of routinely collected healthcare, social and administrative data pertaining to the population of Wales, approximately 3.5 million individuals.^11,12^ SAIL is powered by the ISO 27001 certified UK Secure e-Research Platform, developed at Swansea University.

Ethical approval was granted from SAIL’s independent Information Governance Review Panel (project 0911), under SAIL Databank permissions for the analysis of anonymised linked data.^13^ Results requested out of the SAIL gateway were reviewed independently to ensure compliance with information governance policies.

We used all data linked deterministically or probabilistically with a linkage score ≥ 0.9 from datasets: Welsh Demographic Service Dataset, Office for National Statistics mortality register, Consolidated Deaths Data Source, Wales Longitudinal General Practice (WLGP – covering 80%, i.e. 330/412, of all general practices in Wales), Emergency Department Data Set, Patient Episode Database for Wales (PEDW), Critical Care Dataset, Welsh Laboratory Information Systems (PATD) containing both COVID-19 polymerase chain reaction (PCR)/antigen tests (pillars 1 and 2) and serology/antibodies tests (pillar 3) results,^14^ the care homes data containing geographic information data on care homes, a list of COVID-19 shielded persons who are identified as clinically extremely vulnerable and advised to self-isolate during the pandemic,^15^ and the UK Census 2011 data. These data sources are updated frequently to support COVID-19 research. For each source, we compared data across updates to ascertain the end date of good quality data. Full details in Supplementary Table 1.

### Study population

Our study population included all people living in Wales between 1 January 2016 and 14 March 2021, followed up from 1 January 2016, their 10^th^ birthday or the date they registered with a GP being resident at a Welsh address (whichever was latest), until the 14 March 2021, death or de-registering from a Welsh address (whichever was earliest). Intermittent Welsh residency was allowed during the follow-up period. For weekly measures, the sub-population meeting the criteria on each Monday was considered.

For the analysis of self-harm and COVID-19 infection, we used the sub-population meeting the inclusion criteria on 28 February 2020, the date of the first reported COVID-19 case in Wales (Suppl. Figure 1).^16^ Identification of COVID-19 infection was sourced from WLGP, PEDW and PATD by using Read codes, ICD-10 codes and positive PCR test results. COVID-19 infection was classified in two dimensions: 1) whether an individual was currently or previously infected (history) and 2) whether the infection was laboratory confirmed or suspected/probable.^17,18^

### Measures

We measured variables weekly as per the ISO week date standard (ISO-8601; www.iso.org/iso-8601-date-and-time-format.html) from 4 January 2016 (week 1 of 2016) to 14 March 2021 (week 10 of 2021). We defined Wave 1 of the pandemic from 9 March 2020 to 16 August 2020 and Wave 2 from 17 August 2020 to 14 March 2021. Demographic variables were extracted on 1 January 2016 and at the start of each week from the WDSD: sex; age categories (10-24, >24 years); and level of deprivation by geographical location extracted residential records. We identified the area level deprivation for each individual using the Lower-layer Super Output Area (LSOA) of residence version 2011 (containing approximately 1500 individuals each and the Welsh Index of Multiple Deprivation (WIMD) 2014 score.^19^ WIMD deprivation levels 1 to 5 were defined using national WIMD score quintiles as cut-offs, with level 1 representing the lowest deprivation areas.

We examined all contacts with GP, ED and hospital admissions independently. We defined ‘contact’ as a recorded entry in one of the data sources. In WLGP, we excluded administrative codes and associated diagnoses such as ‘letter from ED’ but included telephone and face-to-face contacts with any member of the primary care team.^20^ From these, we identified self-harm contacts using validated code lists.^21^ For hospital admissions, we collapsed hospital transfers into a single spell (continuous period as an inpatient), propagated diagnoses across episodes within a spell and measured hospital spell admissions and discharges.

Linking across data sources, we measured contacts across settings, ED presentations followed by hospitalisation and hospital admissions with transfers to critical care. When measuring contacts across settings, admissions from ED to hospital were collapsed into a single contact to avoid double counting episodes, but diagnoses were not propagated across settings. Thus, the number of contacts with GP, ED and hospital admissions separately do not necessarily add up to the number of contacts across all settings.

For each week, we counted the number and proportion of self-harm to all contacts across settings and in each specific setting. We also measured the number of individuals with self-harm contacts and the proportion of them presenting to one or more settings with self-harm. This included three overlapping sets (seen in GP, seen in ED, and seen in hospital admissions) and seven non-overlapping sets (seen in GP only; seen in ED only; seen in hospital admissions only; seen in GP and ED only; seen in GP and hospital admissions only; seen in ED and hospital admission only; and seen in GP, ED and hospital admission).

We identified COVID-19 infection based on results from PATD (PCR tests) and diagnostic codes extracted from the WLGP and PEDW (see Supplementary Methods for details) between 28 February 2020 and 27 August 2020, allowing 6 months to identify COVID-19 infection and 6-months to follow-up self-harm. We identified COVID-19 infection by any active, confirmed or suspected infection during case ascertainment period (Suppl. Figure 1), allowing for more accurate timing of defining index dates.^10^

### Statistical analysis

We used SQL DB2 (www.ibm.com/analytics/db2) to interrogate data within the SAIL Databank and to calculate counts, proportions, incidence and prevalence. We used Python (www.python.org/) to represent the results. All statistical analyses were performed using R (www.r-project.org) and Stata version 16.1 (StataCorp. 2019). The level of statistical significance was set at p = 0.05. Detailed model specifications, sensitivity analyses and robustness checks for all the statistical analysis below are described in Suppl. Methods.

#### Self-harm weekly contacts

We compared changes in healthcare service utilisation between before (weeks 1 to 10 of 2020: i.e., 30 December 2019 to 8 March 2020 - start of COVID-19 Wave 1) and during COVID-19 periods (any week(s) of interest from week 11 of 2020 to week 10 of 2021: i.e., 9 March 2020 to 14 March 2021 – non-urgent NHS appointments were suspended in Wales on 13 March 2020)^22^ to equivalent periods in the counterfactual years 2016-2019. The COVID-19 period includes Waves 1 (9 March 2020 to 16 August 2020) and 2 (18 August 2020 to 14 March 2021).

We report weekly time trends for all studied metrics in 2020 and averaged across 2016-2019. Proportions were expressed as percentages, and prevalence and incidence as number of individuals per 100,000 person-week (pw) and person-week at risk (pwar) respectively. We used generalised estimating equations (GEE) to model weekly time trends of outcomes adjusting for demographic variables. We quantified differences between 2020-March 2021 and 2016-2019 trends using the difference in difference (DiD) approach^23^ to account for background fluctuations.^24^ We calculated ratio of rate ratios (RRRs) for counts and ratio of odds ratios (RORs) for proportions of 2020-March 2021 compared to each counterfactual period 2016-2019. For readability, in the text we report mean RRRs and RORs across all counterfactual years and the number of comparisons with p<0.05 (full results provided in Supplementary Tables). Long-term underlying linear trends were assessed using linear regression. Bonferroni adjustment was used to correct for multiple comparisons. We repeated these analyses stratifying by age, sex and WIMD deprivation quintile separately.

#### Self-harm and COVID-19 infection association

To study the association between COVID-19 and self-harm, we defined an index date as the first date of COVID-19 infection or a random date between 28 February 2020 and 27 August 2020 for those not infected. We assessed the association of having a history of self-harm and having a subsequent COVID-19 infection using logistic regression and adjusting for other risk factors such as age, sex and ethnicity (full list in the suppl. Methods). We assessed the risk of having a self-harm contact following a COVID-19 infection by comparing the incidence of self-harm in a pre-infection period (pre-COVID; 14 to 180 days starting from two years before the index date) and post-infection period (post-COVID; 14 to 180 days after the index date) using GEE models and computing the DiD estimator (and 95% confidence intervals, CIs) from the period (pre-COVID vs. post-COVID) by infection status (infected vs. not infected – interaction term), as described in the Suppl methods. We conducted a sensitivity analysis using a more stringent COVID-19 infection ascertainment criteria and robustness checks against the common trend assumption of the DiD estimator as previously proposed.^23^

## Results

We identified 3,552,210 individuals living in Wales at some point between 1 January 2016 and 14 March 2021. The study population included the 3,220,784 aged 10 years or older during the study period (Suppl. Figure 1), of whom 1,770,973 (49.85%) were males. On 1 January 2016, 644,527 (18.14%) were 10-24 years old and 2,379,194 (66.98%) were >24 years old.

### Self-harm weekly contacts

Between 1^st^ Jan 2016 and the 8 March 2020, we identified a weekly average of 365,297 GP contacts, 15,402 ED contacts and 16,523 hospital admissions for any reason. In each setting, from 9 March 2020 the number of weekly contacts was lower than seen in previous years – except during December 2020 and January 2021, when the number of GP contacts was higher than usual. Total number of weekly contacts were used as the denominator when calculating the proportion of self-harm contacts and they can be found in Suppl. Figure 3.

Across settings, weekly counts of self-harm contacts dropped at the start of Wave 1 (mean RRR = 0.6, p<0.05 for 3/3 counterfactual years), returned to before COVID-19 levels by July-August 2020, end of Wave 1 (mean RRR = 0.9, p<0.05 for 1/3 counterfactual years), and dropped again during Wave 2 with an amplified seasonal effect in December 2020 compared to previous years (mean RRR = 0.7, p<0.05 for 3/3 counterfactual years). The weekly proportion of self-harm of all contacts and weekly number of individuals with a self-harm contact followed a similar trend. In all cases (number and proportion of self-harm contacts and number of individuals with self-harm contacts) the drop during Wave 2 was slightly less compared to Wave 1. In both waves, the downward trend was replaced by an upward trend shortly after the start of the stay-at-home measures (23 March and 16 December 2020 respectively). Trends can be seen in Figure 1 (detailed results in Suppl. Table 2).

**Figure 1.**
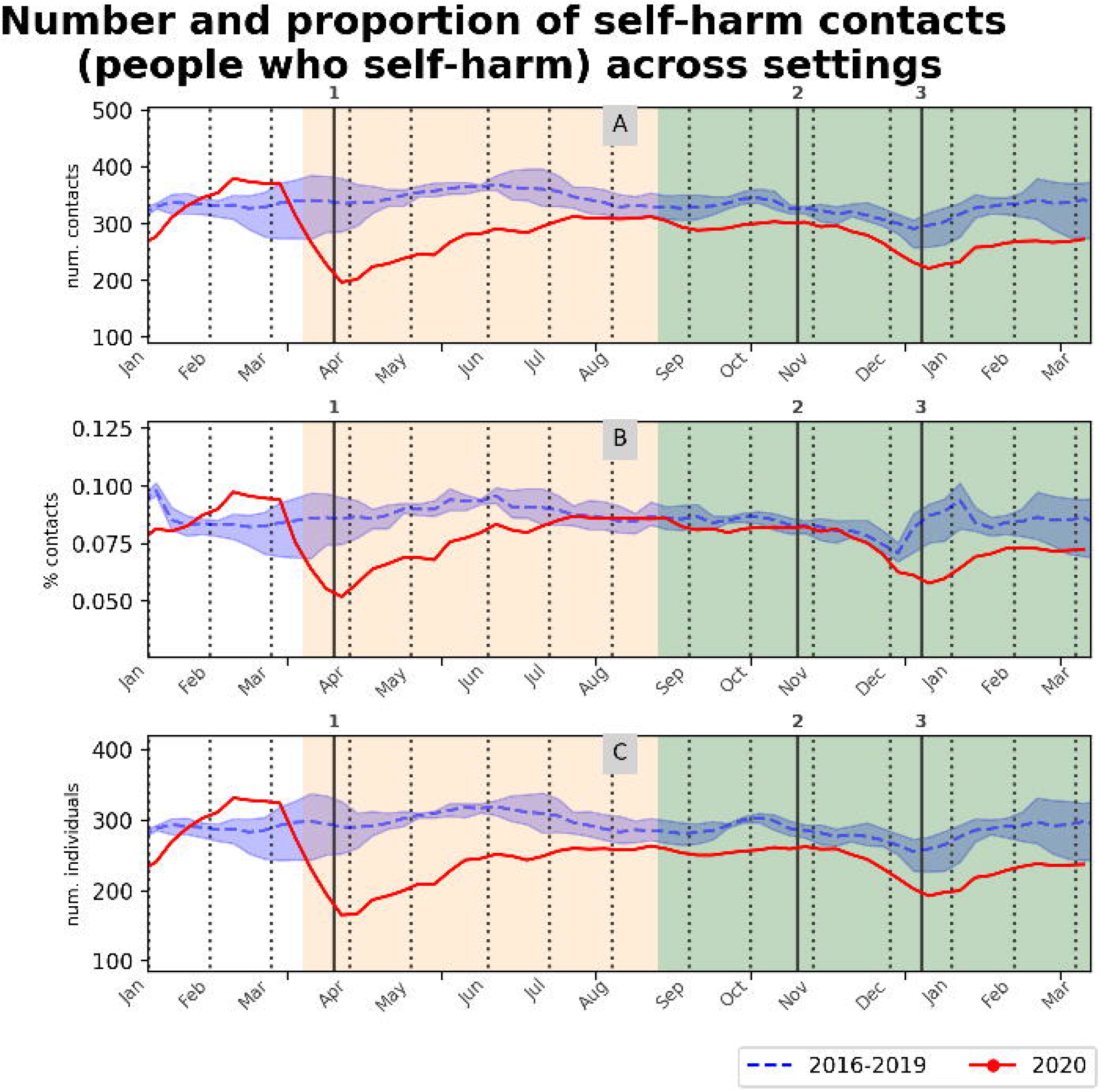
(A) Count and (B) proportion of weekly self-harm contacts in any setting (GP, ED or hospital admissions). (C) Weekly count of individuals with a self-harm contact in any setting. Solid red lines are 4-weeks rolling average of the weekly measurements for 2020. Blue dashed line and shaded area are average and min-max over the previous 4 years, 2016-2019. Changes in background shades correspond to before COVID-19, Wave 1 and Wave 2 periods respectively. Vertical lines are start stay-at-home measures during Wave 1 (1) and start of firebreak (2) and of stay-at-home (3) measures during Wave 2, in 2020.

The proportion of self-harm to all GP contacts was lower in March-June 2020 (mean ROR = 0.7, p < p<0.05 for 3/3 counterfactual years) and December 2020 (mean ROR = 0.7, p<0.05 for 3/3 counterfactual years) compared to previous years. The proportion of self-harm to all contacts in ED peaked in April 2020 (mean ROR = 1.3, p<0.05 for 2/3 counterfactual years) and October-November 2020 (mean ROR = 1.3, p<0.05 for 2/3 counterfactual years) above levels prior to the COVID-19 pandemic. The proportion of self-harm ED contacts admitted to hospital followed a downward trend from June 2020 (monthly reduction of 1.9 [1.5, 2.3] percentage points (pp), p<0.05 for 4/4 counterfactual years). This trend flattened during the start of stay-at-home measures of Wave 2, and there was no clear upward trend like those seen in other settings. The proportion of self-harm to all hospital admissions exhibited a long peak above previous levels during Wave 1 (mean ROR = 1.2, p<0.05 for 2/3 counterfactual years), and a downward trend from June 2020 (monthly reduction of 0.06 [0.07, 0.05] pp, p<0.05 for 4/4 counterfactual years). The proportion of these admissions transferred to critical care remained constant during the study period. Weekly number and single-setting proportions of self-harm contacts per setting can be seen in Figure 2 (detailed results in Suppl. Tables 2 and 3).

**Figure 2.**
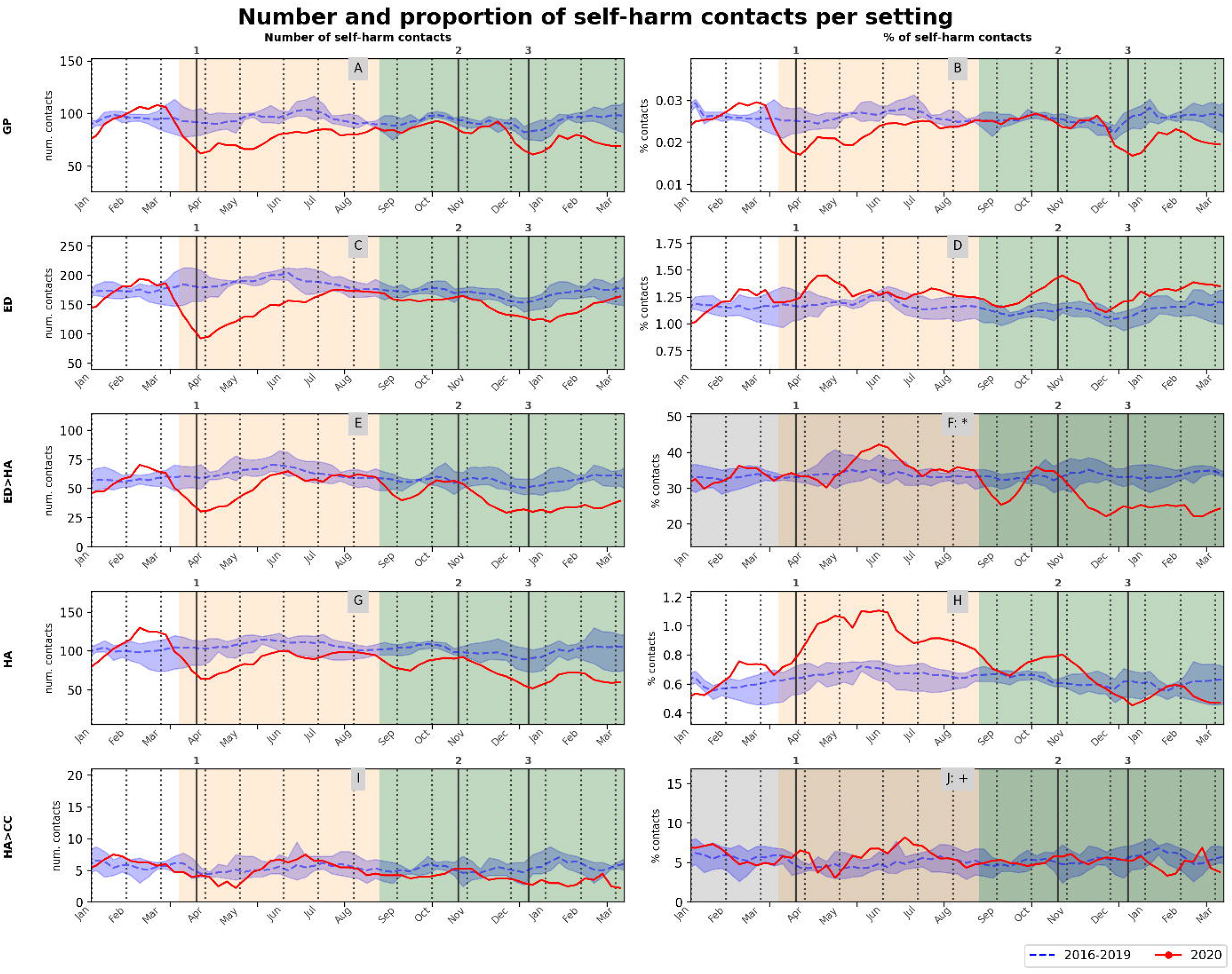
Count (left) and proportion (right) of weekly self-harm contacts in each setting – (A, B) GP, (C, D) ED, (E, F^*^) ED followed by hospital admissions (ED>HA), (G, H) hospital admissions (HA) and (I, J+) hospital admissions with a transfer to critical care (HA>CC). Solid red lines are 4-weeks rolling average of the weekly measurements for 2020. Blue dashed lines and shaded areas are average and min-max respectively over the previous 4 years, 2016-2019. Changes in background shades correspond to before COVID-19, Wave 1 and Wave 2 periods respectively. Vertical lines are start stay-at-home measures during Wave 1 (1) and start of firebreak (2) and of stay-at-home (3) measures during Wave 2, in 2020. Darker panels show proportion of ED presentation with self-harm that resulted in a hospital admission (F: ^*^) and of hospital admissions with self-harm that resulted in a transfer to critical care (J: +).

In April 2020, shortly after the start of Wave 1’s stay-at-home orders, people with self-harm contacts were more likely to be seen only in GP (mean ROR = 1.2, p<0.05 for 1/3 counterfactual years) or across all three settings (GP, ED and hospital admission; mean ROR = 2.3, p<0.05 for 3/3 counterfactual years), and less likely to be seen only in ED (mean ROR = 0.884, p<0.05 for 1/3 counterfactual years), than previous years. Towards the end of Wave 1 (July-August), more presented to ED (mean ROR = 1.1, p<0.05 for 3/3 counterfactual years), resulting in a decrease in the proportion seen only in GP (mean ROR = 0.818, p <0.05 for 2/3 counterfactual years) and an increase in the proportion seen in all three settings (mean ROR = 2.1, p<0.05 for 3/3 counterfactual years). Going into Wave 2 and up to December 2020, fewer individuals with self-harm contacts were admitted to hospital (monthly reduction of 1 [0.3, 1.7] pp, p<0.05 for 2/4 counterfactual years), resulting in fewer individuals been seen in ED and hospitalized (monthly reduction of 1.2 [0.6, 1.7] pp, p<0.05 for 4/4 counterfactual years) and more been seen only in ED (monthly increase of 1 [0.6, 1.3] pp, p<0.05 for 4/4 counterfactual years). Towards the end of the study period (February 2021), the proportion of those with self-harm contacts seen only in ED peaked above prior levels (mean ROR = 1.3, p<0.05 for 3/3 counterfactual years). Figure 3 shows how those with self-harm contacts distributed across settings (detailed results in Suppl. Tables 3 and 4).

**Figure 3:**
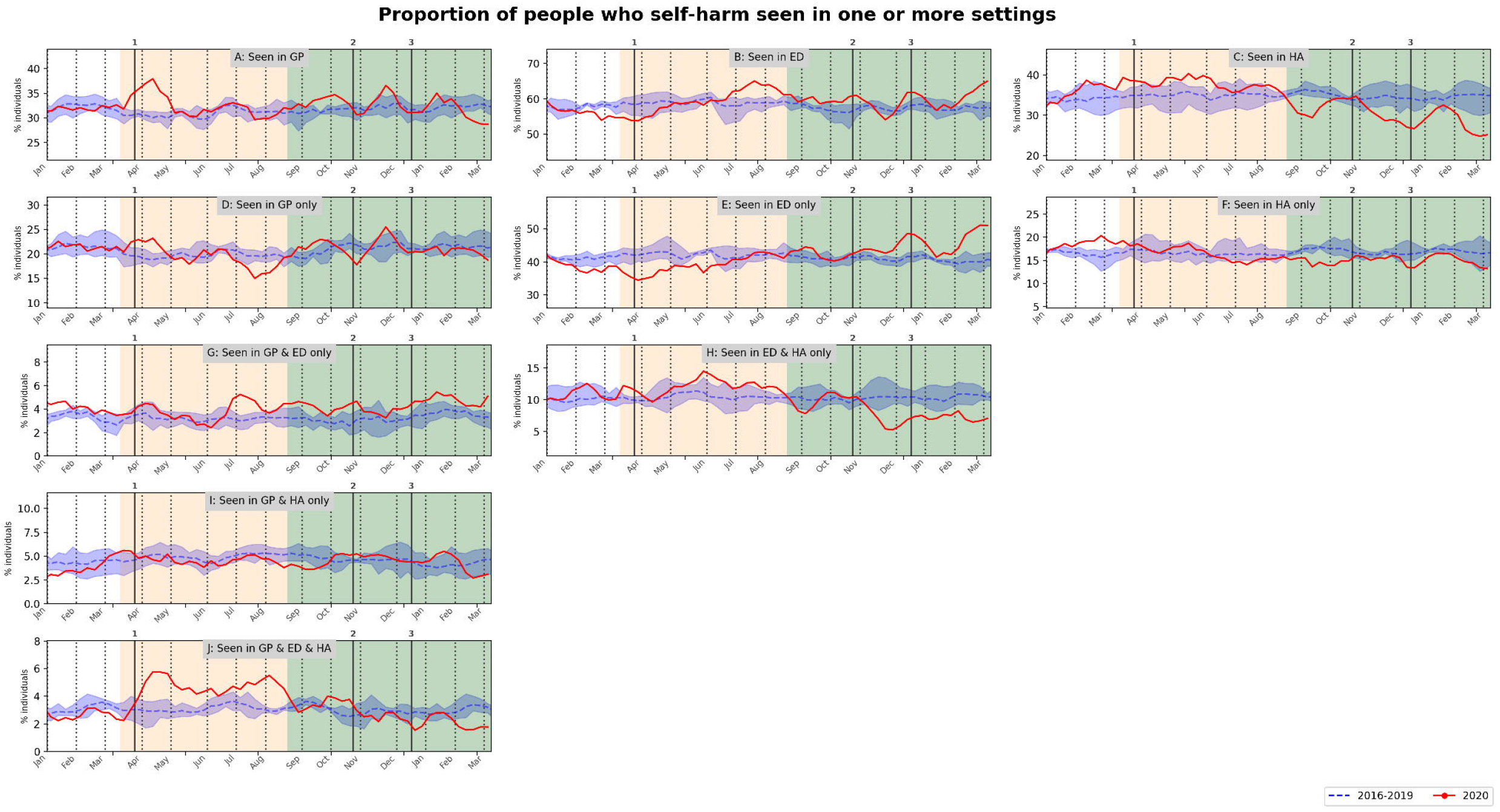
Weekly proportion of individuals with self-harm contacts seen in GP, ED and/or hospital admissions (HA). Solid red lines are 4-weeks rolling average of the weekly measurements for 2020. Blue dashed lines and shaded areas are average and min-max over the previous 4 years, 2016-2019. Panels A to C show overlapping sets. Panels D to J show non-overlapping sets. Changes in background shades correspond to before COVID-19, Wave 1 and Wave 2 periods respectively. Vertical lines are start stay-at-home measures during Wave 1 (1) and start of firebreak (2) and of stay-at-home (3) measures during Wave 2, in 2020.

The proportion of self-harm contacts due to burning increased in ED (mean ROR = 2.3 and 1.7 for Waves 1 and 2 respectively, p>0.05 for 4/4 counterfactual years) and decreased in hospital admissions (mean ROR = 0.7 and 0.1 for Waves 1 and 2 respectively, p>0.05 for 4/4 counterfactual years). The proportion of self-harm contacts due to hanging increased in Wave 1 (mean ROR = 2.0 and 1.4 for GP and hospital admissions respectively, p>0.05 for 4/4 counterfactual years) but not in Wave 2 (mean ROR = 0.8 and 1.0 for GP and hospital admissions respectively, p>0.05 for 4/4 counterfactual years). Contacts for self-harm due to other methods showed no change during the COVID-19 pandemic or their numbers where too small to test (Suppl. Table 5).

Further analysis of incidence and prevalence of self-harm contacts showed virtually identical patterns for new and recurrent events across all and for each setting (Suppl. Figure 11).

#### Sex, age and deprivation stratification

The peaks seen in the proportion of self-harm to all contacts in ED were largely the result of increases in those aged 10-24 years (mean ROR = 1.7, p<0.05 for 3/3 counterfactual years). In Wave 1, a slightly higher than usual proportion of females aged 10-24 years-old who self-harmed were admitted to hospital (mean ROR = 1.5 and 1.5 for age and sex comparisons respectively, p<0.05 for 1/3 counterfactual years), and a higher proportion of those with self-harm contacts were seen in both ED and hospital (unable to test due to low counts). In Wave 2, the number of self-harm hospital admissions in this group remained constant while it dropped for other sex-age groups (mean RRR = 1.4 for both age and sex comparisons, p<0.05 for 3/3 counterfactual years) - resulting in a complementary increase in the proportion of self-harm to all hospital admissions (mean ROR = 1.4 for both age and sex comparisons respectively, p<0.05 for 3/3 counterfactual years). Meanwhile, those aged >24 years-old exhibited a downward trend in the number of self-harm hospital admissions from June to December 2020, more so in males (monthly reduction 3.4 [2.7, 4.2] contacts, p<0.05 for 4/4 counterfactual years) than females (monthly reduction 2.1 [0.8, 3.3] contacts, p<0.05 for 1/4 counterfactual years). We found no changes in the gradient across WIMD deprivation quintiles. Detailed results stratified by age-sex can be found in Suppl. Figures 4-6 and Suppl. Tables 3, 6 and 7, and stratified by WIMD deprivation quintiles in Suppl. Figures 7-9 and Suppl. Tables 8 and 9. Further analysis of incidence and prevalence of self-harm contacts showed no differences across sex, age and WIMD deprivation quintiles (Suppl. Figures 12-13).

### Self-harm and COVID-19 infection association

We identified 33,257 individuals (0.9% of 3,552,210) with active (confirmed or suspected) or a history of COVID-19 infections between 28 February 2020 and 27 August 2020 (Suppl. Figure 1). Based on our inclusion criteria, 23,703 individuals were considered infected. Of these 23,703 individuals, 59.1% (13,999) were confirmed, 56.1% (13,291) suspected and 11.5% (2,721) had a history of infection within the ascertainment period (See Suppl. Figure 10). The proportions of individuals having a history of self-harm before the index date were 8.7% (2,070/23,703) and 5.6% (138,174/2,482,673) for the COVID-19 infected group and not infected group respectively (Suppl. Table 10). Logistic regression revealed self-harm history as a significant risk factor of COVID-19 infection in both unadjusted and adjusted analyses (adjusted OR: 1.4; 95% CI: 1.2-1.6; p<0.001), alongside being female, older age, non-White, higher deprivation and living in care homes (Figure 4A and Suppl. Table 11). From pre-COVID to post-COVID periods, incidence of self-harm decreased slightly in the not-infected group and increased slightly in the infected group (Figure 4B, Suppl. Table 12), resulting in a slight increase of the DID estimator (Figure 4C, Suppl. Table 13; DiD estimator = 1.1; 95% CI: 0.8-1.4; p = 0.729). Results from sensitivity analysis and robustness checks were similar to the main analysis (Figure 4C, Suppl. Figure 11, Suppl. Table 12-15).

**Figure 4.**
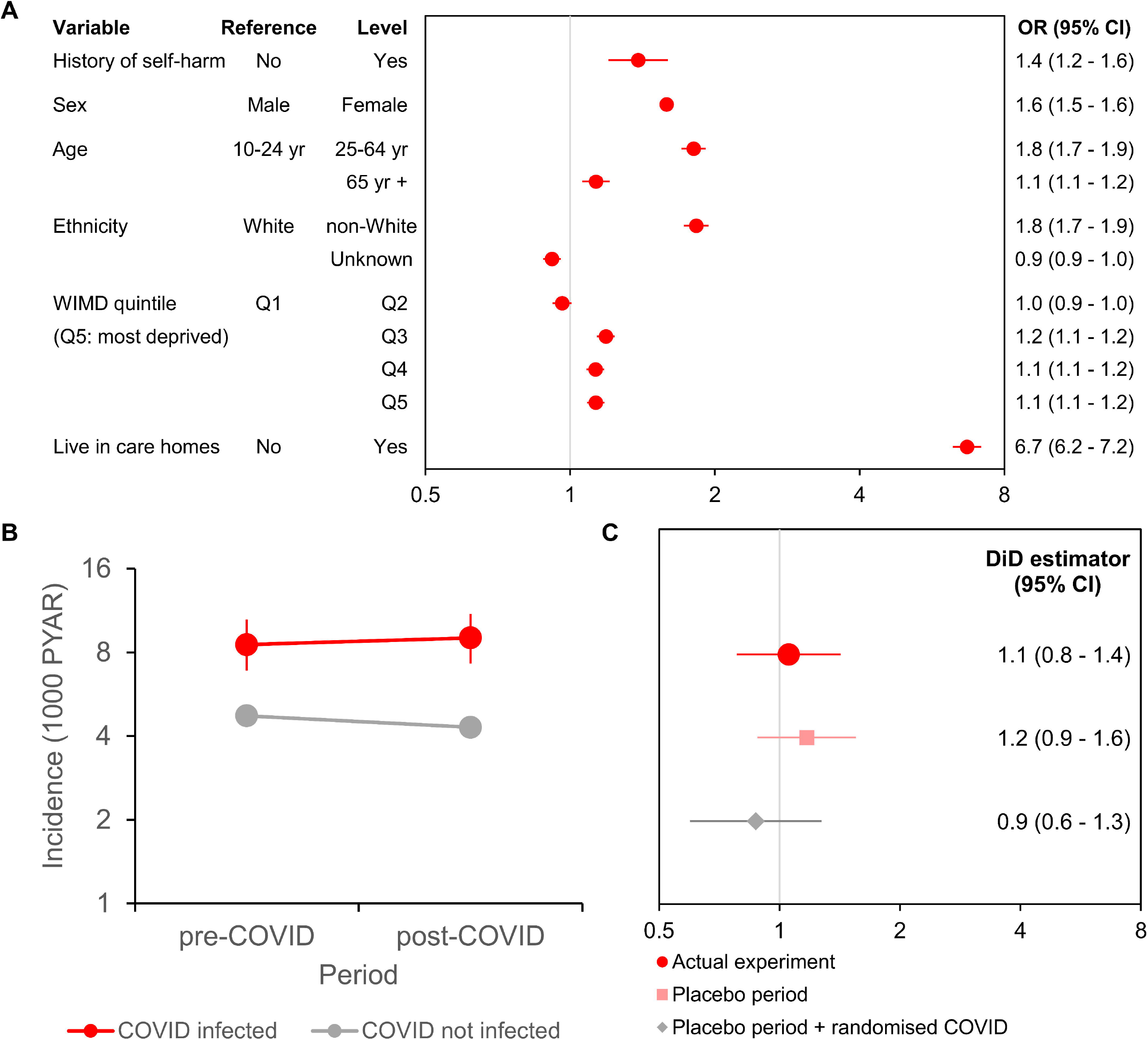
Results summary of self-harm and COVID-19 infection analyses. **(A)** Forest plot of the adjusted ORs of self-harm, sex, age, deprivation and living in care home for risk of COVID-19 infections. **(B)** Observed incidence of self-harm presentations during the pre-and post-COVID follow-up periods. **(C)** Forest plot of the difference-in-difference (DiD) estimators (with results from the two robustness checks, see Methods for details) for the risk of self-harm following COVID-19 infection. Error bars: 95% CIs.

## Discussion

The number of self-harm presentations across healthcare services was at a minimum at the start of stay-at-home restrictions and recovered compared to previous years in the following 3-5 months. This extends to Wave 2 of the COVID-19 pandemic results previously reported on Wave 1 in regions such as France,^25^ Italy,^26^ Ireland,^6^ Portugal,^27^ the UK,^3^ and the USA.^28^ We have also shown that this pattern took place across each individual setting (GP, ED and hospital inpatients) and in those with repeated self-harm presentations and with first presentations. The trough during Wave 2 was an amplified seasonal effect seen every Christmas and was slightly less pronounced than the minimum seen in Wave 1. An additional trough during August-September 2020 has been previously reported in combined data for Northern-Ireland, Scotland and Wales.^3^ However, we found no evidence of this in population-level Welsh data, suggesting that such drop may have been localized in the other two nations or an artefact. Crucially, incidence and prevalence rates of self-harm contacts did not exceed rates seen in previous years at any point during the study period (Waves 1 and 2), but continuous monitoring is necessary.

The number of GP contacts with self-harm reduced disproportionately during both Waves of the pandemic compared to other types of presentations. The same was not true in ED and hospital admissions, where despite lower numbers, at times during both waves self-harm contacts constituted a higher-than-usual proportion of all contacts – this is in line with and extends to Wave 2 the ED literature on Wave 1^26,28^ and may explain the perception by practitioners of an increase in self-harm in each setting. Nevertheless, more than usual of those who self-harmed during Wave 1 presented to GPs. Fear of contracting COVID-19 in ED and hospital settings and protecting emergency NHS provision may have resulted in patients preferentially presenting in GPs during Wave 1 but not during Wave 2.

Population-level unmet need related to the fall in self-harm presentations has been suggested with people unlikely to have received psychosocial assessments and interventions.^29^ We found that the proportion of self-harm ED contacts admitted to hospital and of self-harm hospital admissions transferred to critical care remained constant, except for a reduction in the proportion of subsequent hospitalisations during Wave 2. In keeping with the literature^25^ we also found some evidence that more lethal methods such as hanging increased during the pandemic, particularly during Wave 1. These results suggest more stringent criteria for hospitalisation following ED presentations with self-harm was potentially in operation, supporting such a view of unmet need. Most worryingly, the situation, far from improving during Wave 2, seemed to worsen, as the drop in the proportion of subsequent hospitalisations following ED presentation remained low even after an increase in the number of self-harm ED presentations.

### Sex, age and deprivation stratification

We found the largest differences between age-sex groups in ED and hospital admissions, and despite a larger drop in raw counts and incidence and prevalence rates, self-harm ED and hospital admissions by those aged 10-25 years dropped proportionally less during the pandemic than non-self-harm contacts compared to older patients. This was especially true for young females (10-24 years old). Meanwhile, less of those aged >24 years and that self-harmed during Wave 2 were hospitalized, particularly males. Stratified results from other published studies are highly heterogeneous, perhaps due to the relatively small samples used – usually collected in a single hospital compared to the population-based, multi-setting data analysed in the current study. A UK population-based study with data from combined primary and secondary care-treated episodes during Wave 1 found a most marked drop in the younger ages and most deprived areas.^3^ However, we found that such drop only altered the age gradient significantly in the instances described above, and never the deprivation gradient.

### Self-harm and COVID-19 infection association

We found that previous self-harm was associated with a higher risk (∼40%) of COVID-19 infection and that risk of self-harm was also elevated (∼5-10%) following infection. Although these effects are modest compared to bidirectional associations in psychiatric conditions, the direction of effects remained the same.^10^ These could be expected given the correlation between self-harm and mental illnesses and the neurobiological effects of COVID-19.^8,10^ Our results were not statistically robust across primary and sensitivity analyses due to small sample sizes in the infected group. However, they are consistent (albeit smaller) with a recent UK population online survey who reported unadjusted associations,^9^ and with the association between self-harm and engagement in risky behaviours that may also increase the risk of COVID-19 infection.^30^

## Strengths and limitations

We have presented for the first time a detailed picture of the effects of Waves 1 and 2 of the COVID-19 crisis on self-harm contacts across GP, ED and hospital admissions by linking individual-level routinely collected data covering the entire population of Wales for over five years (2016-01-01 to 2021-03-14). We examined trajectory changes of number, incidence, prevalence and proportion of healthcare service contacts with self-harm, as well as changes in the distribution of self-harm presentations across settings. This is crucial given how evenly distributed contacts are across the three settings (1/3 in GP, 2/3 in ED and 1/3 admitted to hospital; Figure 3). We additionally studied bidirectional associations between self-harm and COVID-19 infection at population level. Such comprehensive analysis has not been reported before and allowed us to identify and quantify for the first time differences across settings and waves in how the COVID-19 pandemic affected self-harm presentations.

We adopted the DiD approach, a widely used quasi-experimental method to compare changes between pre-and post-COVID outcomes to respective changes in previous years.^23^ Given all the assumptions of the DiD approached are met, causal inference could be strengthened from our findings, but caution is required. The assumptions of common trend, correct specification of functional form of trends and no unmeasured confounding may not be valid nor easily verified.^23,24^ We used contacts in previous years as counterfactual when both COVID-19 outbreak and lockdown were absent. Thus, our DiD analyses did not disentangle the effects of social/medical isolation from the pandemic.^31^

Small numbers prevented us from running statistical tests on hospital admissions with critical care transfers and on some method and sex-age stratified analyses. Results on the relationship between self-harm presentation and COVID-19 infection are inconclusive, limited by statistical power and are preliminary. The assumptions of using DiD for improving causal inference may be violated (e.g., omission of key socioeconomic factors)^32^ thus interpretation of the relative risks requires extra caution.^23^ Further research to reduce selection bias by, e.g., large-scale infection survey with appropriate sampling strategies is warranted.^33^

Limitations of the use of routinely collected data for research purposes have been reported elsewhere.^34^ Misclassification bias was reduced using validated definitions of contacts and code lists.^20,21^ To reduce the underestimation of COVID-19 infection due to incomplete coverage of the LIMS datasets,^14^ we also identified COVID-19 infections from the general population who contacted primary and secondary care services.

### Implications for research, policy and practice

This study sheds some light on how the COVID-19 pandemic and the measures taken to curb its spread during Waves 1 and 2 affected healthcare service presentations with self-harm across GP, ED and hospitalisations. The fear of infection, stay at home orders and ‘protect the NHS’ may have prevented those who self-harm from accessing healthcare services, particularly primary care, where we saw the largest drop in the number of self-harm contacts. This is especially worrying since between 15% and 25% of those presenting with self-harm do so to GP only, and indeed, despite the larger drop, GPs were more commonly contacted than usual during Wave 1. This may be the result of those with potentially ‘less severe’ self-harm (i.e., those not requiring ED care or hospital admission) not seeking help during the pandemic as often as before. However, methods and suicidal motivations for self-harm fluctuate, therefore it is important that those who self-harm receive appropriate assessment and intervention.^35^ We have highlighted the need to pay special attention to children and young people, particularly females in light of increases seen in incidence of self-harm in adolescent females before the pandemic.^36^ Not providing the support needed for those who self-harm will likely have negative implications for individuals and health services.^29^ Further research on morbidity and mortality, including suicide, in those with a history of self-harm as well as those who also had COVID-19 will highlight whether this is the case and to what extent.

## Supporting information

RECORD Checklist

Supplementary Results

## Data Availability

The data used in this study are available in the SAIL Databank at Swansea University, Swansea, UK, but as restrictions apply, they are not publicly available. All proposals to use SAIL data are subject to review by an independent Information Governance Review Panel (IGRP). Before any data can be accessed, approval must be given by the IGRP. The IGRP gives careful consideration to each project to ensure proper and appropriate use of SAIL data. When access has been granted, it is gained through a privacy protecting safe haven and remote access system referred to as the SAIL Gateway. SAIL has established an application process to be followed by anyone who would like to access data via SAIL at https://www.saildatabank.com/application-process. Derived data supporting the findings of this study are available from the corresponding author (AJ) on request.

## Data availability

The data used in this study are available in the SAIL Databank at Swansea University, Swansea, UK, but as restrictions apply, they are not publicly available. All proposals to use SAIL data are subject to review by an independent Information Governance Review Panel (IGRP). Before any data can be accessed, approval must be given by the IGRP. The IGRP gives careful consideration to each project to ensure proper and appropriate use of SAIL data. When access has been granted, it is gained through a privacy protecting safe haven and remote access system referred to as the SAIL Gateway. SAIL has established an application process to be followed by anyone who would like to access data via SAIL at https://www.saildatabank.com/application-process. Derived data supporting the findings of this study are available from the corresponding author (MDPB) on request.

## Acknowledgements

This study makes use of anonymized data held in the Secure Anonymised Information Linkage (SAIL) Databank. This work uses data provided by patients and collected by the NHS as part of their care and support. We would also like to acknowledge all data providers who make anonymized data available for research. We wish to acknowledge the collaborative partnership that enabled acquisition and access to the de-identified data, which led to this output. The collaboration was led by the Swansea University Health Data Research UK team under the direction of the Welsh Government Technical Advisory Cell (TAC) and includes the following groups and organizations: the SAIL Databank, Administrative Data Research (ADR) Wales, Digital Health and Care Wales (DHCW), Public Health Wales, NHS Shared Services Partnership (NWSSP) and the Welsh Ambulance Service Trust (WAST). All research conducted has been completed under the permission and approval of the SAIL independent Information Governance Review Panel (IGRP) project number 0911.

## Contributions

All authors were responsible and accountable to all part of works related to the study. AJ conceived the study. AJ, RAL, AA and FT acquired funding. MDPB, LL, YF and AJ contributed to the design of the study. MDPB, LL and YF prepared and analysed the data. MDPB, LL, YF and AJ produced the first draft. All authors interpreted the data, contributed to writing and revised the manuscript, and gave the approval to the final version to be published.

## References

1. Gunnell, D., Appleby, L., Arensman, E., Hawton, K., John, A., Kapur, N., & Yip, P. S. (2020). Suicide risk and prevention during the COVID-19 pandemic. The Lancet Psychiatry, 7(6), 468–471.

2. John A, Eyles E, Webb RT et al. The impact of the COVID-19 pandemic on self-harm and suicidal behaviour: update of living systematic review [version 2; peer review: 1 approved, 2 approved with reservations]. F1000Research 2021, 9:1097

3. Carr, M. J., Steeg, S., Webb, R. T., Kapur, N., Chew-Graham, C. A., Abel, K. M., & Ashcroft, D. M. (2021). Effects of the COVID-19 pandemic on primary care-recorded mental illness and selfharm episodes in the UK: a population-based cohort study. The Lancet Public Health.

4. Rhodes, H. X., Petersen, K., & Biswas, S. (2020). Trauma trends during the initial peak of the COVID-19 pandemic in the midst of lockdown: experiences from a rural trauma center. Cureus, 12(8).

5. Karakasi MV, Zaoutsou A, Theofilidis A, et al. Impact of the SARS-CoV-2 pandemic on psychiatric emergencies in northern Greece: Preliminary study on a sample of the Greek population. Psychiatry and Clinical Neurosciences. 2020 Nov;74(11):613-615. DOI: 10.1111/pcn.13136.

6. McIntyre, A., Tong, K., McMahon, E., & Doherty, A. (2020). COVID-19 and its effect on emergency presentations to a tertiary hospital with self-harm in Ireland. Irish Journal of Psychological Medicine, 1–7. doi:10.1017/ipm.2020.116

7. Wind TR, Rijkeboer M, Andersson G, Riper H. The COVID-19 pandemic: The ‘black swan’ for mental health care and a turning point for e-health. Internet Interv. 2020;20:100317. doi:10.1016/j.invent.2020.100317

8. Niederkrotenthaler T, Gunnell D, Arensman E, Pirkis J, Appleby L, Hawton K, John A, Kapur N, Khan N, C. O’Connor R, Platt S, The International COVID-19 Suicide Prevention Research (2020). Suicide Research, Prevention, and COVID-19. Crisis. 1-10. DOI:https://doi.org/10.1027/0227-5910/a000731

9. Iob, E., Steptoe, A., & Fancourt, D. (2020). Abuse, self-harm and suicidal ideation in the UK during the COVID-19 pandemic. The British Journal of Psychiatry, 217(4), 543–546. doi:10.1192/bjp.2020.130

10. Taquet, M., Luciano, S., Geddes, J. R., & Harrison, P. J. (2021). Bidirectional associations between COVID-19 and psychiatric disorder: retrospective cohort studies of 62 354 COVID-19 cases in the USA. The Lancet Psychiatry, 8(2), 130–140.

11. Ford DV, Jones KH, Verplancke JP, et al. The SAIL Databank: building a national architecture for e-health research and evaluation. BMC Health Services Research 2009; 9:157 doi:10.1186/1472-6963-9-157

12. Lyons, R. A., Jones, K. H., John, G., Brooks, C. J., Verplancke, J. P., Ford, D. V., & Leake, K. (2009). The SAIL databank: linking multiple health and social care datasets. BMC medical informatics and decision making, 9(1), 1–8.

13. Lyons, J., Akbari, A., Torabi, F., Davies, G. I., North, L., Griffiths, R., & Lyons, R. (2020). Understanding and responding to COVID-19 in Wales: protocol for a privacy-protecting data platform for enhanced epidemiology and evaluation of interventions. BMJ open, 10(10), e043010.

14. UK Government (2020). Covid-19 testing data: methodology note. Guidance. https://www.gov.uk/government/publications/coronavirus-covid-19-testing-data-methodology/covid-19-testing-data-methodology-note last accessed 5 August 2020.

15. NHS Wales (2020) COVID-19, High risk shielded patient list identification methodology. https://nwis.nhs.wales/coronavirus/coronavirus-content/coronavirus-documents/covid-19-high-risk-shielded-patient-list-identification-methodology/ last accessed 5 August 2020.

16. BBC (2020) https://www.bbc.co.uk/news/uk-wales-51450517 last accessed 5 August 2020.

17. HDR UK (2020) COVID-19 Phenomics. https://covid19-phenomics.org last accessed 5 August 2020.

18. NHS Digital (2020). COVID-19 National Clinical Coding Standard and Guidance. https://hscic.kahootz.com/t_c_home/view?objectID=19165328 last accessed 5 August 2020.

19. Welsh Government (2014). Welsh Index of Multiple Deprivation (WIMD) 2014 Revised. https://gov.wales/sites/default/files/statistics-and-research/2019-04/welsh-index-of-multiple-deprivation-2014-revised.pdf

20. John, A., DelPozo-Banos, M., Gunnell, D., Dennis, M., Scourfield, J., Ford, D. V., & Lloyd, K. (2020). Contacts with primary and secondary healthcare prior to suicide: case–control whole-population-based study using person-level linked routine data in Wales, UK, 2000–2017. The British Journal of Psychiatry, 1–8.

21. Marchant A, Turner S, Balbuena L, et al. Self-harm presentation across healthcare settings by sex in young people: an e-cohort study using routinely collected linked healthcare data in Wales, UK Archives of Disease in Childhood 2020;105:347–354.

22. Welsh Government (2020). Written Statement: Coronavirus (COVID-19) https://gov.wales/written-statement-coronavirus-covid-19-2 last accessed 5 August 2020.

23. Wing, C., Simon, K., & Bello-Gomez, R. A. (2018). Designing difference in difference studies: best practices for public health policy research. Annual review of public health, 39.

24. Dimick, J. B., & Ryan, A. M. (2014). Methods for evaluating changes in health care policy: the difference-in-differences approach. Jama, 312(22), 2401–2402.

25. Jollant, F., Roussot, A., Corruble, E., Chauvet-Gelinier, J. C., Falissard, B., Mikaeloff, Y., & Quantin, C. (2021). Hospitalization for self-harm during the early months of the COVID-19 pandemic in France: A nationwide retrospective observational cohort study. The Lancet Regional Health-Europe, 6, 100102.

26. Enrico Capuzzi, Carmen Di Brita, Alice Caldiroli, Fabrizia Colmegna, Roberto Nava, Massimiliano Buoli, Massimo Clerici, Psychiatric emergency care during Coronavirus 2019 (COVID 19) pandemic lockdown: results from a Department of Mental Health and Addiction of northern Italy, Psychiatry Research, Volume 293, 2020, 113463, ISSN 0165-1781, https://doi.org/10.1016/j.psychres.2020.113463

27. Gonçalves-Pinho, M., Mota, P., Ribeiro, J. et al. The Impact of COVID-19 Pandemic on Psychiatric Emergency Department Visits – A Descriptive Study. Psychiatr Q (2020). https://doi.org/10.1007/s11126-020-09837-z

28. Laura E. Walker, Heather A. Heaton, Ryan J. Monroe, R. Ross Reichard, Monica Kendall, Aidan F. Mullan, Deepi G. Goyal, Impact of the SARS-CoV-2 Pandemic on Emergency Department Presentations in an Integrated Health System, Mayo Clinic Proceedings, Volume 95, Issue 11, 2020, Pages 2395–2407, ISSN 0025-6196, https://doi.org/10.1016/j.mayocp.2020.09.019

29. Mansfield, K. E., Mathur, R., Tazare, J., Henderson, A. D., Mulick, A. R., Carreira, H., & Langan, S. M. (2021). Indirect acute effects of the COVID-19 pandemic on physical and mental health in the UK: a population-based study. The Lancet Digital Health, 3(4), e217–e230.

30. Shafti, M., Taylor, P. J., Forrester, A., & Pratt, D. (2021). The co-occurrence of self-harm and aggression: a cognitive-emotional model of dual-harm. Frontiers in psychology, 12, 415.

31. Lo, Y. F., Yang, F. C., Huang, J. S., Lin, Y. S., & Liang, C. S. (2021). Disentangling the complex bidirectional associations between COVID-19 and psychiatric disorder. The Lancet Psychiatry, 8(3), 179.

32. Yolken R. COVID-19 and psychiatry: can electronic medical records provide the answers? The Lancet Psychiatry. 2021 Feb 1;8(2):89–91. Pierce, M., McManus, S., Jessop, C., John, A., Hotopf, M., Ford, T., & Abel, K. M. (2020). Says who? The significance of sampling in mental health surveys during COVID-19. The Lancet Psychiatry, 7(7), 567–568. https://doi.org/10.1016/S2215-0366(20)30237-6

33. Pirkis J, Nicholas A, Gunnell D. The case for case–control studies in the field of suicide prevention. Epidemiology and psychiatric sciences. 2019:1–3.

34. Miller, M., Hempstead, K., Nguyen, T., Barber, C., Rosenberg-Wohl, S., & Azrael, D. (2013). Method choice in nonfatal self-harm as a predictor of subsequent episodes of self-harm and suicide: implications for clinical practice. American journal of public health, 103(6), e61–e68.

35. Cybulski, L., Ashcroft, D. M., Carr, M. J., Garg, S., Chew-Graham, C. A., Kapur, N., & Webb, R. T. (2021). Temporal trends in annual incidence rates for psychiatric disorders and self-harm among children and adolescents in the UK, 2003–2018. BMC psychiatry, 21(1), 1–12.

