## Supplementary material for "Healthcare presentations with self-harm and the association with COVID-19: an e-cohort whole-population-based study using individual-level linked routine electronic health records in Wales, UK, 2016 - March 2021": Figures

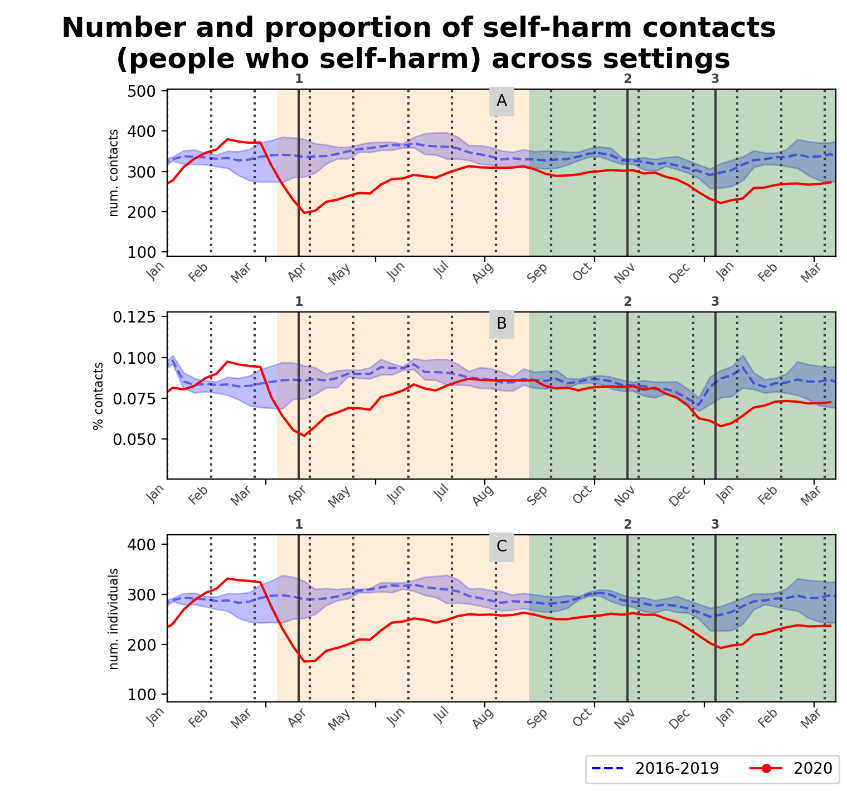

**Figure 1.** (A) Count and (B) proportion of weekly self-harm contacts in any setting (GP, ED or hospital admissions). (C) Weekly count of individuals with a self-harm contact in any setting. Solid red lines are 4-weeks rolling average of the weekly measurements for 2020. Blue dashed line and shaded area are average and min-max over the previous 4 years, 2016-2019. Changes in background shades correspond to before COVID-19, Wave 1 and Wave 2 periods respectively. Vertical lines are start stay-at-home measures during Wave 1 (1) and start of firebreak (2) and of stay-at-home (3) measures during Wave 2, in 2020.

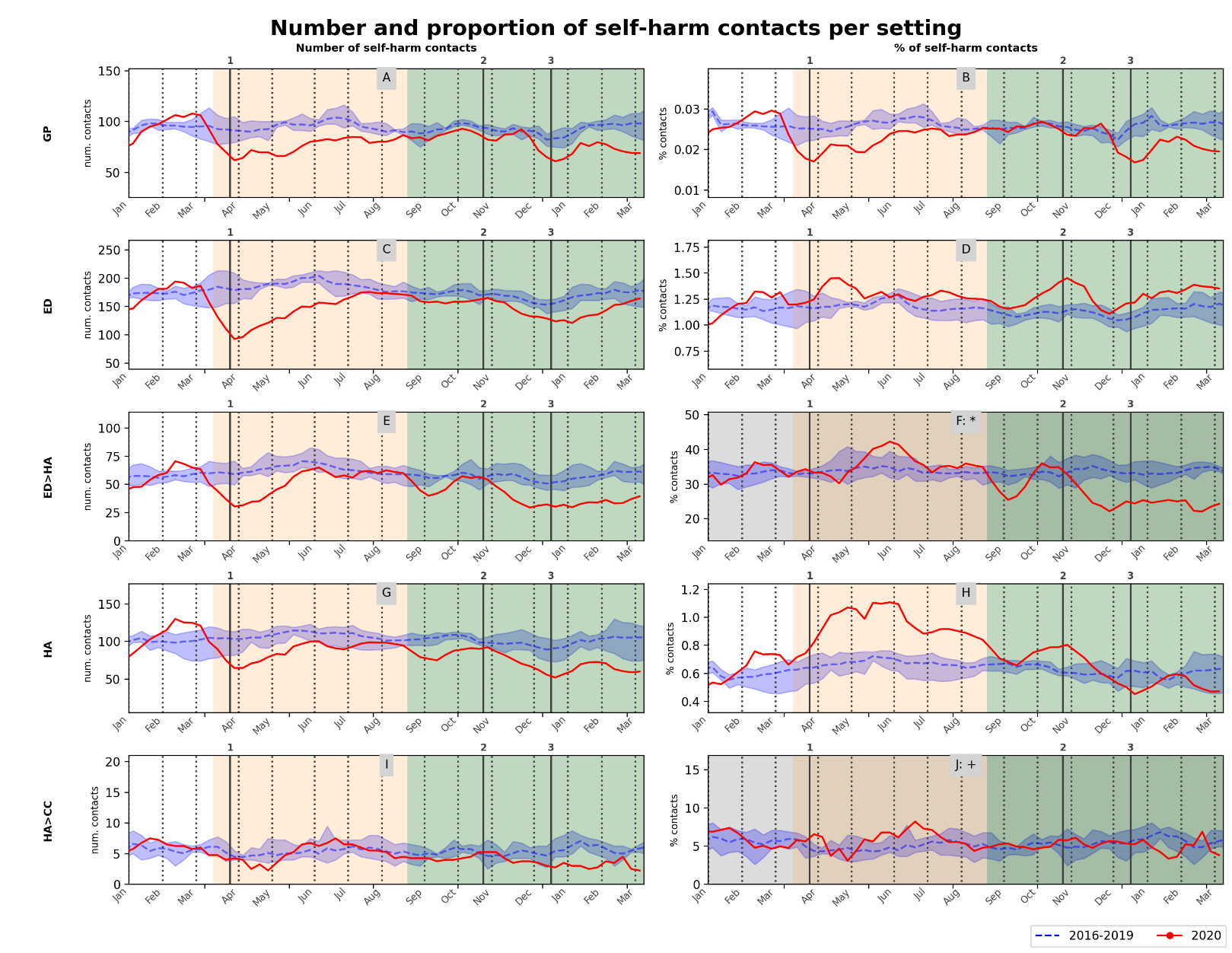

**Figure 2.** Count (left) and proportion (right) of weekly self-harm contacts in each setting – (A, B) GP, (C, D) ED, (E, F*) ED followed by hospital admissions (ED>HA), (G, H) hospital admissions (HA) and (I, J+) hospital admissions with a transfer to critical care (HA>CC). Solid red lines are 4-weeks rolling average of the weekly measurements for 2020. Blue dashed lines and shaded areas are average and min-max respectively over the previous 4 years, 2016-2019. Changes in background shades correspond to before COVID-19, Wave 1 and Wave 2 periods respectively. Vertical lines are start stay-at-home measures during Wave 1 (1) and start of firebreak (2) and of stay-at-home (3) measures during Wave 2, in 2020. Darker panels show proportion of ED presentation with self-harm that resulted in a hospital admission (F: *) and of hospital admissions with self-harm that resulted in a transfer to critical care (J: +).

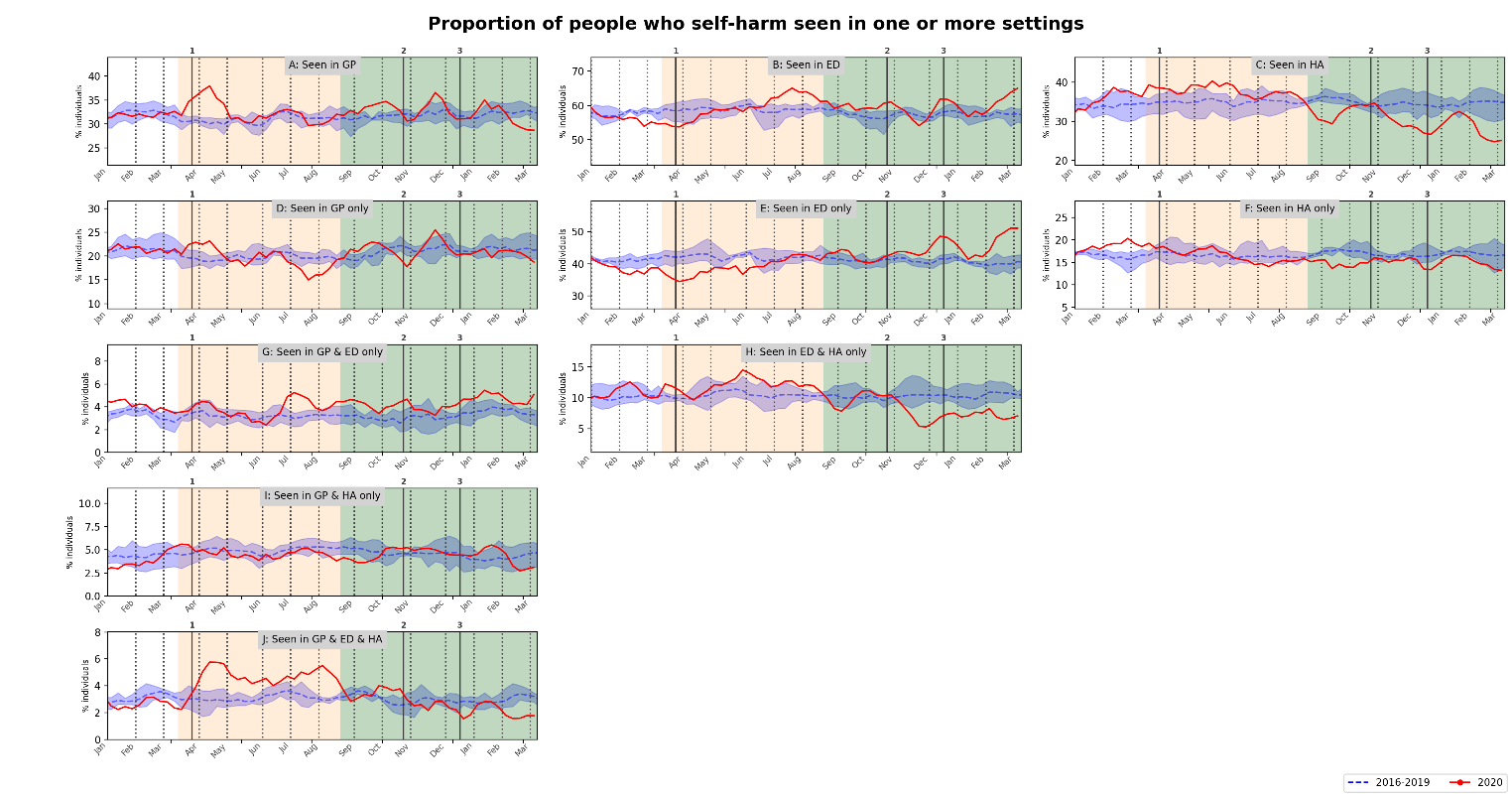

**Figure 3:** Weekly proportion of individuals with self-harm contacts seen in GP, ED and/or hospital admissions (HA). Solid red lines are 4-weeks rolling average of the weekly measurements for 2020. Blue dashed lines and shaded areas are average and min-max over the previous 4 years, 2016-2019. Panels A to C show overlapping sets. Panels D to J show non-overlapping sets. Changes in background shades correspond to before COVID-19, Wave 1 and Wave 2 periods respectively. Vertical lines are start stay-at-home measures during Wave 1 (1) and start of firebreak (2) and of stay-at-home (3) measures during Wave 2, in 2020.

| **A** | 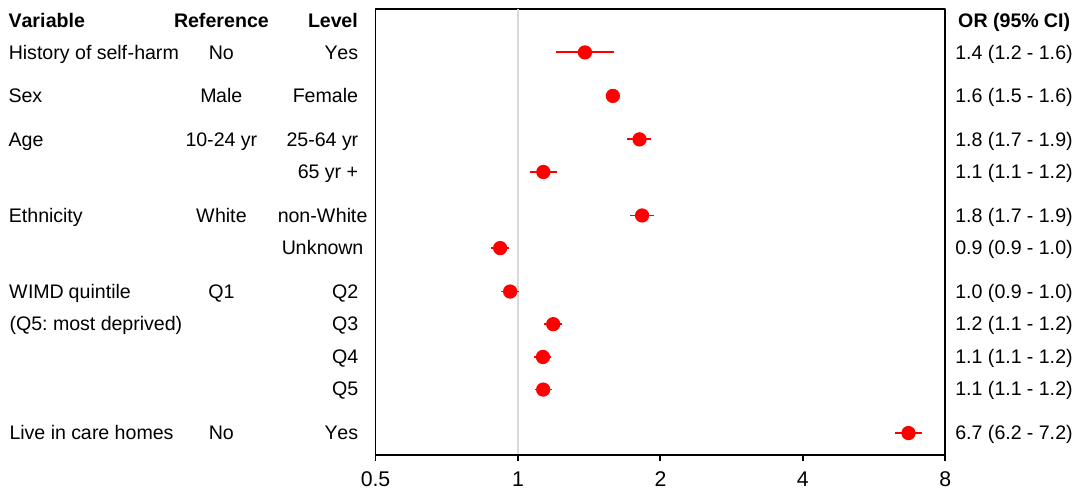 | | |
| --- | --- | --- | --- |
| **B** | 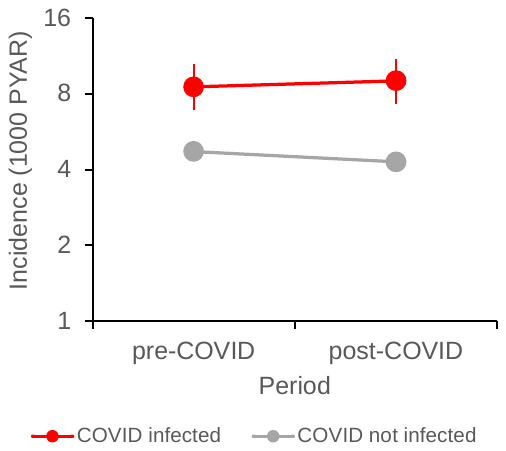 | **C** | 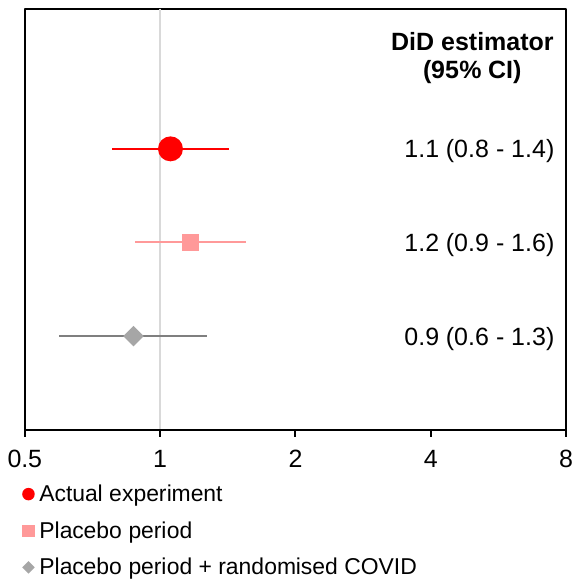 |

**Figure 4**. Results summary of self-harm and COVID-19 infection analyses. (**A**) Forest plot of the adjusted ORs of self-harm, sex, age, deprivation and living in care home for risk of COVID-19 infections. (**B**) Observed incidence of self-harm presentations during the pre- and post-COVID follow-up periods. (**C**) Forest plot of the difference-in-difference (DiD) estimators (with results from the two robustness checks, see Methods for details) for the risk of self-harm following COVID-19 infection. Error bars: 95% CIs.
