## Supplementary Results for "Healthcare presentations with self-harm and the association with COVID-19: an e-cohort whole-population-based study using individual-level linked routine electronic health records in Wales, UK, 2016 - March 2021"

**SUPPLEMENTARY FIGURES & TABLES**

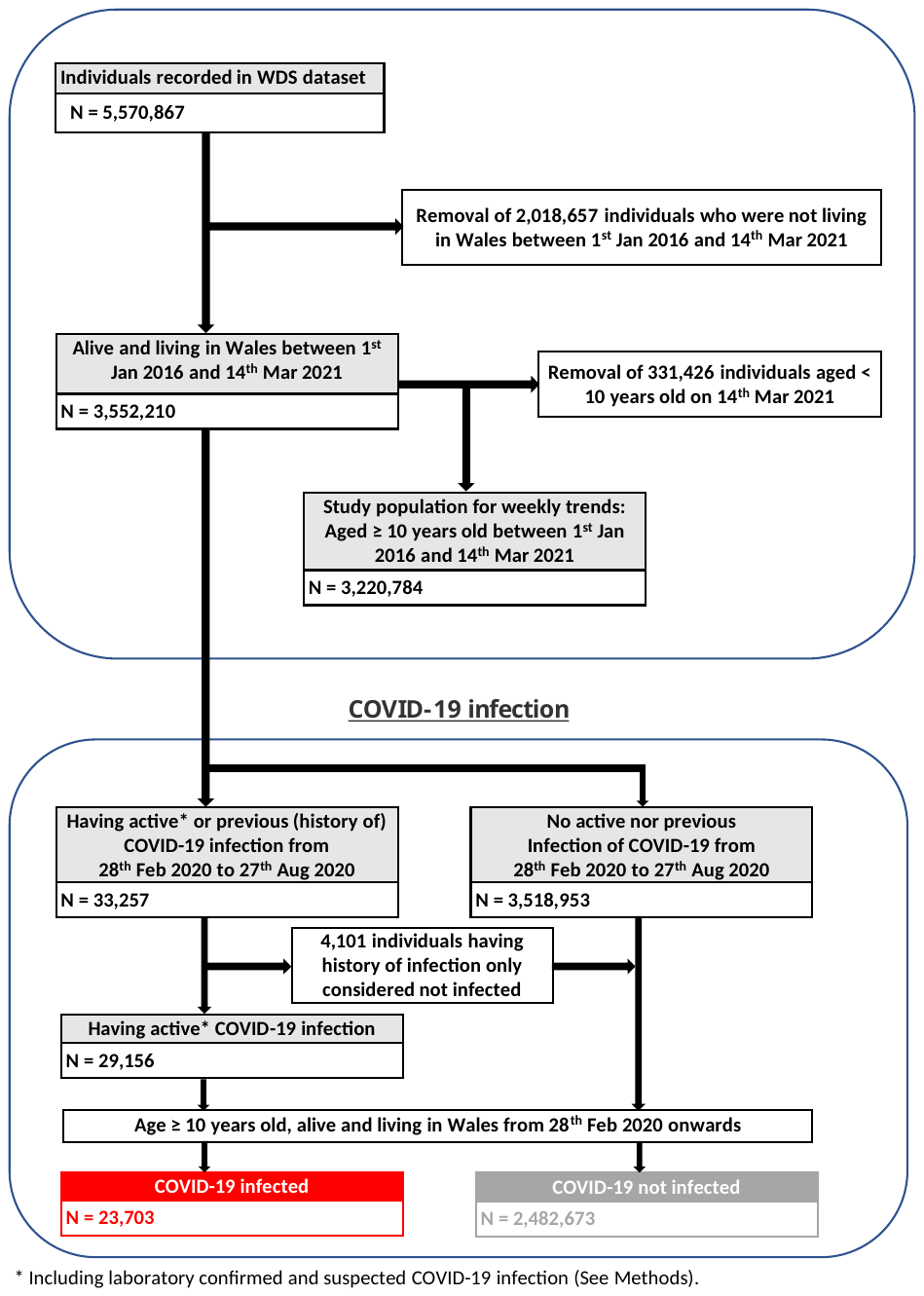

**Supplementary Figure 1.** Study population flow diagram. Inclusion and exclusion decisions leading to the creation of our study population (top) and the cohort used for analysis of self-harm after COVID-19 testing/infection (bottom).

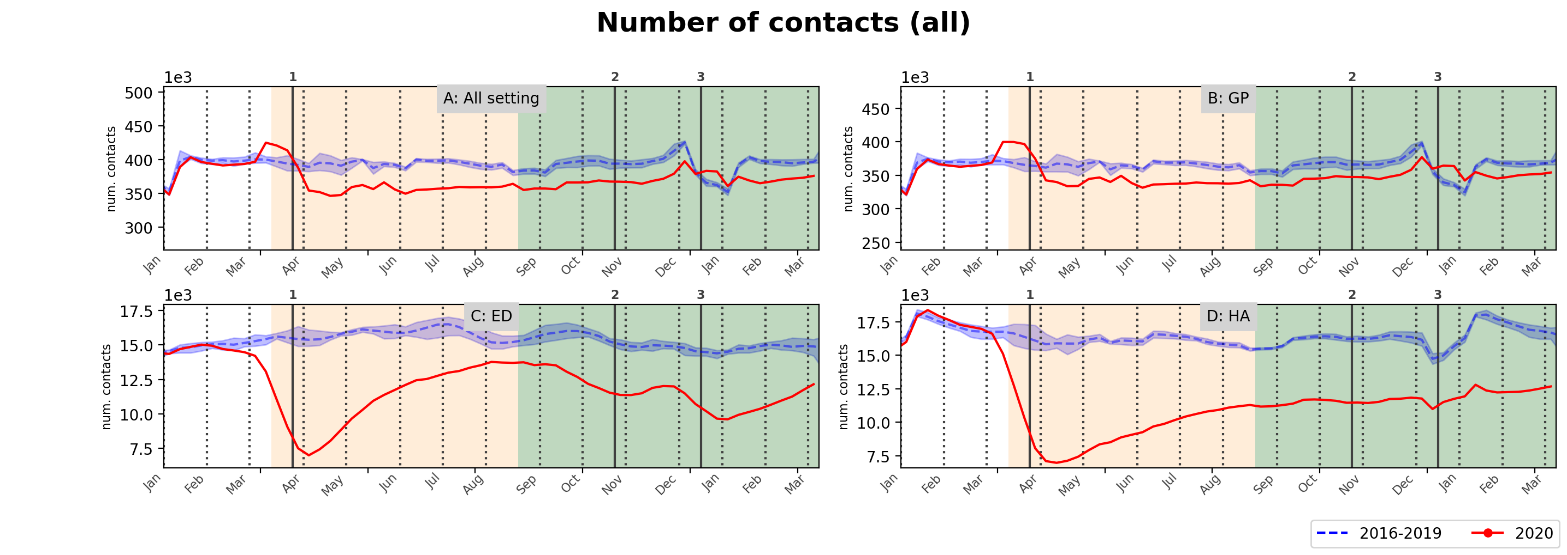

**Supplementary Figure 2.** Weekly number of contacts (A) across all settings, (B) GP, (C) ED, and (D) hospital admissions (HA). Solid red lines are 4-weeks rolling average of the weekly measurements for 2020. Blue dashed line and shaded area are average and min-max over the previous 4 years, 2016-2019. Changes in background shades correspond to before COVID-19, Wave 1 and Wave 2 periods respectively. Vertical lines are start stay-at-home measures during Wave 1 (1) and start of firebreak (2) and of stay-at-home (3) measures during Wave 2, in 2020.

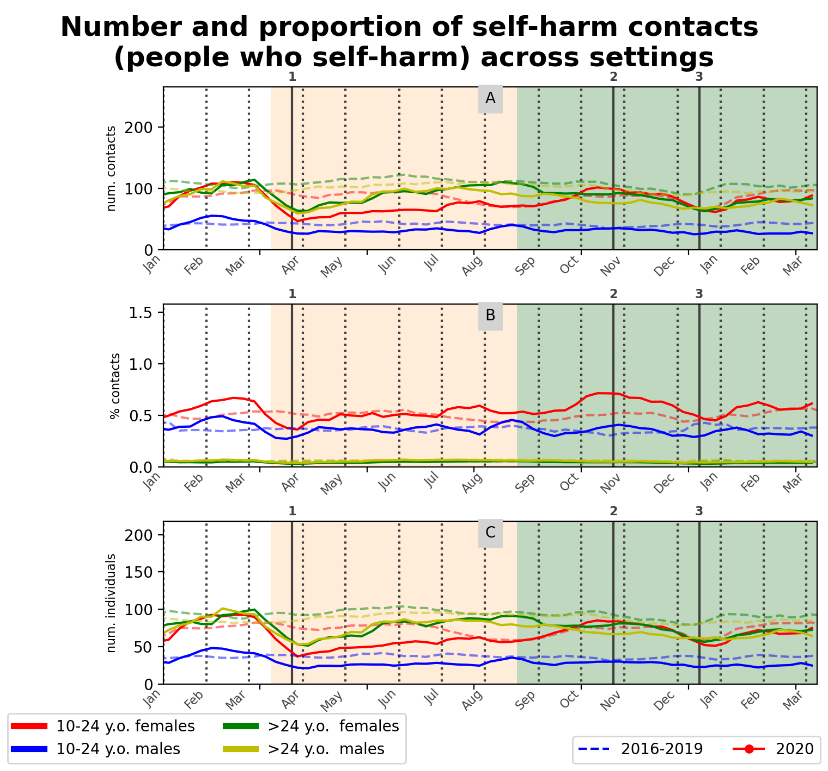

**Supplementary Figure 3.** (A) Count and (C) proportion of weekly self-harm contacts in any setting (GP, ED or hospital admissions) stratified by sex-age groups. (C) Weekly count of individuals with a self-harm contact in any setting stratified by sex-age groups. Solid red lines are 4-weeks rolling average of the weekly measurements for 2020. Blue dashed lines are averages over the previous 4 years, 2016-2019. Changes in background shades correspond to before COVID-19, Wave 1 and Wave 2 periods respectively. Vertical lines are start stay-at-home measures during Wave 1 (1) and start of firebreak (2) and of stay-at-home (3) measures during Wave 2, in 2020.

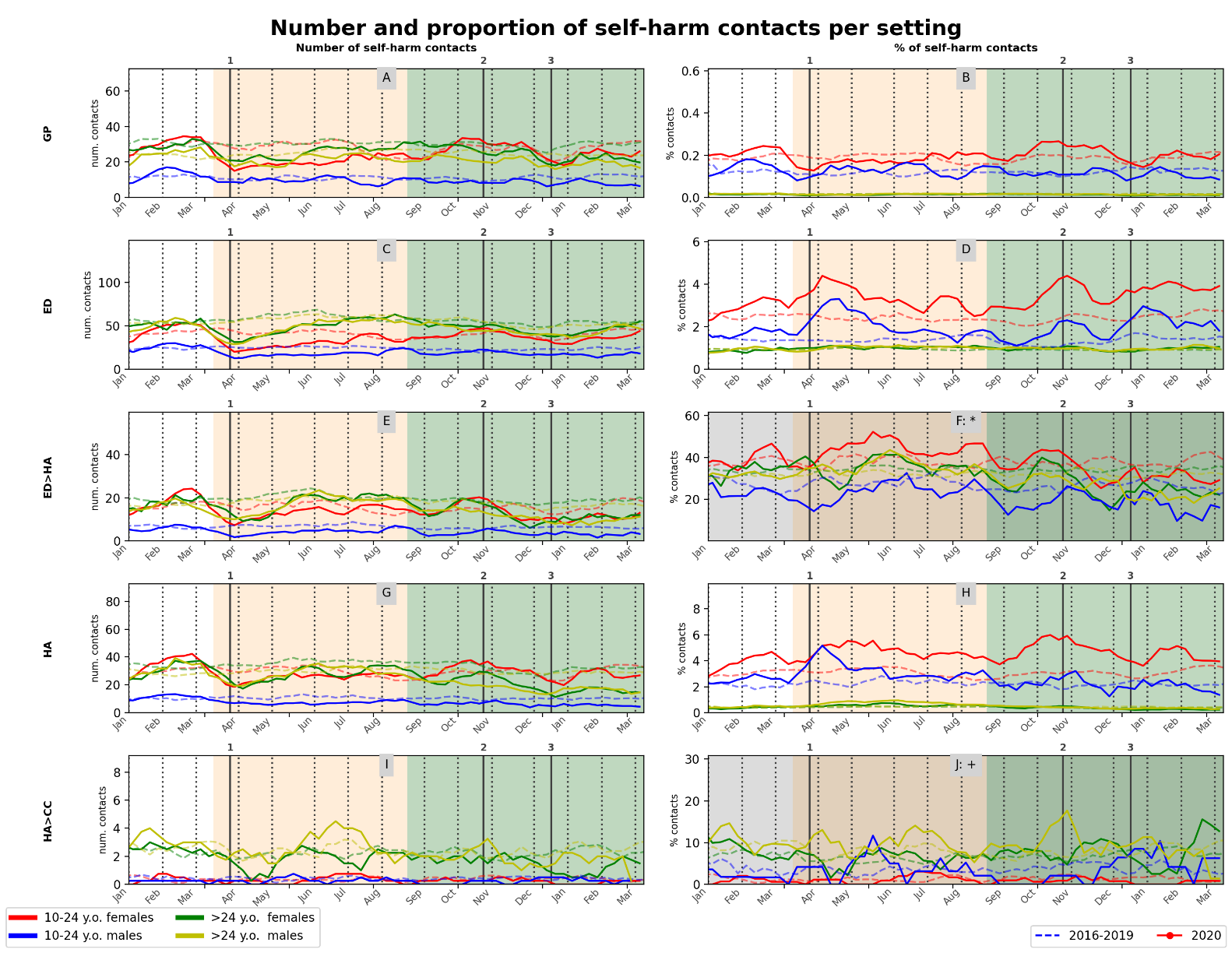

**Supplementary Figure 4.** Count (left) and proportion (right) of weekly self-harm contacts in each setting stratified by sex-age groups – (A, B) GP, (C, D) ED, (E, F*) ED followed by hospital admissions (ED>HA), (G, H) hospital admissions (HA) and (I, J+) hospital admissions with a transfer to critical care (HA>CC). Solid red lines are 4-weeks rolling average of the weekly measurements for 2020. Blue dashed lines and shaded areas are average and min-max respectively over the previous 4 years, 2016-2019. Changes in background shades correspond to before COVID-19, Wave 1 and Wave 2 periods respectively. Vertical lines are start stay-at-home measures during Wave 1 (1) and start of firebreak (2) and of stay-at-home (3) measures during Wave 2, in 2020. Darker panels show proportion of ED presentation with self-harm that resulted in a hospital admission (F*) and of hospital admissions with self-harm that resulted in a transfer to critical care (J+).

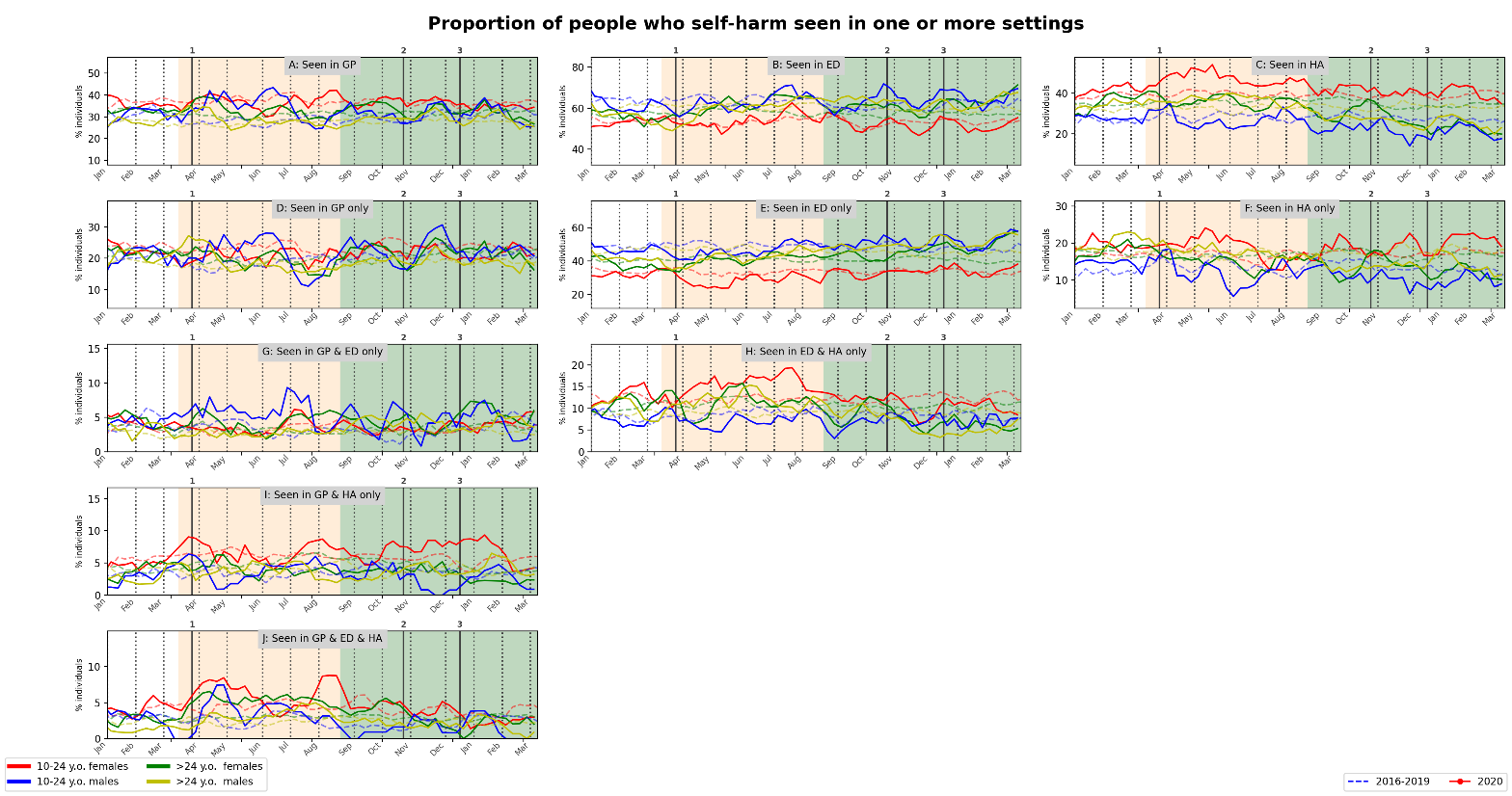

**Supplementary Figure 5.** Weekly proportion of individuals with self-harm contacts seen in GP, ED and/or hospital admissions (HA) stratified by sex-age groups. Solid red lines are 4-weeks rolling average of the weekly measurements for 2020. Blue dashed lines and shaded areas are average and min-max over the previous 4 years, 2016-2019. Panels A to C show overlapping sets. Panels D to J show non-overlapping sets. Changes in background shades correspond to before COVID-19, Wave 1 and Wave 2 periods respectively. Vertical lines are start stay-at-home measures during Wave 1 (1) and start of firebreak (2) and of stay-at-home (3) measures during Wave 2, in 2020.

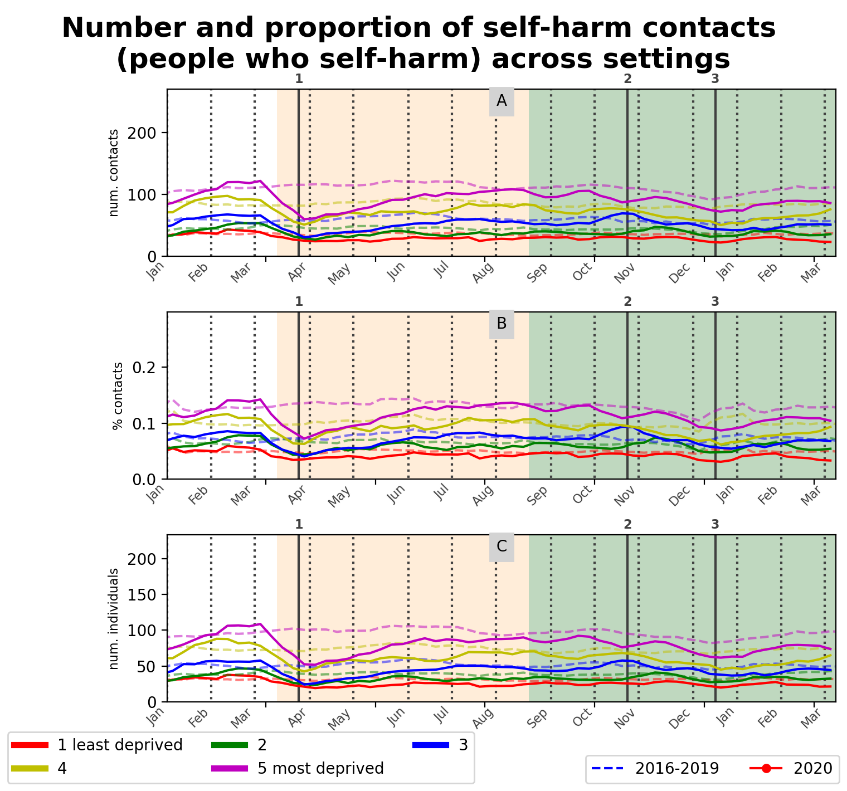

**Supplementary Figure 6**: (A) Count and (C) proportion of weekly self-harm contacts in any setting (GP, ED or hospital admissions) stratified by sex-age groups. (C) Weekly count of individuals with a self-harm contact in any setting stratified by WIMD deprivation quintile. Solid red lines are 4-weeks rolling average of the weekly measurements for 2020. Blue dashed lines are averages over the previous 4 years, 2016-2019. Changes in background shades correspond to before COVID-19, Wave 1 and Wave 2 periods respectively. Vertical lines are start stay-at-home measures during Wave 1 (1) and start of firebreak (2) and of stay-at-home (3) measures during Wave 2, in 2020.

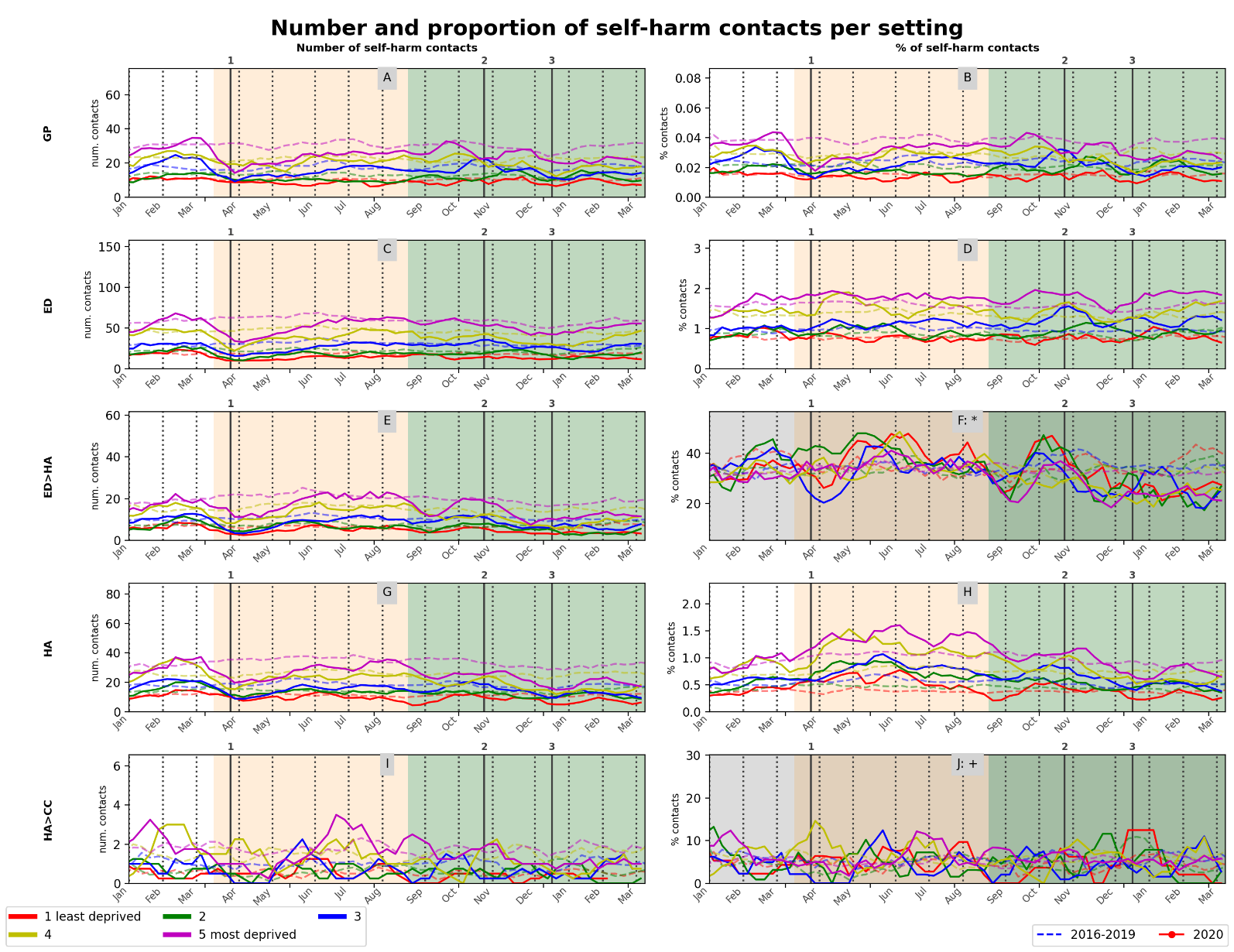

**Supplementary Figure 7.** Count (left) and proportion (right) of weekly self-harm contacts in each setting stratified by WIMD deprivation quintiles – (A, B) GP, (C, D) ED, (E, F*) ED followed by hospital admissions (ED>HA), (G, H) hospital admissions (HA) and (I, J+) hospital admissions with a transfer to critical care (HA>CC). Solid red lines are 4-weeks rolling average of the weekly measurements for 2020. Blue dashed lines and shaded areas are average and min-max respectively over the previous 4 years, 2016-2019. Changes in background shades correspond to before COVID-19, Wave 1 and Wave 2 periods respectively. Vertical lines are start stay-at-home measures during Wave 1 (1) and start of firebreak (2) and of stay-at-home (3) measures during Wave 2, in 2020. Darker panels show proportion of ED presentation with self-harm that resulted in a hospital admission (F*) and of hospital admissions with self-harm that resulted in a transfer to critical care (J+).

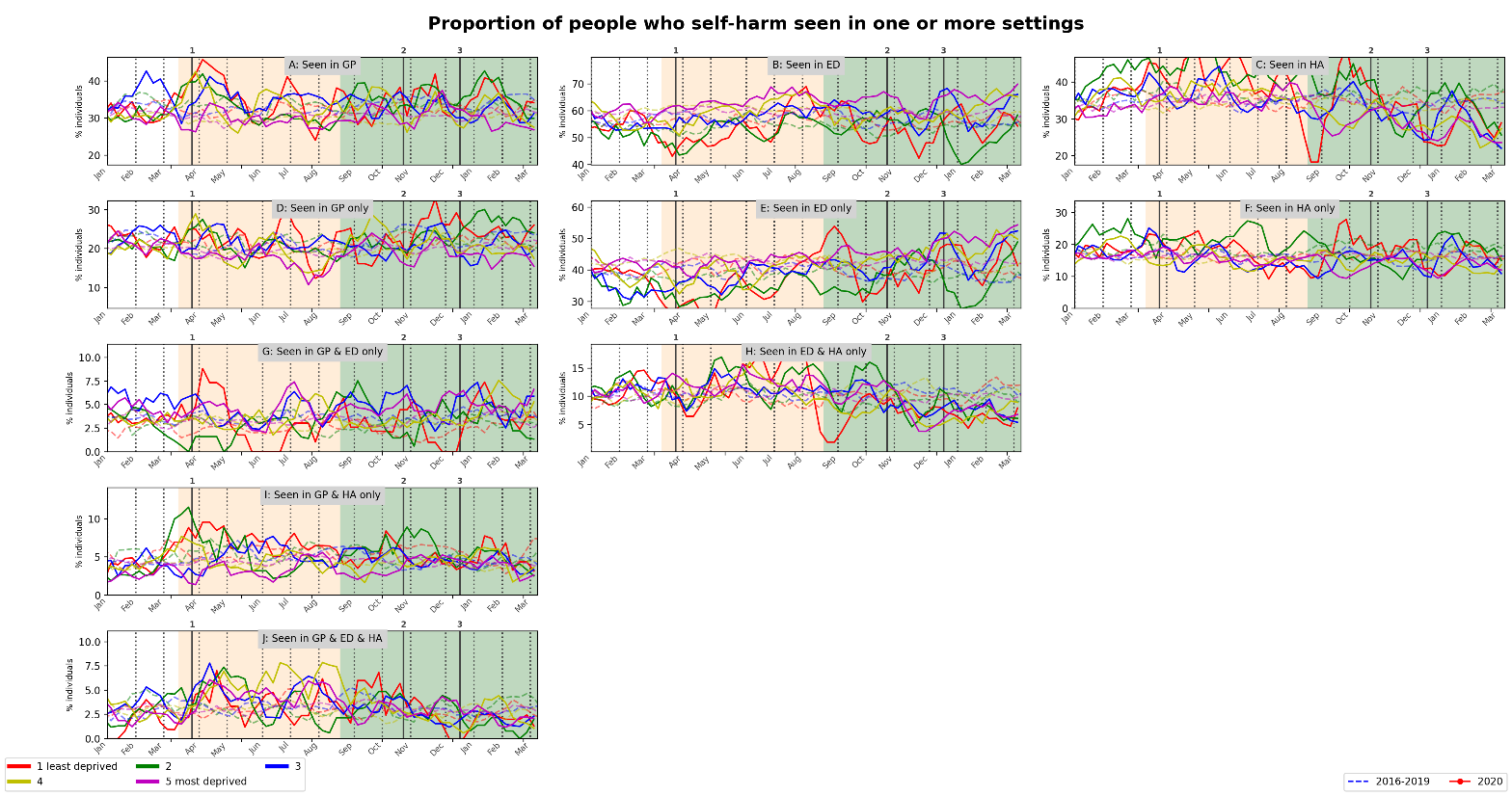

**Supplementary Figure 8.** Weekly proportion of individuals with self-harm contacts seen in GP, ED and/or hospital admissions (HA) stratified by WIMD deprivation quintiles. Solid red lines are 4-weeks rolling average of the weekly measurements for 2020. Blue dashed lines and shaded areas are average and min-max over the previous 4 years, 2016-2019. Panels A to C show overlapping sets. Panels D to J show non-overlapping sets. Changes in background shades correspond to before COVID-19, Wave 1 and Wave 2 periods respectively. Vertical lines are start stay-at-home measures during Wave 1 (1) and start of firebreak (2) and of stay-at-home (3) measures during Wave 2, in 2020.

| **A** | 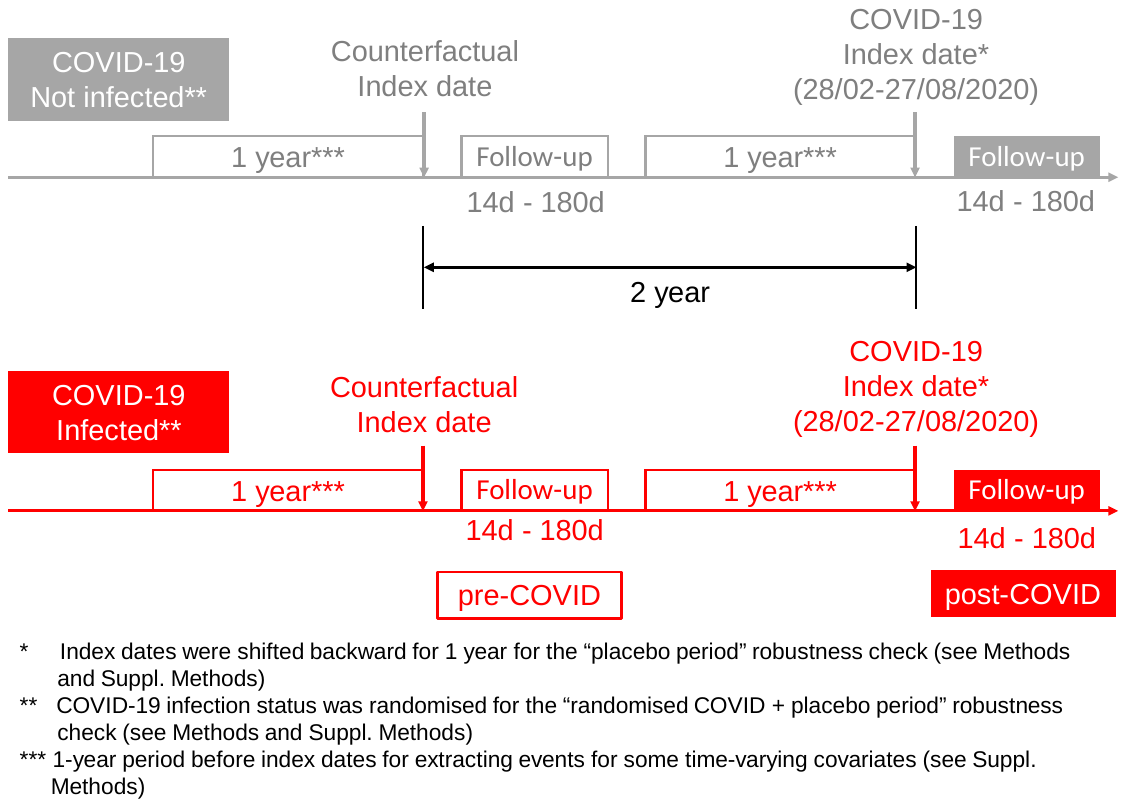 |
| --- | --- |
| **B** | 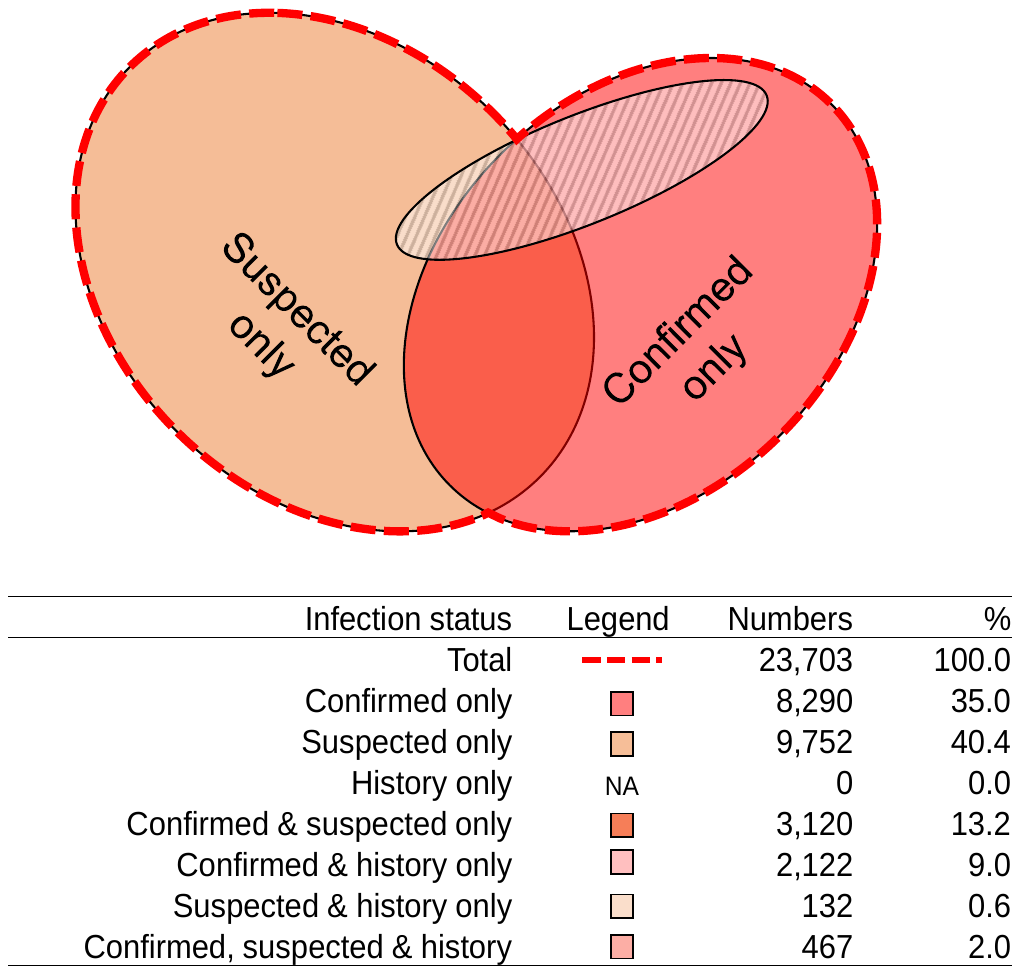 |

**Supplementary Figure 9**. (**A**) Study timeline for analysing self-harm following COVID-19 infection using difference-in-difference (DiD) approach. (**B**) Breakdown of the number of different types of COVID-19 infection events (confirmed, suspected infection or history of infection) during COVID-19 ascertainment period (28/02/2020-27/08/2020) based on the infection ascertainment criteria: ever having active confirmed or suspected infection events within ascertainment period (see Methods).

|  | Actual experiment (solid lines)  Placebo period (dotted lines) | | Actual experiment (solid lines)  Placebo period + randomised COVID (dotted lines) | |
| --- | --- | --- | --- | --- |
| Main analysis | **A** | 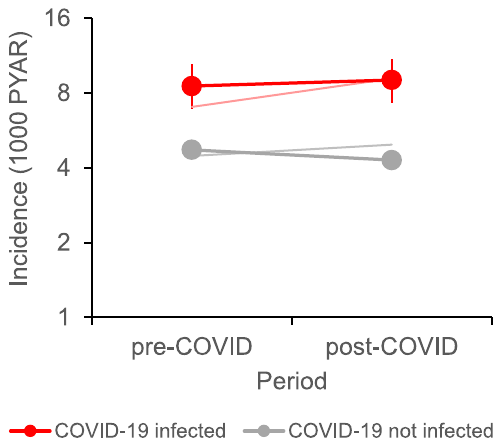 | **B** | 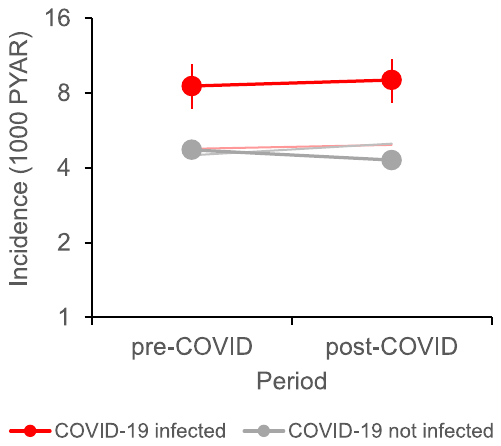 |
| Sensitivity analysis | **C** | 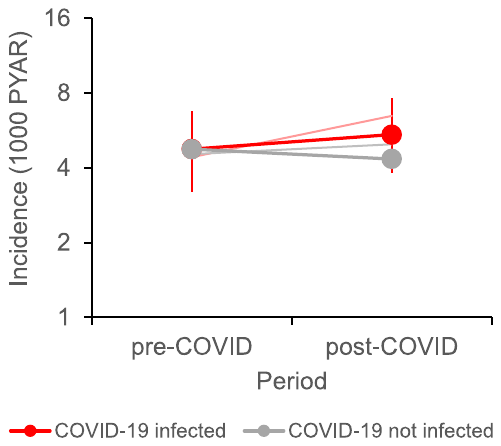 | **D** | 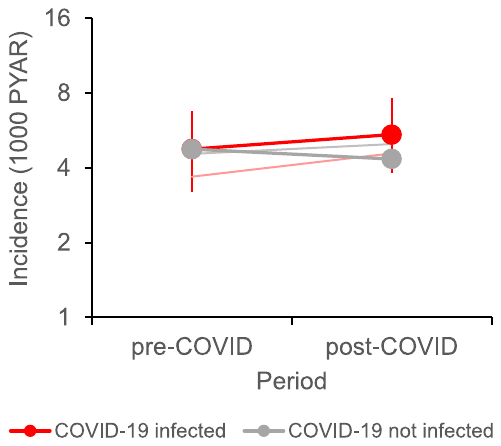 |

**Supplementary Figure 10**. Observed incidence of self-harm presentations during the pre- and post- COVID follow-up periods for the main (**A**-**B**) and sensitivity analyses (**C**-**D**). Dotted lines: observed incidences from the two robustness checks (see Methods for details). Error bars: 95% CIs.

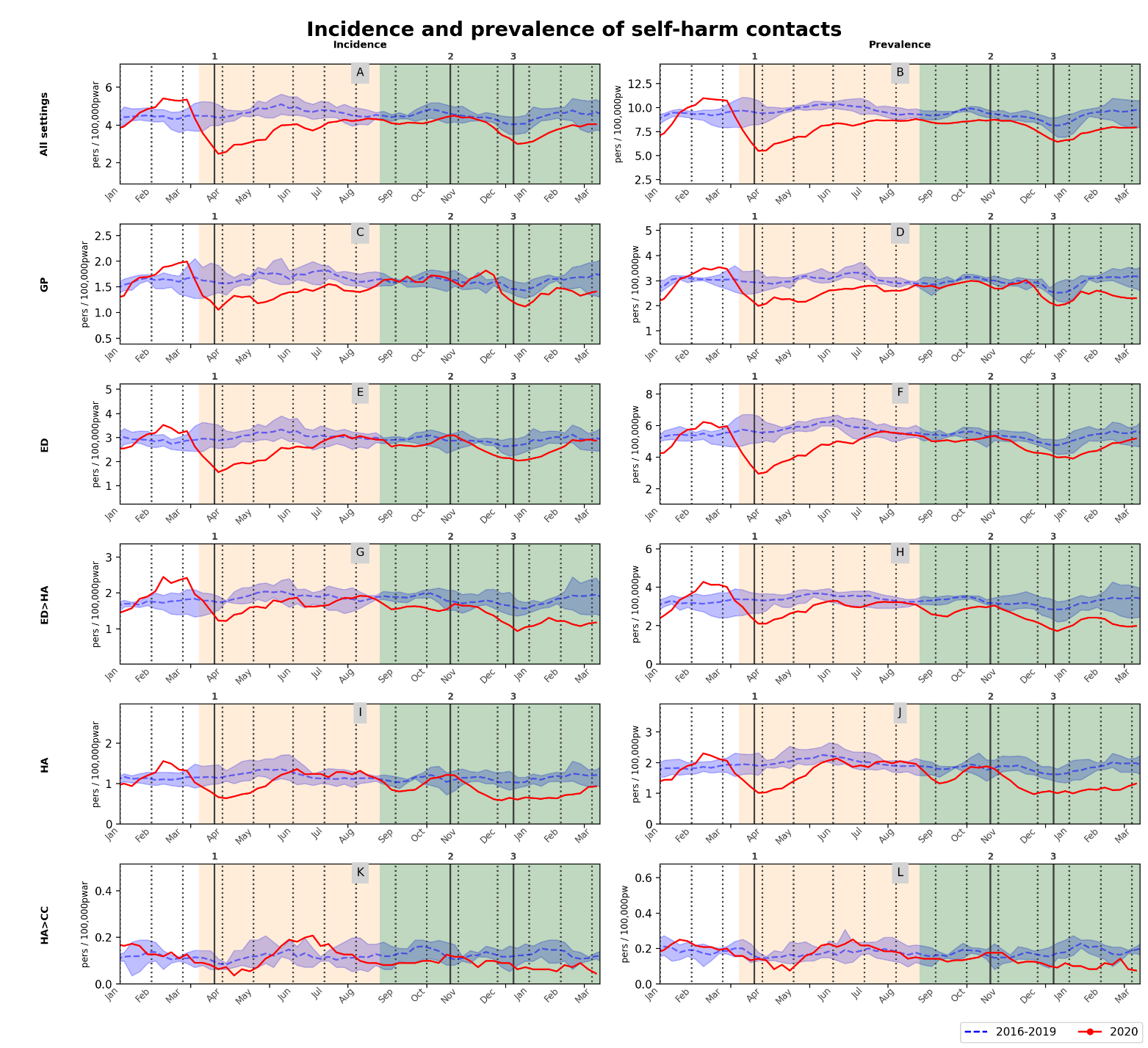

**Supplementary Figure 11.** Weekly incidence (left) and prevalence (right) of self-harm contacts (A, B) across all settings, (C, D) GP, (E, F) ED, (G, H) ED followed by hospital admissions (ED>HA), (I, J) hospital admissions (HA) and (K, L) hospital admissions with a transfer to critical care (HA>CC). Solid red lines are 4-weeks rolling average of the weekly measurements for 2020. Blue dashed line and shaded area are average and min-max over the previous 4 years, 2016-2019. Changes in background shades correspond to before COVID-19, Wave 1 and Wave 2 periods respectively. Vertical lines are start stay-at-home measures during Wave 1 (1) and start of firebreak (2) and of stay-at-home (3) measures during Wave 2, in 2020.

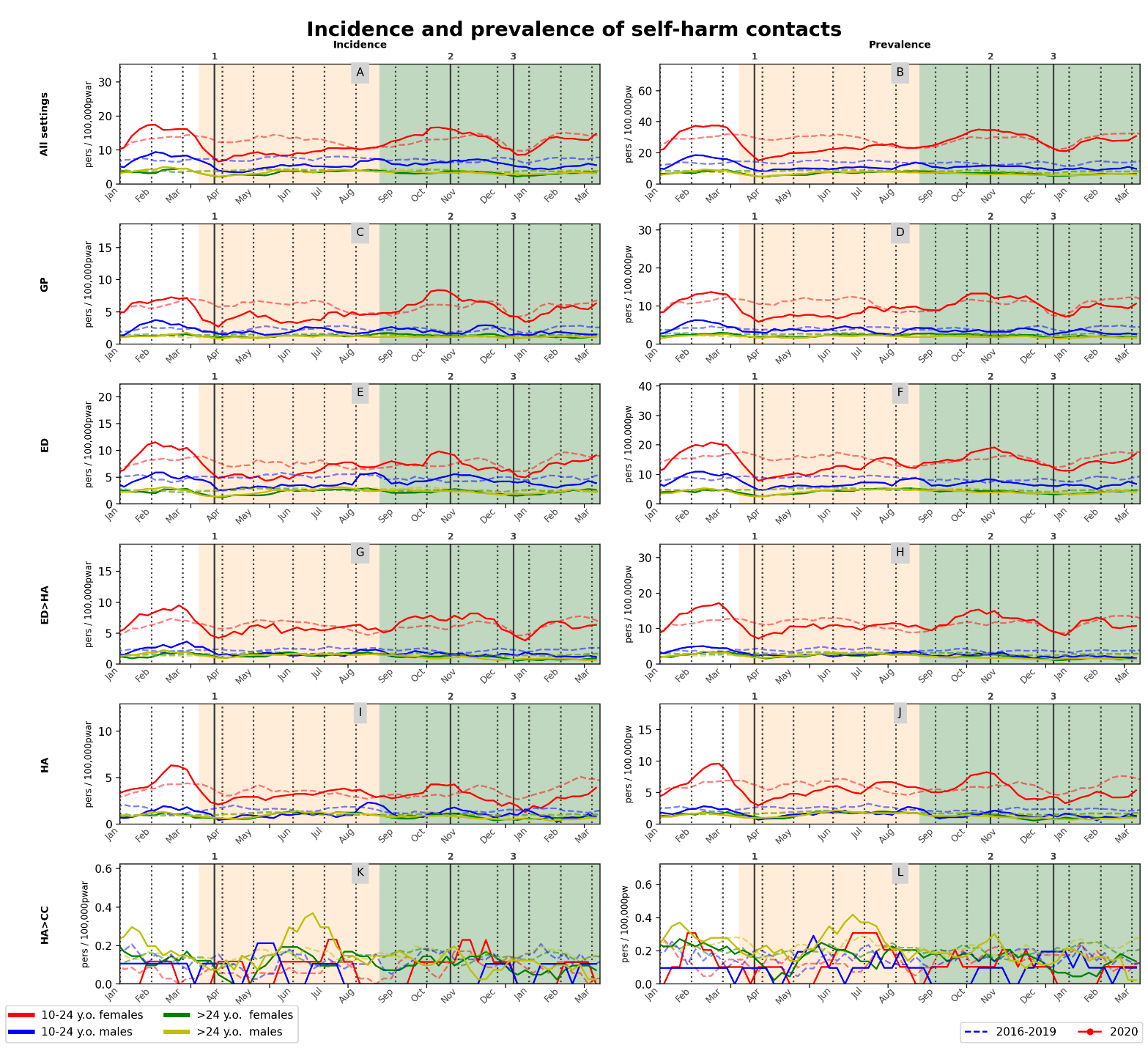

**Supplementary Figure 12.** Weekly incidence (left) and prevalence (right) of self-harm contacts (A, B) across all settings, (C, D) GP, (E, F) ED, (G, H) ED followed by hospital admissions (ED>HA), (I, J) hospital admissions (HA) and (K, L) hospital admissions with a transfer to critical care (HA>CC) stratified by sex-age groups. Solid red lines are 4-weeks rolling average of the weekly measurements for 2020. Blue dashed line and shaded area are average and min-max over the previous 4 years, 2016-2019. Changes in background shades correspond to before COVID-19, Wave 1 and Wave 2 periods respectively. Vertical lines are start stay-at-home measures during Wave 1 (1) and start of firebreak (2) and of stay-at-home (3) measures during Wave 2, in 2020.

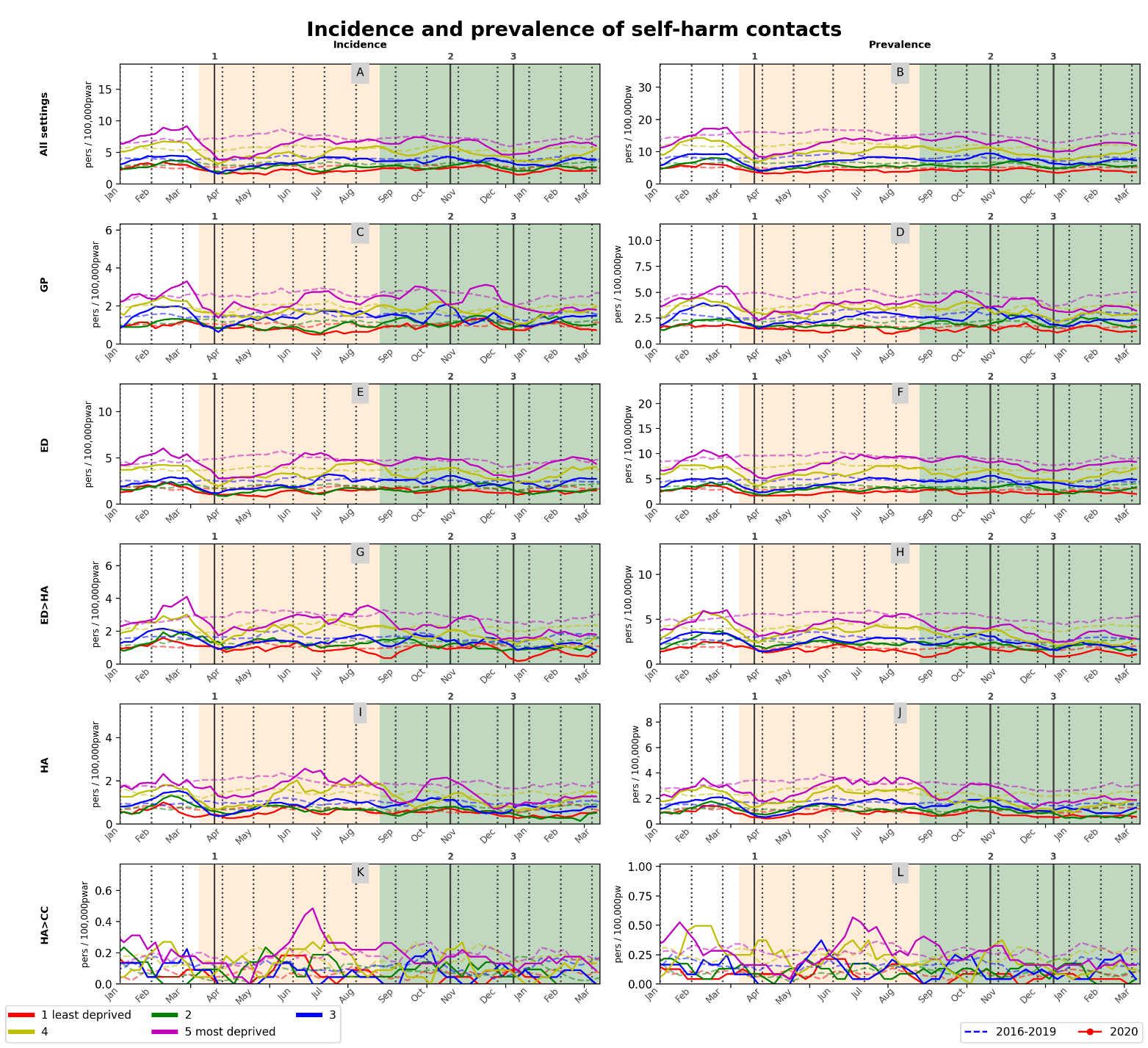

**Supplementary Figure 13.** Weekly incidence (left) and prevalence (right) of self-harm contacts (A, B) across all settings, (C, D) GP, (E, F) ED, (G, H) ED followed by hospital admissions (ED>HA), (I, J) hospital admissions (HA) and (K, L) hospital admissions with a transfer to critical care (HA>CC) stratified by WIMD deprivation quintiles. Solid red lines are 4-weeks rolling average of the weekly measurements for 2020. Blue dashed line and shaded area are average and min-max over the previous 4 years, 2016-2019. Changes in background shades correspond to before COVID-19, Wave 1 and Wave 2 periods respectively. Vertical lines are start stay-at-home measures during Wave 1 (1) and start of firebreak (2) and of stay-at-home (3) measures during Wave 2, in 2020.

**Supplementary Table 1:** Data sources used in this analysis.

| **Database** | **Description** | **Coverage at time of data extraction (28/04/2021)** |
| --- | --- | --- |
| Welsh Demographic Service | An administrative register of all individuals in Wales that use NHS services, containing anonymised demographics and GP practice registration history with anonymised residential data |  |
| Office for National Statistics – Mortality register^§^ | Death register of all deaths and causes in Wales, coded using International Classification of Diseases (ICD), version 10 codes, derived from information collected at registration of death. Daily and monthly extracts available. | Up to 28/3/2021 |
| Consolidated Deaths Data Source^§^ | Combination of death records from the Wales Demographic Service Dataset, Master Patient Index and ONS Deaths. | Up to 18/4/2021 |
| Welsh Longitudinal General Practice | Primary care records with diagnoses, symptoms, investigations, prescribed medication, referrals, coded hospital contacts, and test results coded using Read Codes v2 | 77% (333/432) of all general practices in Wales up to 21/3/2021 |
| Emergency Department Data Set^†^ | Administrative and clinical information (general reason for attendance and attendance group to identify types of contacts) for all NHS Wales Accident and Emergency department attendances. | Up to 25/4/2021 |
| Patient Episode Database for Wales^‡^ | Clinical information (specialty and diagnoses) of all NHS Wales hospital admissions (inpatient and day cases) – diagnostic information coded using ICD-10 codes. | Up to 25/4/2021 |
| Welsh Laboratory Information Systems Database | Clinical system that stores, records and exchanges both COVID-19 antigen/swab tests (pillar 1 and partial pillar 2) and serology/antibodies tests (pillar 3) results from NHS Wales laboratories. | Form 05/02/2020 to 08/06/2021 |
| Care Homes Data | Contains geographic information data about care homes. | Up to 03/2021 |
| COVID-19 Shielded People list | List of high-risk individuals advised to self-isolate during COVID-19 pandemic. | Up to 03/2021 |
| Office of National Statistics Census 2011 (Wales) | Data of the latest census held on March 2011 for all people and households. | 03/2011 |
| * Data have full Wales coverage throughout the study period unless otherwise shown.  § User guide to mortality statistics - Office for National Statistics. [Online]. Available: https://www.ons.gov.uk/peoplepopulationandcommunity/birthsdeathsandmarriages/deaths/methodologies/userguidetomortalitystatisticsjuly2017  † Digital Health and Care Wales (previously NHS Wales Informatics Service), Emergency Department Data Set structure. [Online]. Available: http://www.datadictionary.wales.nhs.uk/#!WordDocuments/datasetstructure4.htm  ‡ Digital Health and Care Wales (previously NHS Wales Informatics Service), Patient Episode Database for Wales Data Set structure. [Online]. Available: http://www.datadictionary.wales.nhs.uk/#!WordDocuments/datasetstructure.htm | | |

**Supplementary Table 2**. Summary of RORs and RRRs comparing weekly change in self-harm contacts between reference and target COVID-19 periods to the respective changes in previous years across settings (Any), per setting (GP, ED and hospital admissions; HA) and for ED presentations with subsequent hospitalisation (ED to HA).

|  |  |  | |  | | Year as |  |  | | | | |  |  |
| --- | --- | --- | --- | --- | --- | --- | --- | --- | --- | --- | --- | --- | --- | --- |
| Setting | Outcome | Reference period^a^ | | Target period^a^ | | counterfactual | RRR/ROR^b^ | 95% CI | | | | | *p*-value | *p*-value* |
| Any | numbers | week 1-10 | 30/12/2019 | week 12-14 | 16/03/2020 | 2016-2017 | 0.576 | ( | 0.510 | , | 0.652 | ) | <0.001 | **<0.001** |
|  |  |  | to |  | to | 2017-2018 | 0.576 | ( | 0.509 | , | 0.652 | ) | <0.001 | **<0.001** |
|  |  |  | 08/03/2020 |  | 05/04/2020 | 2018-2019 | 0.600 | ( | 0.529 | , | 0.680 | ) | <0.001 | **<0.001** |
| Any | numbers | week 1-10 | 30/12/2019 | week 30-33 | 20/07/2020 | 2016-2017 | 0.979 | ( | 0.886 | , | 1.082 | ) | 0.683 | >0.999 |
|  |  |  | to |  | to | 2017-2018 | 0.914 | ( | 0.828 | , | 1.009 | ) | 0.074 | 0.223 |
|  |  |  | 08/03/2020 |  | 16/08/2020 | 2018-2019 | 0.788 | ( | 0.714 | , | 0.869 | ) | <0.001 | **<0.001** |
| Any | numbers | week 1-10 | 30/12/2019 | week 50-53 | 07/12/2020 | 2016-2017 | 0.730 | ( | 0.654 | , | 0.816 | ) | <0.001 | **<0.001** |
|  |  |  | to |  | to | 2017-2018 | 0.649 | ( | 0.582 | , | 0.723 | ) | <0.001 | **<0.001** |
|  |  |  | 08/03/2020 |  | 03/01/2021 | 2018-2019 | 0.622 | ( | 0.557 | , | 0.694 | ) | <0.001 | **<0.001** |
| Any | proportion | week 1-10 | 30/12/2019 | week 12-14 | 16/03/2020 | 2016-2017 | 0.449 | ( | 0.446 | , | 0.559 | ) | <0.001 | **<0.001** |
|  |  |  | to |  | to | 2017-2018 | 0.553 | ( | 0.494 | , | 0.620 | ) | <0.001 | **<0.001** |
|  |  |  | 08/03/2020 |  | 05/04/2020 | 2018-2019 | 0.538 | ( | 0.478 | , | 0.605 | ) | <0.001 | **<0.001** |
| Any | proportion | week 1-10 | 30/12/2019 | week 30-33 | 20/07/2020 | 2016-2017 | 1.025 | ( | 0.936 | , | 1.124 | ) | 0.590 | >0.999 |
|  |  |  | to |  | to | 2017-2018 | 0.925 | ( | 0.879 | , | 1.054 | ) | 0.407 | >0.999 |
|  |  |  | 08/03/2020 |  | 16/08/2020 | 2018-2019 | 0.829 | ( | 0.757 | , | 0.908 | ) | <0.001 | **<0.001** |
| Any | proportion | week 1-10 | 30/12/2019 | week 50-53 | 07/12/2020 | 2016-2017 | 0.763 | ( | 0.692 | , | 0.842 | ) | <0.001 | **<0.001** |
|  |  |  | to |  | to | 2017-2018 | 0.734 | ( | 0.666 | , | 0.810 | ) | <0.001 | **<0.001** |
|  |  |  | 08/03/2020 |  | 03/01/2021 | 2018-2019 | 0.681 | ( | 0.617 | , | 0.753 | ) | <0.001 | **<0.001** |
| GP | numbers | week 1-10 | 30/12/2019 | week 12-14 | 16/03/2020 | 2016-2017 | 0.665 | ( | 0.535 | , | 0.826 | ) | <0.001 | **<0.001** |
|  |  |  | to |  | to | 2017-2018 | 0.643 | ( | 0.518 | , | 0.799 | ) | <0.001 | **<0.001** |
|  |  |  | 08/03/2020 |  | 05/04/2020 | 2018-2019 | 0.748 | ( | 0.598 | , | 0.935 | ) | 0.011 | **0.032** |
| GP | numbers | week 1-10 | 30/12/2019 | week 30-33 | 20/07/2020 | 2016-2017 | 0.944 | ( | 0.789 | , | 1.130 | ) | 0.529 | >0.999 |
|  |  |  | to |  | to | 2017-2018 | 0.938 | ( | 0.783 | , | 1.123 | ) | 0.484 | >0.999 |
|  |  |  | 08/03/2020 |  | 16/08/2020 | 2018-2019 | 0.808 | ( | 0.677 | , | 0.964 | ) | 0.018 | 0.054 |
| GP | numbers | week 1-10 | 30/12/2019 | week 50-53 | 07/12/2020 | 2016-2017 | 0.731 | ( | 0.599 | , | 0.893 | ) | 0.002 | **0.006** |
|  |  |  | to |  | to | 2017-2018 | 0.612 | ( | 0.504 | , | 0.743 | ) | <0.001 | **<0.001** |
|  |  |  | 08/03/2020 |  | 03/01/2021 | 2018-2019 | 0.698 | ( | 0.571 | , | 0.852 | ) | <0.001 | **0.001** |
| GP | proportion | week 1-10 | 30/12/2019 | week 12-26 | 16/03/2020 | 2016-2017 | 0.731 | ( | 0.648 | , | 0.823 | ) | <0.001 | **<0.001** |
|  |  |  | to |  | to | 2017-2018 | 0.742 | ( | 0.658 | , | 0.836 | ) | <0.001 | **<0.001** |
|  |  |  | 08/03/2020 |  | 28/06/2020 | 2018-2019 | 0.752 | ( | 0.666 | , | 0.848 | ) | <0.001 | **<0.001** |
| GP | proportion | week 1-10 | 30/12/2019 | week 30-33 | 20/07/2020 | 2016-2017 | 0.944 | ( | 0.789 | , | 1.130 | ) | 0.529 | >0.999 |
|  |  |  | to |  | to | 2017-2018 | 0.938 | ( | 0.783 | , | 1.123 | ) | 0.484 | >0.999 |
|  |  |  | 08/03/2020 |  | 16/08/2020 | 2018-2019 | 0.808 | ( | 0.677 | , | 0.964 | ) | 0.018 | 0.054 |
| GP | proportion | week 1-10 | 30/12/2019 | week 50-53 | 07/12/2020 | 2016-2017 | 0.684 | ( | 0.566 | , | 0.825 | ) | <0.001 | **<0.001** |
|  |  |  | to |  | to | 2017-2018 | 0.656 | ( | 0.544 | , | 0.791 | ) | <0.001 | **<0.001** |
|  |  |  | 08/03/2020 |  | 03/01/2021 | 2018-2019 | 0.695 | ( | 0.574 | , | 0.842 | ) | <0.001 | **<0.001** |
| ED | numbers | week 1-10 | 30/12/2019 | week 12-14 | 16/03/2020 | 2016-2017 | 0.528 | ( | 0.458 | , | 0.608 | ) | <0.001 | **<0.001** |
|  |  |  | to |  | to | 2017-2018 | 0.568 | ( | 0.493 | , | 0.656 | ) | <0.001 | **<0.001** |
|  |  |  | 08/03/2020 |  | 05/04/2020 | 2018-2019 | 0.576 | ( | 0.498 | , | 0.666 | ) | <0.001 | **<0.001** |
| ED | proportion | week 1-10 | 30/12/2019 | week 16-17 | 13/04/2020 | 2016-2017 | 1.270 | ( | 1.070 | , | 1.508 | ) | 0.006 | **0.019** |
|  |  |  | to |  | to | 2017-2018 | 1.426 | ( | 1.198 | , | 1.698 | ) | <0.001 | **<0.001** |
|  |  |  | 08/03/2020 |  | 26/04/2020 | 2018-2019 | 1.195 | ( | 1.006 | , | 1.420 | ) | 0.043 | 0.129 |
| ED | numbers | week 1-10 | 30/12/2019 | week 30-33 | 20/07/2020 | 2016-2017 | 1.104 | ( | 0.988 | , | 1.234 | ) | 0.081 | 0.244 |
|  |  |  | to |  | to | 2017-2018 | 0.978 | ( | 0.877 | , | 1.091 | ) | 0.692 | >0.999 |
|  |  |  | 08/03/2020 |  | 16/08/2020 | 2018-2019 | 0.891 | ( | 0.799 | , | 0.994 | ) | 0.038 | 0.115 |
| ED | proportion | week 1-10 | 30/12/2019 | week 41-45 | 05/10/2020 | 2016-2017 | 1.305 | ( | 1.161 | , | 1.467 | ) | <0.001 | **<0.001** |
|  |  |  | to |  | to | 2017-2018 | 1.261 | ( | 1.123 | , | 1.415 | ) | <0.001 | **<0.001** |
|  |  |  | 08/03/2020 |  | 08/11/2020 | 2018-2019 | 1.064 | ( | 0.947 | , | 1.194 | ) | 0.295 | 0.886 |
| ED | numbers | week 1-10 | 30/12/2019 | week 50-53 | 07/12/2020 | 2016-2017 | 0.703 | ( | 0.620 | , | 0.797 | ) | <0.001 | **<0.001** |
|  |  |  | to |  | to | 2017-2018 | 0.664 | ( | 0.586 | , | 0.752 | ) | <0.001 | **<0.001** |
|  |  |  | 08/03/2020 |  | 03/01/2021 | 2018-2019 | 0.635 | ( | 0.560 | , | 0.720 | ) | <0.001 | **<0.001** |
| ED | proportion | week 1-10 | 30/12/2019 | week 53-58 | 28/12/2020 | 2016-2017 | 1.092 | ( | 0.981 | , | 1.216 | ) | 0.107 | 0.321 |
|  |  |  | to |  | to | 2017-2018 | 1.158 | ( | 1.039 | , | 1.290 | ) | 0.008 | **0.024** |
|  |  |  | 08/03/2020 |  | 07/02/2021 | 2018-2019 | 0.932 | ( | 0.837 | , | 1.037 | ) | 0.196 | 0.587 |
| ED to HA | numbers | week 1-10 | 30/12/2019 | week 12-14 | 16/03/2020 | 2016-2017 | 0.515 | ( | 0.390 | , | 0.680 | ) | <0.001 | **<0.001** |
|  |  |  | to |  | to | 2017-2018 | 0.580 | ( | 0.439 | , | 0.767 | ) | <0.001 | **<0.001** |
|  |  |  | 08/03/2020 |  | 05/04/2020 | 2018-2019 | 0.669 | ( | 0.502 | , | 0.892 | ) | 0.006 | **0.018** |
| ED to HA | numbers | week 1-10 | 30/12/2019 | week 30-33 | 20/07/2020 | 2016-2017 | 1.091 | ( | 0.875 | , | 1.360 | ) | 0.437 | >0.999 |
|  |  |  | to |  | to | 2017-2018 | 0.986 | ( | 0.797 | , | 1.221 | ) | 0.901 | >0.999 |
|  |  |  | 08/03/2020 |  | 16/08/2020 | 2018-2019 | 0.965 | ( | 0.779 | , | 1.196 | ) | 0.746 | >0.999 |
| ED to HA | numbers | week 1-10 | 30/12/2019 | week 50-53 | 07/12/2020 | 2016-2017 | 0.527 | ( | 0.402 | , | 0.691 | ) | <0.001 | **<0.001** |
|  |  |  | to |  | to | 2017-2018 | 0.503 | ( | 0.385 | , | 0.656 | ) | <0.001 | **<0.001** |
|  |  |  | 08/03/2020 |  | 03/01/2021 | 2018-2019 | 0.484 | ( | 0.371 | , | 0.632 | ) | <0.001 | **<0.001** |
| HA | numbers | week 1-10 | 30/12/2019 | week 12-14 | 16/03/2020 | 2016-2017 | 0.561 | ( | 0.459 | , | 0.685 | ) | <0.001 | **<0.001** |
|  |  |  | to |  | to | 2017-2018 | 0.583 | ( | 0.477 | , | 0.712 | ) | <0.001 | **<0.001** |
|  |  |  | 08/03/2020 |  | 05/04/2020 | 2018-2019 | 0.630 | ( | 0.510 | , | 0.779 | ) | <0.001 | **<0.001** |
| HA | proportion | week 1-10 | 30/12/2019 | week 15-33 | 06/04/2020 | 2016-2017 | 1.292 | ( | 1.162 | , | 1.437 | ) | <0.001 | **<0.001** |
|  |  |  | to |  | to | 2017-2018 | 1.297 | ( | 1.166 | , | 1.442 | ) | <0.001 | **<0.001** |
|  |  |  | 08/03/2020 |  | 16/08/2020 | 2018-2019 | 1.128 | ( | 1.010 | , | 1.259 | ) | 0.032 | 0.096 |
| HA | numbers | week 1-10 | 30/12/2019 | week 30-33 | 20/07/2020 | 2016-2017 | 0.931 | ( | 0.789 | , | 1.099 | ) | 0.398 | >0.999 |
|  |  |  | to |  | to | 2017-2018 | 0.945 | ( | 0.802 | , | 1.115 | ) | 0.504 | >0.999 |
|  |  |  | 08/03/2020 |  | 16/08/2020 | 2018-2019 | 0.753 | ( | 0.638 | , | 0.889 | ) | <0.001 | **0.003** |
| HA | proportion | week 1-10 | 30/12/2019 | week 41-44 | 05/10/2020 | 2016-2017 | 1.201 | ( | 1.018 | , | 1.418 | ) | 0.030 | 0.091 |
|  |  |  | to |  | to | 2017-2018 | 1.256 | ( | 1.063 | , | 1.483 | ) | 0.007 | **0.022** |
|  |  |  | 08/03/2020 |  | 01/11/2020 | 2018-2019 | 0.999 | ( | 0.846 | , | 1.181 | ) | 0.993 | >0.999 |
| HA | numbers | week 1-10 | 30/12/2019 | week 30-33 | 20/07/2020 | 2016-2017 | 0.522 | ( | 0.426 | , | 0.640 | ) | <0.001 | **<0.001** |
|  |  |  | to |  | to | 2017-2018 | 0.453 | ( | 0.371 | , | 0.553 | ) | <0.001 | **<0.001** |
|  |  |  | 08/03/2020 |  | 16/08/2020 | 2018-2019 | 0.370 | ( | 0.303 | , | 0.453 | ) | <0.001 | **<0.001** |
| Any | numbers | week 36-44 | 31/08/2020 | week 50-53 | 07/12/2020 | 2016-2017 | 0.803 | ( | 0.717 | , | 0.899 | ) | <0.001 | **<0.001** |
|  |  |  | to |  | to | 2017-2018 | 0.727 | ( | 0.651 | , | 0.813 | ) | <0.001 | **<0.001** |
|  |  |  | 01/11/2020 |  | 03/01/2021 | 2018-2019 | 0.768 | ( | 0.687 | , | 0.859 | ) | <0.001 | **<0.001** |
| GP | numbers | week 36-44 | 31/08/2020 | week 50-53 | 07/12/2020 | 2016-2017 | 0.777 | ( | 0.634 | , | 0.952 | ) | 0.015 | **0.045** |
|  |  |  | to |  | to | 2017-2018 | 0.643 | ( | 0.528 | , | 0.784 | ) | <0.001 | **<0.001** |
|  |  |  | 01/11/2020 |  | 03/01/2021 | 2018-2019 | 0.732 | ( | 0.597 | , | 0.897 | ) | 0.003 | **0.008** |
| ED | numbers | week 36-44 | 31/08/2020 | week 50-53 | 07/12/2020 | 2016-2017 | 0.711 | ( | 0.625 | , | 0.808 | ) | <0.001 | **<0.001** |
|  |  |  | to |  | to | 2017-2018 | 0.731 | ( | 0.644 | , | 0.830 | ) | <0.001 | **<0.001** |
|  |  |  | 01/11/2020 |  | 03/01/2021 | 2018-2019 | 0.756 | ( | 0.666 | , | 0.859 | ) | <0.001 | **<0.001** |
| ED to HA | numbers | week 36-44 | 31/08/2020 | week 50-53 | 07/12/2020 | 2016-2017 | 0.598 | ( | 0.453 | , | 0.788 | ) | <0.001 | **<0.001** |
|  |  |  | to |  | to | 2017-2018 | 0.617 | ( | 0.471 | , | 0.809 | ) | <0.001 | **0.001** |
|  |  |  | 01/11/2020 |  | 03/01/2021 | 2018-2019 | 0.613 | ( | 0.468 | , | 0.803 | ) | <0.001 | **0.001** |
| HA | numbers | week 36-44 | 31/08/2020 | week 50-53 | 07/12/2020 | 2016-2017 | 0.688 | ( | 0.559 | , | 0.848 | ) | <0.001 | **0.001** |
|  |  |  | to |  | to | 2017-2018 | 0.570 | ( | 0.465 | , | 0.699 | ) | <0.001 | **<0.001** |
|  |  |  | 01/11/2020 |  | 03/01/2021 | 2018-2019 | 0.586 | ( | 0.478 | , | 0.717 | ) | <0.001 | **<0.001** |
| * Bonferroni corrected | | | | | | | | | | | | | | |
| ^a^ Period > 1 week represented by the mean of the model coefficients within the period | | | | | | | | | | | | | | |
| ^b^ RRR-ratio of rate ratios for prevalence/incidence outcomes; ROR-ratio of odds ratio for proportion outcomes | | | | | | | | | | | | | | |

**Supplementary Table 3**. Results of linear trends of weekly number and proportion of self-harm contacts and weekly proportion of people with self-harm contacts for GP, ED and hospital admissions (HA).

| Setting | Outcome  (units) | Fitted period | Slope per month^a^  (95% CI) | p-value  of slope | Adjusted p-value  of slope^b^ | p-value for  slope comparisons^c^ |
| --- | --- | --- | --- | --- | --- | --- |
| ED | Proportion of contacts (%) | 6/2020-3/2021 | -1.899 (-2.264, -1.530) | <0.001 | **<0.001** | reference |
|  |  | 6/2016-3/2017 | -0.158 (-0.479, 0.162) | 0.328 | >0.999 | **0.001** |
|  |  | 6/2017-3/2018 | 0.109 (-0.163 ,0.380) | 0.427 | >0.999 | **<0.001** |
|  |  | 6/2018-3/2019 | 0.159 (-0.121 ,0.440) | 0.261 | >0.999 | **<0.001** |
|  |  | 6/2019-3/2020 | 0.148 (-0.176 ,0.471) | 0.366 | >0.999 | **<0.001** |
| HA | Number of contacts (#) | 6/2020-3/2021 | -4.702 (-6.025, -3.380) | <0.001 | **<0.001** | reference |
|  |  | 6/2016-3/2017 | -2.066 (-3.466, -0.660) | 0.005 | **0.025** | **0.010** |
|  |  | 6/2017-3/2018 | -4.07(-5.655, -2.480) | <0.001 | **<0.001** | 0.500 |
|  |  | 6/2018-3/2019 | -0.429 (-1.762 ,0.903) | 0.518 | >0.999 | **<0.001** |
|  |  | 6/2019-3/2020 | 1.292 (0.64 ,3.220) | 0.184 | 0.919 | **<0.001** |
| HA | Proportions of people (%) | 8/2020-3/2021 | -0.992 (-1.686, -0.297) | <0.007 | **0.034** | reference |
|  |  | 8/2016-3/2017 | -0.393 (-0.837, 0.052) | 0.081 | 0.406 | 0.112 |
|  |  | 8/2017-3/2018 | -0.649 (-1.147, -0.152) | 0.012 | 0.062 | 0.362 |
|  |  | 8/2018-3/2019 | 0.385 (-0.083, 0.853) | 0.103 | 0.514 | **<0.001** |
|  |  | 8/2019-3/2020 | 0.192 (-0.390, 0.773) | 0.505 | >0.999 | **0.002** |
| ED and HA only | Proportions of people (%) | 6/2020-12/2020 | -1.163 (-1.677, -0.649) | <0.001 | **<0.001** | reference |
|  |  | 6/2016-12/2016 | -0.046 (-0.420, 0.329) | 0.804 | >0.999 | **<0.001** |
|  |  | 6/2017-12/2017 | -0.421 (-0.779, -0.063) | 0.023 | 0.115 | **0.008** |
|  |  | 6/2018-12/2018 | 0.171 ( -0.242, 0.583) | 0.402 | >0.999 | **<0.001** |
|  |  | 6/2019-12/2019 | 0.207 ( -0.106 ,0.520) | 0.186 | 0.928 | **<0.001** |
| ED, HA and GP | Proportions of people (%) | 4/2020-end 12/2020 | -0.382 (-0.524, -0.240) | <0.001 | **<0.001** | reference |
|  |  | 4/2016-end 12/2016 | -0.125 (-0.266, 0.016) | 0.081 | 0.404 | **0.006** |
|  |  | 4/2017-end 12/2017 | 0.049 (-0.089, 0.186) | 0.476 | >0.999 | **<0.001** |
|  |  | 4/2018-end 12/2018 | -0.049 (-0.166, 0.068) | 0.402 | >0.999 | **<0.001** |
|  |  | 4/2019-end 12/2019 | 0.025 (-0.109, 0.159) | 0.705 | >0.999 | **<0.001** |
| ED only | Proportions of people (%) | 4/2020-end 2/2021 | 0.977 (0.639, 1.315) | <0.001 | **<0.001** | reference |
|  |  | 4/2016- end 2/2017 | 0.073 (-0.217, 0.362) | 0.616 | >0.999 | **<0.001** |
|  |  | 4/2017-end 2/2018 | 0.028 (-0.239, 0.295) | 0.834 | >0.999 | **<0.001** |
|  |  | 4/2018-end 2/2019 | -0.362 (-0.617, -0.107) | 0.006 | 0.032 | **<0.001** |
|  |  | 4/2019-end 2/2020 | -0.610 (-0.885, -0.335) | <0.001 | **0.002** | **<0.001** |
| HA | Male adults number of contacts (#) | 6/2020-12/2020 | -3.441 (-4.192, -2.691) | <0.001 | **<0.001** | reference |
|  |  | 6/2016-12/2016 | -0.405 (1.641 ,0.830) | 0.507 | >0.999 | **<0.001** |
|  |  | 6/2017-12/2017 | -0.652 (-1.834, 0.529) | 0.267 | >0.999 | **<0.001** |
|  |  | 6/2018-12/2018 | -0.726 (-2.103 ,0.651) | 0.289 | >0.999 | **<0.001** |
|  |  | 6/2019-12/2019 | -0.076 (-1.008 ,0.856) | 0.868 | >0.999 | **<0.001** |
| HA | Female adults number of contacts (#) | 6/2020-12/2020 | -2.056 (-3.331, -0.782) | 0.003 | 0.013 | reference |
|  |  | 6/2016-12/2016 | -2.992 (-4.296, -1.687) | <0.001 | <0.001 | 0.246 |
|  |  | 6/2017-12/2017 | -1.879 (-3.117, -0.641) | 0.004 | 0.022 | 0.825 |
|  |  | 6/2018-12/2018 | 0.038 (-0.913, 0.989) | 0.935 | >0.999 | 0.01 |
|  |  | 6/2019-12/2019 | -1.408 (-2.411, -0.405) | 0.008 | 0.038 | 0.420 |

^a^ Slope and CI columns holds the monthly change in the weekly outcomes and the confidence interval

^b^ Bonferroni adjusted

^c^ p-value for the interaction terms between time and counterfactual period. P<0.05 for a counterfactual period means that the slope for that year is different than that in our COVID-19 reference period.

**Supplementary Table 4**. Summary of RORs comparing change in proportion of people who self-harmed and were in contact with GP, ED and/or hospital admissions (HA) between reference and target periods to the respective changes in previous years.

|  |  | |  | | Year as |  |  | | | | |  |  |
| --- | --- | --- | --- | --- | --- | --- | --- | --- | --- | --- | --- | --- | --- |
| Setting | Reference period^a^ | | Target period^a^ | | counterfactual | RRR/ROR^b^ | 95% CI | | | | | *p*-value | *p*-value* |
| GP | week 1-10 | 30/12/2019 | week 14-18 | 30/03/2020 | 2016-2017 | 1.198 | ( | 1.048 | , | 1.370 | ) | 0.008 | **0.025** |
|  |  | to |  | to | 2017-2018 | 1.211 | ( | 1.056 | , | 1.388 | ) | 0.006 | **0.018** |
|  |  | 08/03/2020 |  | 03/05/2020 | 2018-2019 | 1.301 | ( | 1.135 | , | 1.493 | ) | <0.001 | **<0.001** |
| ED | week 1-10 | 30/12/2019 | week 28-33 | 06/07/2020 | 2016-2017 | 1.165 | ( | 1.085 | , | 1.251 | ) | <0.001 | **<0.001** |
|  |  | to |  | to | 2017-2018 | 1.096 | ( | 1.023 | , | 1.175 | ) | 0.009 | **0.027** |
|  |  | 08/03/2020 |  | 16/08/2020 | 2018-2019 | 1.117 | ( | 1.041 | , | 1.198 | ) | 0.002 | **0.006** |
| HA | week 1-10 | 30/12/2019 | week 58-61 | 01/02/2021 | 2016-2017 | 0.705 | ( | 0.605 | , | 0.821 | ) | <0.001 | **<0.001** |
|  |  | to |  | to | 2017-2018 | 0.830 | ( | 0.707 | , | 0.975 | ) | 0.023 | 0.070 |
|  |  | 08/03/2020 |  | 28/02/2021 | 2018-2019 | 0.591 | ( | 0.508 | , | 0.688 | ) | <0.001 | **<0.001** |
| GP only | week 1-10 | 30/12/2019 | week 14-18 | 30/03/2020 | 2016-2017 | 1.151 | ( | 0.957 | , | 1.383 | ) | 0.135 | 0.405 |
|  |  | to |  | to | 2017-2018 | 1.198 | ( | 0.992 | , | 1.446 | ) | 0.061 | 0.183 |
|  |  | 08/03/2020 |  | 03/05/2020 | 2018-2019 | 1.291 | ( | 1.073 | , | 1.553 | ) | 0.007 | **0.020** |
| GP only | week 1-10 | 30/12/2019 | week 28-33 | 06/07/2020 | 2016-2017 | 0.790 | ( | 0.659 | , | 0.947 | ) | 0.011 | **0.032** |
|  |  | to |  | to | 2017-2018 | 0.791 | ( | 0.660 | , | 0.948 | ) | 0.011 | **0.033** |
|  |  | 08/03/2020 |  | 16/08/2020 | 2018-2019 | 0.875 | ( | 0.733 | , | 1.044 | ) | 0.139 | 0.416 |
| ED only | week 1-10 | 30/12/2019 | week 12-15 | 16/03/2020 | 2016-2017 | 0.905 | ( | 0.786 | , | 1.041 | ) | 0.162 | 0.487 |
|  |  | to |  | to | 2017-2018 | 0.916 | ( | 0.794 | , | 1.056 | ) | 0.225 | 0.674 |
|  |  | 08/03/2020 |  | 12/04/2021 | 2018-2019 | 0.830 | ( | 0.720 | , | 0.957 | ) | 0.010 | **0.031** |
| ED only | week 1-10 | 30/12/2019 | week 50-53 | 07/12/2020 | 2016-2017 | 1.243 | ( | 1.105 | , | 1.400 | ) | <0.001 | **<0.001** |
|  |  | to |  | to | 2017-2018 | 1.297 | ( | 1.150 | , | 1.463 | ) | <0.001 | **<0.001** |
|  |  | 08/03/2020 |  | 03/01/2021 | 2018-2019 | 1.283 | ( | 1.136 | , | 1.449 | ) | <0.001 | **<0.001** |
| ED only | week 1-10 | 30/12/2019 | week 58-61 | 01/02/2021 | 2016-2017 | 1.254 | ( | 1.120 | , | 1.404 | ) | <0.001 | **<0.001** |
|  |  | to |  | to | 2017-2018 | 1.228 | ( | 1.092 | , | 1.380 | ) | <0.001 | **0.002** |
|  |  | 08/03/2020 |  | 28/02/2021 | 2018-2019 | 1.328 | ( | 1.180 | , | 1.495 | ) | <0.001 | **<0.001** |
| GP, ED & HA only | week 1-10 | 30/12/2019 | week 14-18 | 30/03/2020 | 2016-2017 | 2.101 | ( | 1.280 | , | 3.447 | ) | 0.003 | **0.010** |
|  |  | to |  | to | 2017-2018 | 2.794 | ( | 1.589 | , | 4.912 | ) | <0.001 | **0.001** |
|  |  | 08/03/2020 |  | 03/05/2020 | 2018-2019 | 2.038 | ( | 1.231 | , | 3.372 | ) | 0.006 | **0.017** |
| GP, ED & HA only | week 1-10 | 30/12/2019 | week 28-33 | 06/07/2020 | 2016-2017 | 1.939 | ( | 1.215 | , | 3.094 | ) | 0.005 | **0.016** |
|  |  | to |  | to | 2017-2018 | 1.865 | ( | 1.161 | , | 2.996 | ) | 0.010 | **0.030** |
|  |  | 08/03/2020 |  | 16/08/2020 | 2018-2019 | 2.550 | ( | 1.577 | , | 4.121 | ) | <0.001 | **<0.001** |
| * Bonferroni corrected | | | | | | | | | | | | | |
| ^a^ Period > 1 week represented by the mean of the model coefficients within the period | | | | | | | | | | | | | |
| ^b^ RRR-ratio of rate ratios for prevalence/incidence outcomes; ROR-ratio of odds ratio for proportion outcomes | | | | | | | | | | | | | |

**Supplementary Table 5**. Summary of RRR/RORs based on the difference-in-difference (DiD) approach comparing changes in number and proportion of self-harm contacts per method across GP, ED and hospital admissions between reference and target periods to the respective changes in previous years (as counterfactual).

|  |  |  |  | Wave 1 | | | | | | | |  | Wave 2 | | | | | | | |
| --- | --- | --- | --- | --- | --- | --- | --- | --- | --- | --- | --- | --- | --- | --- | --- | --- | --- | --- | --- | --- |
|  |  |  |  | Reference period: week 1-10 (30/12/2019-08/03/2020) | | | | | | | |  | Reference period: week 1-10 (30/12/2019-08/03/2020) | | | | | | | |
|  | Method of |  | Year as | Target period: week 11-33 (03/09/2019-16/08/2020) | | | | | | | |  | Target period: week 34 onwards (17/08/2019 onwards) | | | | | | | |
| Setting | Self-harm | Outcome | counterfactual | RRR/ROR | 95% CI | | | | | *p*-value | *p*-value* |  | RRR/ROR | 95% CI | | | | | *p*-value | *p*-value* |
| ED | Burning | numbers | 2016 | 2.667 | ( | 0.492 | , | 14.461 | ) | 0.256 | >0.999 |  | 2.000 | ( | 0.271 | , | 14.784 | ) | 0.497 | >0.999 |
|  |  |  | 2017 | 1.167 | ( | 0.208 | , | 6.559 | ) | 0.861 | >0.999 |  | 0.833 | ( | 0.114 | , | 6.111 | ) | 0.858 | >0.999 |
|  |  |  | 2018 | 1.037 | ( | 0.173 | , | 6.233 | ) | 0.968 | >0.999 |  | 2.000 | ( | 0.194 | , | 20.614 | ) | 0.560 | >0.999 |
|  |  |  | 2019 | 2.800 | ( | 0.463 | , | 16.929 | ) | 0.262 | >0.999 |  | 2.000 | ( | 0.241 | , | 16.612 | ) | 0.521 | >0.999 |
| ED |  | proportion | 2016 | 3.122 | ( | 0.576 | , | 16.912 | ) | 0.187 | 0.746 |  | 1.952 | ( | 0.264 | , | 14.416 | ) | 0.512 | >0.999 |
|  |  |  | 2017 | 1.495 | ( | 0.266 | , | 8.398 | ) | 0.648 | >0.999 |  | 0.862 | ( | 0.118 | , | 6.314 | ) | 0.884 | >0.999 |
|  |  |  | 2018 | 1.272 | ( | 0.212 | , | 7.639 | ) | 0.792 | >0.999 |  | 2.049 | ( | 0.199 | , | 21.103 | ) | 0.546 | >0.999 |
|  |  |  | 2019 | 3.291 | ( | 0.545 | , | 19.876 | ) | 0.194 | 0.777 |  | 1.827 | ( | 0.220 | , | 15.159 | ) | 0.577 | >0.999 |
| ED | Injuries | numbers | 2016 | 1.054 | ( | 0.845 | , | 1.316 | ) | 0.640 | >0.999 |  | 1.016 | ( | 0.804 | , | 1.283 | ) | 0.896 | >0.999 |
|  |  |  | 2017 | 0.909 | ( | 0.728 | , | 1.134 | ) | 0.396 | >0.999 |  | 1.052 | ( | 0.831 | , | 1.333 | ) | 0.672 | >0.999 |
|  |  |  | 2018 | 0.736 | ( | 0.586 | , | 0.924 | ) | 0.008 | **0.033** |  | 0.815 | ( | 0.640 | , | 1.038 | ) | 0.098 | 0.391 |
|  |  |  | 2019 | 0.662 | ( | 0.523 | , | 0.838 | ) | <0.001 | **0.002** |  | 0.983 | ( | 0.760 | , | 1.271 | ) | 0.896 | >0.999 |
| ED |  | proportion | 2016 | 1.234 | ( | 0.999 | , | 1.526 | ) | 0.052 | 0.206 |  | 0.992 | ( | 0.792 | , | 1.241 | ) | 0.941 | >0.999 |
|  |  |  | 2017 | 1.165 | ( | 0.942 | , | 1.440 | ) | 0.159 | 0.636 |  | 1.089 | ( | 0.867 | , | 1.366 | ) | 0.464 | >0.999 |
|  |  |  | 2018 | 0.903 | ( | 0.726 | , | 1.124 | ) | 0.362 | >0.999 |  | 0.835 | ( | 0.662 | , | 1.054 | ) | 0.129 | 0.518 |
|  |  |  | 2019 | 0.778 | ( | 0.620 | , | 0.977 | ) | 0.030 | 0.122 |  | 0.898 | ( | 0.700 | , | 1.151 | ) | 0.396 | >0.999 |
| ED | Poisoning | numbers | 2016 | 0.826 | ( | 0.703 | , | 0.972 | ) | 0.021 | 0.084 |  | 0.887 | ( | 0.749 | , | 1.051 | ) | 0.167 | 0.667 |
|  |  |  | 2017 | 0.761 | ( | 0.647 | , | 0.895 | ) | <0.001 | **0.004** |  | 0.966 | ( | 0.812 | , | 1.148 | ) | 0.691 | >0.999 |
|  |  |  | 2018 | 0.829 | ( | 0.703 | , | 0.978 | ) | 0.027 | 0.106 |  | 0.898 | ( | 0.755 | , | 1.068 | ) | 0.222 | 0.889 |
|  |  |  | 2019 | 0.860 | ( | 0.732 | , | 1.012 | ) | 0.069 | 0.276 |  | 1.043 | ( | 0.878 | , | 1.239 | ) | 0.633 | >0.999 |
| ED |  | proportion | 2016 | 0.968 | ( | 0.834 | , | 1.123 | ) | 0.664 | >0.999 |  | 0.866 | ( | 0.741 | , | 1.012 | ) | 0.071 | 0.283 |
|  |  |  | 2017 | 0.975 | ( | 0.841 | , | 1.132 | ) | 0.744 | >0.999 |  | 0.999 | ( | 0.852 | , | 1.171 | ) | 0.989 | >0.999 |
|  |  |  | 2018 | 1.018 | ( | 0.873 | , | 1.185 | ) | 0.823 | >0.999 |  | 0.920 | ( | 0.783 | , | 1.080 | ) | 0.307 | >0.999 |
|  |  |  | 2019 | 1.011 | ( | 0.871 | , | 1.174 | ) | 0.882 | >0.999 |  | 0.953 | ( | 0.812 | , | 1.117 | ) | 0.551 | >0.999 |
| GP | Hanging | numbers | 2016 | 2.667 | ( | 0.892 | , | 7.976 | ) | 0.079 | 0.317 |  | 1.091 | ( | 0.304 | , | 3.910 | ) | 0.894 | >0.999 |
|  |  |  | 2017 | 1.123 | ( | 0.377 | , | 3.341 | ) | 0.835 | >0.999 |  | 0.571 | ( | 0.157 | , | 2.074 | ) | 0.395 | >0.999 |
|  |  |  | 2018 | 1.504 | ( | 0.479 | , | 4.728 | ) | 0.485 | >0.999 |  | 0.667 | ( | 0.177 | , | 2.517 | ) | 0.550 | >0.999 |
|  |  |  | 2019 | 1.778 | ( | 0.567 | , | 5.577 | ) | 0.324 | >0.999 |  | 1.143 | ( | 0.284 | , | 4.595 | ) | 0.851 | >0.999 |
| GP |  | proportion | 2016 | 3.061 | ( | 1.027 | , | 9.127 | ) | 0.045 | 0.179 |  | 1.086 | ( | 0.304 | , | 3.880 | ) | 0.899 | >0.999 |
|  |  |  | 2017 | 1.323 | ( | 0.446 | , | 3.925 | ) | 0.614 | >0.999 |  | 0.562 | ( | 0.155 | , | 2.033 | ) | 0.380 | >0.999 |
|  |  |  | 2018 | 1.699 | ( | 0.542 | , | 5.323 | ) | 0.363 | >0.999 |  | 0.649 | ( | 0.172 | , | 2.444 | ) | 0.523 | >0.999 |
|  |  |  | 2019 | 2.063 | ( | 0.660 | , | 6.451 | ) | 0.213 | 0.853 |  | 1.098 | ( | 0.274 | , | 4.402 | ) | 0.895 | >0.999 |
| GP | Injuries | numbers | 2016 | 0.908 | ( | 0.763 | , | 1.081 | ) | 0.276 | >0.999 |  | 1.076 | ( | 0.901 | , | 1.285 | ) | 0.418 | >0.999 |
|  |  |  | 2017 | 0.864 | ( | 0.726 | , | 1.029 | ) | 0.101 | 0.406 |  | 1.029 | ( | 0.861 | , | 1.229 | ) | 0.756 | >0.999 |
|  |  |  | 2018 | 0.855 | ( | 0.718 | , | 1.018 | ) | 0.078 | 0.313 |  | 1.071 | ( | 0.896 | , | 1.281 | ) | 0.449 | >0.999 |
|  |  |  | 2019 | 0.783 | ( | 0.659 | , | 0.931 | ) | 0.006 | **0.022** |  | 1.093 | ( | 0.914 | , | 1.307 | ) | 0.330 | >0.999 |
| GP |  | proportion | 2016 | 1.042 | ( | 0.895 | , | 1.213 | ) | 0.597 | >0.999 |  | 1.071 | ( | 0.918 | , | 1.250 | ) | 0.384 | >0.999 |
|  |  |  | 2017 | 1.019 | ( | 0.874 | , | 1.186 | ) | 0.813 | >0.999 |  | 1.011 | ( | 0.867 | , | 1.180 | ) | 0.886 | >0.999 |
|  |  |  | 2018 | 0.966 | ( | 0.830 | , | 1.124 | ) | 0.652 | >0.999 |  | 1.043 | ( | 0.894 | , | 1.218 | ) | 0.591 | >0.999 |
|  |  |  | 2019 | 0.909 | ( | 0.782 | , | 1.056 | ) | 0.213 | 0.853 |  | 1.050 | ( | 0.899 | , | 1.226 | ) | 0.538 | >0.999 |
| GP | Poisoning | numbers | 2016 | 0.865 | ( | 0.781 | , | 0.958 | ) | 0.005 | **0.022** |  | 0.991 | ( | 0.891 | , | 1.102 | ) | 0.863 | >0.999 |
|  |  |  | 2017 | 0.849 | ( | 0.767 | , | 0.941 | ) | 0.002 | **0.007** |  | 1.054 | ( | 0.947 | , | 1.173 | ) | 0.340 | >0.999 |
|  |  |  | 2018 | 0.897 | ( | 0.808 | , | 0.996 | ) | 0.041 | 0.164 |  | 1.018 | ( | 0.914 | , | 1.135 | ) | 0.742 | >0.999 |
|  |  |  | 2019 | 0.887 | ( | 0.800 | , | 0.984 | ) | 0.023 | 0.094 |  | 1.030 | ( | 0.925 | , | 1.147 | ) | 0.591 | >0.999 |
| GP |  | proportion | 2016 | 0.993 | ( | 0.938 | , | 1.051 | ) | 0.811 | >0.999 |  | 0.986 | ( | 0.929 | , | 1.047 | ) | 0.645 | >0.999 |
|  |  |  | 2017 | 1.001 | ( | 0.945 | , | 1.060 | ) | 0.979 | >0.999 |  | 1.036 | ( | 0.975 | , | 1.101 | ) | 0.256 | >0.999 |
|  |  |  | 2018 | 1.013 | ( | 0.954 | , | 1.075 | ) | 0.675 | >0.999 |  | 0.992 | ( | 0.932 | , | 1.055 | ) | 0.794 | >0.999 |
|  |  |  | 2019 | 1.030 | ( | 0.971 | , | 1.092 | ) | 0.327 | >0.999 |  | 0.990 | ( | 0.931 | , | 1.052 | ) | 0.736 | >0.999 |
| HA | Burning | numbers | 2016 | 0.857 | ( | 0.189 | , | 3.888 | ) | 0.842 | >0.999 |  | 0.107 | ( | 0.010 | , | 1.121 | ) | 0.062 | 0.249 |
|  |  |  | 2017 | 0.333 | ( | 0.062 | , | 1.779 | ) | 0.199 | 0.794 |  | 0.086 | ( | 0.007 | , | 1.084 | ) | 0.058 | 0.231 |
|  |  |  | 2018 | 0.700 | ( | 0.168 | , | 2.910 | ) | 0.624 | >0.999 |  | 0.111 | ( | 0.011 | , | 1.127 | ) | 0.063 | 0.252 |
|  |  |  | 2019 | 0.571 | ( | 0.147 | , | 2.228 | ) | 0.420 | >0.999 |  | 0.190 | ( | 0.018 | , | 1.992 | ) | 0.166 | 0.665 |
| HA |  | proportion | 2016 | 0.987 | ( | 0.218 | , | 4.470 | ) | 0.987 | >0.999 |  | 0.121 | ( | 0.012 | , | 1.267 | ) | 0.078 | 0.312 |
|  |  |  | 2017 | 0.394 | ( | 0.074 | , | 2.100 | ) | 0.275 | >0.999 |  | 0.096 | ( | 0.008 | , | 1.207 | ) | 0.070 | 0.278 |
|  |  |  | 2018 | 0.786 | ( | 0.189 | , | 3.259 | ) | 0.740 | >0.999 |  | 0.131 | ( | 0.013 | , | 1.331 | ) | 0.086 | 0.343 |
|  |  |  | 2019 | 0.525 | ( | 0.135 | , | 2.043 | ) | 0.353 | >0.999 |  | 0.176 | ( | 0.017 | , | 1.835 | ) | 0.146 | 0.585 |
| HA | Hanging | numbers | 2016 | 1.086 | ( | 0.537 | , | 2.195 | ) | 0.819 | >0.999 |  | 0.584 | ( | 0.289 | , | 1.178 | ) | 0.133 | 0.532 |
|  |  |  | 2017 | 1.052 | ( | 0.536 | , | 2.064 | ) | 0.883 | >0.999 |  | 0.992 | ( | 0.481 | , | 2.047 | ) | 0.983 | >0.999 |
|  |  |  | 2018 | 1.503 | ( | 0.760 | , | 2.974 | ) | 0.241 | 0.966 |  | 0.960 | ( | 0.480 | , | 1.922 | ) | 0.908 | >0.999 |
|  |  |  | 2019 | 1.425 | ( | 0.744 | , | 2.729 | ) | 0.285 | >0.999 |  | 1.159 | ( | 0.583 | , | 2.301 | ) | 0.674 | >0.999 |
| HA |  | proportion | 2016 | 1.251 | ( | 0.622 | , | 2.517 | ) | 0.530 | >0.999 |  | 0.661 | ( | 0.329 | , | 1.327 | ) | 0.244 | 0.976 |
|  |  |  | 2017 | 1.244 | ( | 0.637 | , | 2.429 | ) | 0.523 | >0.999 |  | 1.106 | ( | 0.539 | , | 2.269 | ) | 0.784 | >0.999 |
|  |  |  | 2018 | 1.687 | ( | 0.857 | , | 3.322 | ) | 0.130 | 0.520 |  | 1.136 | ( | 0.570 | , | 2.261 | ) | 0.717 | >0.999 |
|  |  |  | 2019 | 1.310 | ( | 0.687 | , | 2.496 | ) | 0.412 | >0.999 |  | 1.069 | ( | 0.541 | , | 2.112 | ) | 0.848 | >0.999 |
| HA | Injuries | numbers | 2016 | 0.828 | ( | 0.651 | , | 1.053 | ) | 0.125 | 0.499 |  | 0.841 | ( | 0.656 | , | 1.079 | ) | 0.174 | 0.694 |
|  |  |  | 2017 | 0.734 | ( | 0.580 | , | 0.928 | ) | 0.010 | **0.039** |  | 0.953 | ( | 0.743 | , | 1.222 | ) | 0.703 | >0.999 |
|  |  |  | 2018 | 0.785 | ( | 0.623 | , | 0.989 | ) | 0.040 | 0.159 |  | 0.924 | ( | 0.725 | , | 1.177 | ) | 0.521 | >0.999 |
|  |  |  | 2019 | 0.902 | ( | 0.717 | , | 1.133 | ) | 0.375 | >0.999 |  | 1.053 | ( | 0.828 | , | 1.339 | ) | 0.674 | >0.999 |
| HA |  | proportion | 2016 | 0.954 | ( | 0.761 | , | 1.197 | ) | 0.685 | >0.999 |  | 0.953 | ( | 0.755 | , | 1.202 | ) | 0.683 | >0.999 |
|  |  |  | 2017 | 0.868 | ( | 0.696 | , | 1.081 | ) | 0.206 | 0.825 |  | 1.062 | ( | 0.841 | , | 1.341 | ) | 0.614 | >0.999 |
|  |  |  | 2018 | 0.881 | ( | 0.709 | , | 1.094 | ) | 0.250 | >0.999 |  | 1.093 | ( | 0.871 | , | 1.371 | ) | 0.445 | >0.999 |
|  |  |  | 2019 | 0.829 | ( | 0.669 | , | 1.026 | ) | 0.085 | 0.341 |  | 0.971 | ( | 0.776 | , | 1.216 | ) | 0.799 | >0.999 |
| HA | Poisoning | numbers | 2016 | 0.895 | ( | 0.818 | , | 0.979 | ) | 0.016 | 0.063 |  | 0.912 | ( | 0.829 | , | 1.005 | ) | 0.062 | 0.248 |
|  |  |  | 2017 | 0.852 | ( | 0.779 | , | 0.932 | ) | <0.001 | **0.002** |  | 0.898 | ( | 0.815 | , | 0.989 | ) | 0.029 | 0.116 |
|  |  |  | 2018 | 0.918 | ( | 0.839 | , | 1.005 | ) | 0.064 | 0.255 |  | 0.847 | ( | 0.770 | , | 0.932 | ) | <0.001 | **0.003** |
|  |  |  | 2019 | 1.121 | ( | 1.025 | , | 1.226 | ) | 0.013 | 0.050 |  | 1.108 | ( | 1.007 | , | 1.220 | ) | 0.035 | 0.142 |
| HA |  | proportion | 2016 | 1.031 | ( | 0.993 | , | 1.072 | ) | 0.113 | 0.452 |  | 1.033 | ( | 0.991 | , | 1.077 | ) | 0.126 | 0.504 |
|  |  |  | 2017 | 1.007 | ( | 0.969 | , | 1.047 | ) | 0.718 | >0.999 |  | 1.001 | ( | 0.960 | , | 1.044 | ) | 0.965 | >0.999 |
|  |  |  | 2018 | 1.030 | ( | 0.990 | , | 1.073 | ) | 0.142 | 0.566 |  | 1.002 | ( | 0.960 | , | 1.045 | ) | 0.928 | >0.999 |
|  |  |  | 2019 | 1.030 | ( | 0.990 | , | 1.072 | ) | 0.140 | 0.561 |  | 1.022 | ( | 0.980 | , | 1.067 | ) | 0.308 | >0.999 |
| HA | Jumping | numbers | 2016 | 0.208 | ( | 0.075 | , | 0.574 | ) | 0.002 | **0.010** |  | 0.277 | ( | 0.098 | , | 0.782 | ) | 0.015 | 0.062 |
|  |  |  | 2017 | 0.428 | ( | 0.155 | , | 1.177 | ) | 0.100 | 0.401 |  | 0.428 | ( | 0.155 | , | 1.177 | ) | 0.100 | 0.401 |
|  |  |  | 2018 | 0.684 | ( | 0.268 | , | 1.750 | ) | 0.429 | >0.999 |  | 0.684 | ( | 0.268 | , | 1.750 | ) | 0.429 | >0.999 |
|  |  |  | 2019 | 0.640 | ( | 0.270 | , | 1.515 | ) | 0.310 | >0.999 |  | 1.067 | ( | 0.425 | , | 2.675 | ) | 0.891 | >0.999 |
| HA |  | proportion | 2016 | 0.239 | ( | 0.087 | , | 0.659 | ) | 0.006 | **0.023** |  | 0.314 | ( | 0.111 | , | 0.883 | ) | 0.028 | 0.112 |
|  |  |  | 2017 | 0.506 | ( | 0.184 | , | 1.388 | ) | 0.186 | 0.742 |  | 0.477 | ( | 0.174 | , | 1.307 | ) | 0.150 | 0.601 |
|  |  |  | 2018 | 0.768 | ( | 0.302 | , | 1.957 | ) | 0.581 | >0.999 |  | 0.810 | ( | 0.318 | , | 2.062 | ) | 0.658 | >0.999 |
|  |  |  | 2019 | 0.588 | ( | 0.249 | , | 1.388 | ) | 0.226 | 0.902 |  | 0.984 | ( | 0.394 | , | 2.458 | ) | 0.973 | >0.999 |
| * Bonferroni corrected | | | | | | | | | | | | | | | | | | | | |

**Supplementary Table 6**. Summary of RORs and RRRs for self-harm contacts stratified by age and sex for GP, ED and hospital admissions (HA)._

|  |  |  |  |  | |  | | Year as |  |  | | | | |  |  |
| --- | --- | --- | --- | --- | --- | --- | --- | --- | --- | --- | --- | --- | --- | --- | --- | --- |
| Setting | Outcome | Variable | Category | Reference period^a^ | | Target period^a^ | | counterfactual | RRR/ROR^b^ | 95% CI | | | | | *p*-value | *p*-value* |
| ED | proportion | Age group | (10-24) vs. (>24) | week 1-10 | 30/12/2019 | week 16-17 | 13/04/2020 | 2016-2017 | 1.669 | ( | 1.168 | , | 2.386 | ) | 0.005 | **0.015** |
|  |  | (years) |  |  | to |  | to | 2017-2018 | 1.840 | ( | 1.270 | , | 2.666 | ) | 0.001 | **0.004** |
|  |  |  |  |  | 08/03/2020 |  | 26/04/2020 | 2018-2019 | 1.560 | ( | 1.089 | , | 2.234 | ) | 0.015 | **0.046** |
|  |  | Sex | Female vs. Male |  |  |  |  | 2016-2017 | 0.812 | ( | 0.574 | , | 1.148 | ) | 0.239 | 0.718 |
|  |  |  |  |  |  |  |  | 2017-2018 | 1.172 | ( | 0.825 | , | 1.665 | ) | 0.375 | >0.999 |
|  |  |  |  |  |  |  |  | 2018-2019 | 0.896 | ( | 0.632 | , | 1.269 | ) | 0.535 | >0.999 |
| ED | proportion | Age group | (10-24) vs. (>24) | week 1-10 | 30/12/2019 | week 41-45 | 05/10/2020 | 2016-2017 | 1.440 | ( | 1.133 | , | 1.830 | ) | 0.003 | **0.009** |
|  |  | (years) |  |  | to |  | to | 2017-2018 | 1.415 | ( | 1.116 | , | 1.794 | ) | 0.004 | **0.013** |
|  |  |  |  |  | 08/03/2020 |  | 08/11/2020 | 2018-2019 | 1.432 | ( | 1.130 | , | 1.816 | ) | 0.003 | **0.009** |
|  |  | Sex | Female vs. Male |  |  |  |  | 2016-2017 | 1.000 | ( | 0.789 | , | 1.267 | ) | 0.998 | >0.999 |
|  |  |  |  |  |  |  |  | 2017-2018 | 1.286 | ( | 1.018 | , | 1.625 | ) | 0.035 | 0.106 |
|  |  |  |  |  |  |  |  | 2018-2019 | 1.033 | ( | 0.817 | , | 1.306 | ) | 0.788 | >0.999 |
| HA | numbers | Age group | (10-24) vs. (>24) | week 1-10 | 30/12/2019 | week 32-63 | 03/08/2020 | 2016-2017 | 1.356 | ( | 1.097 | , | 1.676 | ) | 0.005 | **0.015** |
|  |  | (years) |  |  | to |  | to | 2017-2018 | 1.469 | ( | 1.189 | , | 1.817 | ) | <0.001 | **0.001** |
|  |  |  |  |  | 08/03/2020 |  | 14/03/2021 | 2018-2019 | 1.323 | ( | 1.062 | , | 1.648 | ) | 0.013 | **0.038** |
|  |  | Sex | Female vs. Male |  |  |  |  | 2016-2017 | 1.477 | ( | 1.190 | , | 1.832 | ) | <0.001 | **0.001** |
|  |  |  |  |  |  |  |  | 2017-2018 | 1.376 | ( | 1.110 | , | 1.708 | ) | 0.004 | **0.011** |
|  |  |  |  |  |  |  |  | 2018-2019 | 1.405 | ( | 1.124 | , | 1.755 | ) | 0.003 | **0.008** |
| HA | proportion | Age group | (10-24) vs. (>24) | week 1-10 | 30/12/2019 | week 32-63 | 03/08/2020 | 2016-2017 | 1.432 | ( | 1.094 | , | 1.647 | ) | 0.005 | **0.015** |
|  |  | (years) |  |  | to |  | to | 2017-2018 | 1.455 | ( | 1.185 | , | 1.786 | ) | <0.001 | **0.001** |
|  |  |  |  |  | 08/03/2020 |  | 14/03/2021 | 2018-2019 | 1.388 | ( | 1.121 | , | 1.717 | ) | 0.003 | **0.008** |
|  |  | Sex | Female vs. Male |  |  |  |  | 2016-2017 | 1.468 | ( | 1.189 | , | 1.811 | ) | <0.001 | **0.001** |
|  |  |  |  |  |  |  |  | 2017-2018 | 1.312 | ( | 1.063 | , | 1.618 | ) | 0.011 | **0.034** |
|  |  |  |  |  |  |  |  | 2018-2019 | 1.340 | ( | 1.078 | , | 1.666 | ) | 0.008 | **0.025** |
| * Bonferroni corrected | | | | | | | | | | | | | | | | |
| ^a^ Period > 1 week represented by the mean of the model coefficients within the period | | | | | | | | | | | | | | | | |
| ^b^ RRR-ratio of rate ratios for prevalence/incidence outcomes; ROR-ratio of odds ratio for proportion outcomes | | | | | | | | | | | | | | | | |

**Supplementary Table 7**. Summary of RORs comparing change in proportion of people who self-harm and are in contact with GP, ED and/or hospital admissions (HA) between reference and target periods to the respective changes in previous years stratified by age and sex.

|  |  |  |  | |  | | Year as |  |  | | | | |  |  |
| --- | --- | --- | --- | --- | --- | --- | --- | --- | --- | --- | --- | --- | --- | --- | --- |
| Setting | Variable | Category | Reference period^a^ | | Target period^a^ | | counterfactual | RRR/ROR^b^ | 95% CI | | | | | *p*-value | *p*-value* |
| ED only | Age group | (10-24) vs. (>24) | week 1-10 | 30/12/2019 | week 50-53 | 07/12/2020 | 2016-2017 | 0.760 | ( | 0.585 | , | 0.986 | ) | 0.039 | 0.117 |
|  | (years) |  |  | to |  | to | 2017-2018 | 0.705 | ( | 0.542 | , | 0.916 | ) | 0.009 | **0.027** |
|  |  |  |  | 08/03/2020 |  | 03/01/2021 | 2018-2019 | 0.743 | ( | 0.568 | , | 0.972 | ) | 0.030 | 0.091 |
|  | Sex | Female vs. Male |  |  |  |  | 2016-2017 | 0.741 | ( | 0.585 | , | 0.939 | ) | 0.013 | **0.039** |
|  |  |  |  |  |  |  | 2017-2018 | 0.927 | ( | 0.730 | , | 1.179 | ) | 0.537 | >0.999 |
|  |  |  |  |  |  |  | 2018-2019 | 0.886 | ( | 0.695 | , | 1.129 | ) | 0.328 | 0.984 |
| HA only | Age group | (10-24) vs. (>24) | week 1-10 | 30/12/2019 | week 14-18 | 30/03/2020 | 2016-2017 | 1.437 | ( | 1.040 | , | 1.987 | ) | 0.028 | 0.084 |
|  | (years) |  |  | to |  | to | 2017-2018 | 1.681 | ( | 1.212 | , | 2.333 | ) | 0.002 | **0.006** |
|  |  |  |  | 08/03/2020 |  | 03/05/2020 | 2018-2019 | 1.350 | ( | 0.968 | , | 1.883 | ) | 0.077 | 0.232 |
|  | Sex | Female vs. Male |  |  |  |  | 2016-2017 | 1.692 | ( | 1.227 | , | 2.333 | ) | 0.001 | **0.004** |
|  |  |  |  |  |  |  | 2017-2018 | 1.355 | ( | 0.979 | , | 1.874 | ) | 0.067 | 0.201 |
|  |  |  |  |  |  |  | 2018-2019 | 1.432 | ( | 1.031 | , | 1.988 | ) | 0.032 | 0.096 |
| * Bonferroni corrected | | | | | | | | | | | | | | | |
| ^a^ Period > 1 week represented by the mean of the model coefficients within the period | | | | | | | | | | | | | | | |
| ^b^ RRR-ratio of rate ratios for prevalence/incidence outcomes; ROR-ratio of odds ratio for proportion outcomes | | | | | | | | | | | | | | | |

**Supplementary Table 8**. Summary of RORs and RRRs for change in gradient of self-harm contacts by WIMD quintile for all settings (Any), GP, ED and hospital admissions (HA), as well as for ED presentations with subsequent hospitalisation (ED to HA).

|  |  |  | |  | | Year as |  |  | | | | |  |  |
| --- | --- | --- | --- | --- | --- | --- | --- | --- | --- | --- | --- | --- | --- | --- |
| Setting | Outcome | Reference period^a^ | | Target period^a^ | | counterfactual | RRR/ROR^b^ | 95% CI | | | | | *p*-value | *p*-value* |
| Any | numbers | week 1-10 | 30/12/2019 | week 12-14 | 16/03/2020 | 2016-2017 | 0.986 | ( | 0.900 | , | 1.081 | ) | 0.770 | >0.999 |
|  |  |  | to |  | to | 2017-2018 | 0.903 | ( | 0.823 | , | 0.990 | ) | 0.029 | 0.087 |
|  |  |  | 08/03/2020 |  | 05/04/2020 | 2018-2019 | 0.979 | ( | 0.892 | , | 1.076 | ) | 0.663 | >0.999 |
| Any | proportion | week 1-10 | 30/12/2019 | week 12-14 | 16/03/2020 | 2016-2017 | 0.993 | ( | 0.913 | , | 1.080 | ) | 0.864 | >0.999 |
|  |  |  | to |  | to | 2017-2018 | 0.913 | ( | 0.839 | , | 0.994 | ) | 0.035 | 0.105 |
|  |  |  | 08/03/2020 |  | 05/04/2020 | 2018-2019 | 0.987 | ( | 0.904 | , | 1.076 | ) | 0.761 | >0.999 |
| Any | numbers | week 1-10 | 30/12/2019 | week 30-33 | 20/07/2020 | 2016-2017 | 1.077 | ( | 1.000 | , | 1.161 | ) | 0.052 | 0.155 |
|  |  |  | to |  | to | 2017-2018 | 1.029 | ( | 0.955 | , | 1.108 | ) | 0.456 | >0.999 |
|  |  |  | 08/03/2020 |  | 16/08/2020 | 2018-2019 | 1.093 | ( | 1.015 | , | 1.177 | ) | 0.019 | 0.056 |
| Any | proportion | week 1-10 | 30/12/2019 | week 30-33 | 20/07/2020 | 2016-2017 | 1.074 | ( | 1.002 | , | 1.151 | ) | 0.043 | 0.129 |
|  |  |  | to |  | to | 2017-2018 | 1.039 | ( | 0.970 | , | 1.112 | ) | 0.277 | 0.832 |
|  |  |  | 08/03/2020 |  | 16/08/2020 | 2018-2019 | 1.100 | ( | 1.027 | , | 1.178 | ) | 0.006 | **0.019** |
| Any | numbers | week 1-10 | 30/12/2019 | week 50-53 | 07/12/2020 | 2016-2017 | 1.027 | ( | 0.945 | , | 1.117 | ) | 0.530 | >0.999 |
|  |  |  | to |  | to | 2017-2018 | 1.018 | ( | 0.938 | , | 1.105 | ) | 0.668 | >0.999 |
|  |  |  | 08/03/2020 |  | 03/01/2021 | 2018-2019 | 1.002 | ( | 0.922 | , | 1.089 | ) | 0.968 | >0.999 |
| Any | proportion | week 1-10 | 30/12/2019 | week 50-53 | 07/12/2020 | 2016-2017 | 1.026 | ( | 0.953 | , | 1.104 | ) | 0.500 | >0.999 |
|  |  |  | to |  | to | 2017-2018 | 1.038 | ( | 0.965 | , | 1.116 | ) | 0.319 | 0.957 |
|  |  |  | 08/03/2020 |  | 03/01/2021 | 2018-2019 | 0.995 | ( | 0.923 | , | 1.072 | ) | 0.887 | >0.999 |
| GP | numbers | week 1-10 | 30/12/2019 | week 12-14 | 16/03/2020 | 2016-2017 | 0.837 | ( | 0.714 | , | 0.982 | ) | 0.029 | 0.086 |
|  |  |  | to |  | to | 2017-2018 | 0.860 | ( | 0.734 | , | 1.008 | ) | 0.063 | 0.188 |
|  |  |  | 08/03/2020 |  | 05/04/2020 | 2018-2019 | 0.937 | ( | 0.797 | , | 1.102 | ) | 0.435 | >0.999 |
| GP | proportion | week 1-10 | 30/12/2019 | week 12-14 | 16/03/2020 | 2016-2017 | 0.853 | ( | 0.729 | , | 0.997 | ) | 0.046 | 0.139 |
|  |  |  | to |  | to | 2017-2018 | 0.882 | ( | 0.755 | , | 1.030 | ) | 0.114 | 0.341 |
|  |  |  | 08/03/2020 |  | 05/04/2020 | 2018-2019 | 0.952 | ( | 0.812 | , | 1.116 | ) | 0.545 | >0.999 |
| GP | numbers | week 1-10 | 30/12/2019 | week 30-33 | 20/07/2020 | 2016-2017 | 1.037 | ( | 0.907 | , | 1.185 | ) | 0.597 | >0.999 |
|  |  |  | to |  | to | 2017-2018 | 1.065 | ( | 0.932 | , | 1.218 | ) | 0.356 | >0.999 |
|  |  |  | 08/03/2020 |  | 16/08/2020 | 2018-2019 | 1.019 | ( | 0.893 | , | 1.163 | ) | 0.779 | >0.999 |
| GP | proportion | week 1-10 | 30/12/2019 | week 30-33 | 20/07/2020 | 2016-2017 | 1.059 | ( | 0.928 | , | 1.207 | ) | 0.396 | >0.999 |
|  |  |  | to |  | to | 2017-2018 | 1.071 | ( | 0.939 | , | 1.222 | ) | 0.307 | 0.922 |
|  |  |  | 08/03/2020 |  | 16/08/2020 | 2018-2019 | 1.035 | ( | 0.909 | , | 1.179 | ) | 0.600 | >0.999 |
| GP | numbers | week 1-10 | 30/12/2019 | week 50-53 | 07/12/2020 | 2016-2017 | 1.021 | ( | 0.880 | , | 1.186 | ) | 0.782 | >0.999 |
|  |  |  | to |  | to | 2017-2018 | 1.038 | ( | 0.898 | , | 1.200 | ) | 0.614 | >0.999 |
|  |  |  | 08/03/2020 |  | 03/01/2021 | 2018-2019 | 0.939 | ( | 0.808 | , | 1.092 | ) | 0.415 | >0.999 |
| GP | proportion | week 1-10 | 30/12/2019 | week 50-53 | 07/12/2020 | 2016-2017 | 1.007 | ( | 0.875 | , | 1.160 | ) | 0.919 | >0.999 |
|  |  |  | to |  | to | 2017-2018 | 1.046 | ( | 0.910 | , | 1.202 | ) | 0.531 | >0.999 |
|  |  |  | 08/03/2020 |  | 03/01/2021 | 2018-2019 | 0.950 | ( | 0.823 | , | 1.097 | ) | 0.484 | >0.999 |
| ED | numbers | week 1-10 | 30/12/2019 | week 12-14 | 16/03/2020 | 2016-2017 | 1.107 | ( | 0.994 | , | 1.234 | ) | 0.064 | 0.192 |
|  |  |  | to |  | to | 2017-2018 | 0.953 | ( | 0.854 | , | 1.063 | ) | 0.388 | >0.999 |
|  |  |  | 08/03/2020 |  | 05/04/2020 | 2018-2019 | 1.046 | ( | 0.936 | , | 1.169 | ) | 0.425 | >0.999 |
| ED | proportion | week 1-10 | 30/12/2019 | week 16-17 | 13/04/2020 | 2016-2017 | 1.014 | ( | 0.889 | , | 1.156 | ) | 0.841 | >0.999 |
|  |  |  | to |  | to | 2017-2018 | 1.054 | ( | 0.924 | , | 1.202 | ) | 0.435 | >0.999 |
|  |  |  | 08/03/2020 |  | 26/04/2020 | 2018-2019 | 0.995 | ( | 0.873 | , | 1.135 | ) | 0.946 | >0.999 |
| ED | numbers | week 1-10 | 30/12/2019 | week 30-33 | 20/07/2020 | 2016-2017 | 1.127 | ( | 1.036 | , | 1.227 | ) | 0.005 | **0.016** |
|  |  |  | to |  | to | 2017-2018 | 0.983 | ( | 0.905 | , | 1.068 | ) | 0.680 | >0.999 |
|  |  |  | 08/03/2020 |  | 16/08/2020 | 2018-2019 | 1.090 | ( | 1.004 | , | 1.184 | ) | 0.040 | 0.121 |
| ED | proportion | week 1-10 | 30/12/2019 | week 41-45 | 05/10/2020 | 2016-2017 | 1.079 | ( | 0.987 | , | 1.179 | ) | 0.094 | 0.282 |
|  |  |  | to |  | to | 2017-2018 | 0.948 | ( | 0.868 | , | 1.035 | ) | 0.230 | 0.689 |
|  |  |  | 08/03/2020 |  | 08/11/2020 | 2018-2019 | 1.041 | ( | 0.954 | , | 1.136 | ) | 0.370 | >0.999 |
| ED | numbers | week 1-10 | 30/12/2019 | week 50-53 | 07/12/2020 | 2016-2017 | 1.080 | ( | 0.981 | , | 1.189 | ) | 0.115 | 0.346 |
|  |  |  | to |  | to | 2017-2018 | 1.041 | ( | 0.947 | , | 1.144 | ) | 0.405 | >0.999 |
|  |  |  | 08/03/2020 |  | 03/01/2021 | 2018-2019 | 0.990 | ( | 0.899 | , | 1.090 | ) | 0.836 | >0.999 |
| ED | proportion | week 1-10 | 30/12/2019 | week 53-58 | 28/12/2020 | 2016-2017 | 1.123 | ( | 1.035 | , | 1.218 | ) | 0.005 | **0.015** |
|  |  |  | to |  | to | 2017-2018 | 0.988 | ( | 0.910 | , | 1.072 | ) | 0.768 | >0.999 |
|  |  |  | 08/03/2020 |  | 07/02/2021 | 2018-2019 | 1.058 | ( | 0.976 | , | 1.147 | ) | 0.174 | 0.521 |
| ED to HA | numbers | week 1-10 | 30/12/2019 | week 12-14 | 16/03/2020 | 2016-2017 | 1.103 | ( | 0.891 | , | 1.364 | ) | 0.368 | >0.999 |
|  |  |  | to |  | to | 2017-2018 | 0.993 | ( | 0.801 | , | 1.229 | ) | 0.946 | >0.999 |
|  |  |  | 08/03/2020 |  | 05/04/2020 | 2018-2019 | 1.079 | ( | 0.867 | , | 1.345 | ) | 0.495 | >0.999 |
| ED to HA | proportion | week 1-10 | 30/12/2019 | week 19-24 | 04/05/2020 | 2016-2017 | 1.017 | ( | 0.908 | , | 1.139 | ) | 0.769 | >0.999 |
|  |  |  | to |  | to | 2017-2018 | 0.953 | ( | 0.852 | , | 1.067 | ) | 0.406 | >0.999 |
|  |  |  | 08/03/2020 |  | 14/06/2020 | 2018-2019 | 0.997 | ( | 0.895 | , | 1.110 | ) | 0.956 | >0.999 |
| ED to HA | numbers | week 1-10 | 30/12/2019 | week 30-33 | 20/07/2020 | 2016-2017 | 1.153 | ( | 0.974 | , | 1.364 | ) | 0.099 | 0.297 |
|  |  |  | to |  | to | 2017-2018 | 0.996 | ( | 0.847 | , | 1.172 | ) | 0.961 | >0.999 |
|  |  |  | 08/03/2020 |  | 16/08/2020 | 2018-2019 | 1.072 | ( | 0.911 | , | 1.262 | ) | 0.401 | >0.999 |
| ED to HA | proportion | week 1-10 | 30/12/2019 | week 36-38 | 31/08/2020 | 2016-2017 | 1.002 | ( | 0.850 | , | 1.182 | ) | 0.979 | >0.999 |
|  |  |  | to |  | to | 2017-2018 | 0.911 | ( | 0.768 | , | 1.079 | ) | 0.280 | 0.841 |
|  |  |  | 08/03/2020 |  | 20/09/2020 | 2018-2019 | 0.842 | ( | 0.708 | , | 1.000 | ) | 0.050 | 0.149 |
| ED to HA | numbers | week 1-10 | 30/12/2019 | week 50-53 | 07/12/2020 | 2016-2017 | 1.091 | ( | 0.890 | , | 1.336 | ) | 0.402 | >0.999 |
|  |  |  | to |  | to | 2017-2018 | 0.986 | ( | 0.808 | , | 1.203 | ) | 0.891 | >0.999 |
|  |  |  | 08/03/2020 |  | 03/01/2021 | 2018-2019 | 0.895 | ( | 0.730 | , | 1.096 | ) | 0.282 | 0.846 |
| ED to HA | proportion | week 1-10 | 30/12/2019 | week 47-60 | 16/11/2020 | 2016-2017 | 1.038 | ( | 0.935 | , | 1.153 | ) | 0.483 | >0.999 |
|  |  |  | to |  | to | 2017-2018 | 0.947 | ( | 0.855 | , | 1.048 | ) | 0.295 | 0.884 |
|  |  |  | 08/03/2020 |  | 21/02/2021 | 2018-2019 | 0.981 | ( | 0.887 | , | 1.085 | ) | 0.705 | >0.999 |
| HA | numbers | week 1-10 | 30/12/2019 | week 12-14 | 16/03/2020 | 2016-2017 | 0.931 | ( | 0.803 | , | 1.079 | ) | 0.343 | >0.999 |
|  |  |  | to |  | to | 2017-2018 | 0.865 | ( | 0.746 | , | 1.004 | ) | 0.056 | 0.169 |
|  |  |  | 08/03/2020 |  | 05/04/2020 | 2018-2019 | 0.944 | ( | 0.807 | , | 1.104 | ) | 0.472 | >0.999 |
| HA | proportion | week 1-10 | 30/12/2019 | week 15-33 | 06/04/2020 | 2016-2017 | 0.993 | ( | 0.918 | , | 1.075 | ) | 0.863 | >0.999 |
|  |  |  | to |  | to | 2017-2018 | 0.951 | ( | 0.879 | , | 1.028 | ) | 0.207 | 0.620 |
|  |  |  | 08/03/2020 |  | 16/08/2020 | 2018-2019 | 0.999 | ( | 0.921 | , | 1.085 | ) | 0.988 | >0.999 |
| HA | numbers | week 1-10 | 30/12/2019 | week 30-33 | 20/07/2020 | 2016-2017 | 1.051 | ( | 0.927 | , | 1.191 | ) | 0.439 | >0.999 |
|  |  |  | to |  | to | 2017-2018 | 1.038 | ( | 0.916 | , | 1.175 | ) | 0.560 | >0.999 |
|  |  |  | 08/03/2020 |  | 16/08/2020 | 2018-2019 | 1.162 | ( | 1.025 | , | 1.316 | ) | 0.019 | 0.056 |
| HA | proportion | week 1-10 | 30/12/2019 | week 41-44 | 05/10/2020 | 2016-2017 | 0.963 | ( | 0.852 | , | 1.090 | ) | 0.552 | >0.999 |
|  |  |  | to |  | to | 2017-2018 | 0.851 | ( | 0.751 | , | 0.964 | ) | 0.011 | **0.033** |
|  |  |  | 08/03/2020 |  | 01/11/2020 | 2018-2019 | 1.001 | ( | 0.884 | , | 1.132 | ) | 0.993 | >0.999 |
| HA | numbers | week 1-10 | 30/12/2019 | week 30-33 | 20/07/2020 | 2016-2017 | 0.984 | ( | 0.846 | , | 1.144 | ) | 0.832 | >0.999 |
|  |  |  | to |  | to | 2017-2018 | 0.941 | ( | 0.813 | , | 1.090 | ) | 0.419 | >0.999 |
|  |  |  | 08/03/2020 |  | 16/08/2020 | 2018-2019 | 0.960 | ( | 0.827 | , | 1.114 | ) | 0.590 | >0.999 |
| HA | proportion | week 1-10 | 30/12/2019 | week 50-53 | 07/12/2020 | 2016-2017 | 0.975 | ( | 0.848 | , | 1.122 | ) | 0.725 | >0.999 |
|  |  |  | to |  | to | 2017-2018 | 0.939 | ( | 0.819 | , | 1.077 | ) | 0.367 | >0.999 |
|  |  |  | 08/03/2020 |  | 03/01/2021 | 2018-2019 | 0.956 | ( | 0.832 | , | 1.099 | ) | 0.527 | >0.999 |
| * Bonferroni corrected | | | | | | | | | | | | | | |
| ^a^ Period > 1 week represented by the mean of the model coefficients within the period | | | | | | | | | | | | | | |
| ^b^ RRR-ratio of rate ratios for prevalence/incidence outcomes; ROR-ratio of odds ratio for proportion outcomes | | | | | | | | | | | | | | |

**Supplementary Table 9**. Summary of RORs comparing change in proportion of people who self-harmed and were in contract with GP, ED and/or hospital admissions between reference and target periods to the respective changes in previous years stratified by WIMD deprivation level.

|  |  | |  | | Year as |  |  | | | | |  |  |
| --- | --- | --- | --- | --- | --- | --- | --- | --- | --- | --- | --- | --- | --- |
| Setting | Reference period^a^ | | Target period^a^ | | counterfactual | RRR/ROR^b^ | 95% CI | | | | | *p*-value | *p*-value* |
| GP | week 1-10 | 30/12/2019 | week 14-18 | 30/03/2020 | 2016-2017 | 0.921 | ( | 0.835 | , | 1.016 | ) | 0.101 | 0.303 |
|  |  | to |  | to | 2017-2018 | 0.924 | ( | 0.835 | , | 1.022 | ) | 0.123 | 0.369 |
|  |  | 08/03/2020 |  | 03/05/2020 | 2018-2019 | 0.957 | ( | 0.866 | , | 1.058 | ) | 0.390 | >0.999 |
| GP | week 1-10 | 30/12/2019 | week 28-33 | 06/07/2020 | 2016-2017 | 0.942 | ( | 0.857 | , | 1.037 | ) | 0.222 | 0.667 |
|  |  | to |  | to | 2017-2018 | 0.999 | ( | 0.908 | , | 1.098 | ) | 0.977 | >0.999 |
|  |  | 08/03/2020 |  | 16/08/2020 | 2018-2019 | 0.957 | ( | 0.871 | , | 1.052 | ) | 0.363 | >0.999 |
| ED | week 1-10 | 30/12/2019 | week 12-15 | 16/03/2020 | 2016-2017 | 1.094 | ( | 1.015 | , | 1.179 | ) | 0.018 | 0.055 |
|  |  | to |  | to | 2017-2018 | 1.053 | ( | 0.977 | , | 1.136 | ) | 0.177 | 0.532 |
|  |  | 08/03/2020 |  | 12/04/2020 | 2018-2019 | 1.034 | ( | 0.958 | , | 1.116 | ) | 0.396 | >0.999 |
| ED | week 1-10 | 30/12/2019 | week 28-33 | 06/07/2020 | 2016-2017 | 1.045 | ( | 0.989 | , | 1.104 | ) | 0.121 | 0.362 |
|  |  | to |  | to | 2017-2018 | 0.990 | ( | 0.938 | , | 1.044 | ) | 0.711 | >0.999 |
|  |  | 08/03/2020 |  | 16/08/2020 | 2018-2019 | 1.020 | ( | 0.966 | , | 1.077 | ) | 0.472 | >0.999 |
| HA | week 1-10 | 30/12/2019 | week 12-13 | 16/03/2020 | 2016-2017 | 0.926 | ( | 0.814 | , | 1.054 | ) | 0.243 | 0.729 |
|  |  | to |  | to | 2017-2018 | 0.989 | ( | 0.873 | , | 1.121 | ) | 0.868 | >0.999 |
|  |  | 08/03/2020 |  | 29/03/2020 | 2018-2019 | 0.982 | ( | 0.848 | , | 1.136 | ) | 0.803 | >0.999 |
| HA | week 1-10 | 30/12/2019 | week 28-33 | 06/07/2020 | 2016-2017 | 0.981 | ( | 0.899 | , | 1.070 | ) | 0.658 | >0.999 |
|  |  | to |  | to | 2017-2018 | 1.000 | ( | 0.918 | , | 1.089 | ) | 0.998 | >0.999 |
|  |  | 08/03/2020 |  | 16/08/2020 | 2018-2019 | 1.016 | ( | 0.929 | , | 1.111 | ) | 0.730 | >0.999 |
| HA | week 1-10 | 30/12/2019 | week 58-61 | 01/02/2021 | 2016-2017 | 1.014 | ( | 0.906 | , | 1.135 | ) | 0.807 | >0.999 |
|  |  | to |  | to | 2017-2018 | 0.963 | ( | 0.855 | , | 1.085 | ) | 0.536 | >0.999 |
|  |  | 08/03/2020 |  | 28/02/2021 | 2018-2019 | 1.005 | ( | 0.897 | , | 1.126 | ) | 0.933 | >0.999 |
| GP only | week 1-10 | 30/12/2019 | week 14-18 | 30/03/2020 | 2016-2017 | 0.898 | ( | 0.785 | , | 1.027 | ) | 0.117 | 0.350 |
|  |  | to |  | to | 2017-2018 | 0.958 | ( | 0.833 | , | 1.100 | ) | 0.540 | >0.999 |
|  |  | 08/03/2020 |  | 03/05/2020 | 2018-2019 | 0.963 | ( | 0.841 | , | 1.103 | ) | 0.587 | >0.999 |
| GP only | week 1-10 | 30/12/2019 | week 28-33 | 06/07/2020 | 2016-2017 | 0.882 | ( | 0.772 | , | 1.008 | ) | 0.065 | 0.194 |
|  |  | to |  | to | 2017-2018 | 0.960 | ( | 0.840 | , | 1.097 | ) | 0.546 | >0.999 |
|  |  | 08/03/2020 |  | 16/08/2020 | 2018-2019 | 0.893 | ( | 0.784 | , | 1.017 | ) | 0.087 | 0.262 |
| ED only | week 1-10 | 30/12/2019 | week 12-15 | 16/03/2020 | 2016-2017 | 1.120 | ( | 1.006 | , | 1.246 | ) | 0.039 | 0.116 |
|  |  | to |  | to | 2017-2018 | 1.062 | ( | 0.953 | , | 1.184 | ) | 0.277 | 0.831 |
|  |  | 08/03/2020 |  | 12/04/2020 | 2018-2019 | 1.019 | ( | 0.914 | , | 1.137 | ) | 0.732 | >0.999 |
| ED only | week 1-10 | 30/12/2019 | week 28-33 | 06/07/2020 | 2016-2017 | 1.041 | ( | 0.962 | , | 1.127 | ) | 0.321 | 0.963 |
|  |  | to |  | to | 2017-2018 | 1.004 | ( | 0.929 | , | 1.086 | ) | 0.914 | >0.999 |
|  |  | 08/03/2020 |  | 16/08/2020 | 2018-2019 | 1.028 | ( | 0.951 | , | 1.112 | ) | 0.489 | >0.999 |
| ED only | week 1-10 | 30/12/2019 | week 50-53 | 07/12/2020 | 2016-2017 | 1.017 | ( | 0.927 | , | 1.116 | ) | 0.723 | >0.999 |
|  |  | to |  | to | 2017-2018 | 1.048 | ( | 0.957 | , | 1.148 | ) | 0.315 | 0.944 |
|  |  | 08/03/2020 |  | 03/01/2021 | 2018-2019 | 1.036 | ( | 0.944 | , | 1.137 | ) | 0.455 | >0.999 |
| ED only | week 1-10 | 30/12/2019 | week 58-61 | 01/02/2021 | 2016-2017 | 1.027 | ( | 0.942 | , | 1.120 | ) | 0.541 | >0.999 |
|  |  | to |  | to | 2017-2018 | 1.032 | ( | 0.944 | , | 1.129 | ) | 0.488 | >0.999 |
|  |  | 08/03/2020 |  | 28/02/2021 | 2018-2019 | 1.012 | ( | 0.923 | , | 1.110 | ) | 0.796 | >0.999 |
| HA only | week 1-10 | 30/12/2019 | week 14-18 | 30/03/2020 | 2016-2017 | 0.959 | ( | 0.775 | , | 1.186 | ) | 0.697 | >0.999 |
|  |  | to |  | to | 2017-2018 | 0.983 | ( | 0.797 | , | 1.212 | ) | 0.873 | >0.999 |
|  |  | 08/03/2020 |  | 03/05/2020 | 2018-2019 | 0.948 | ( | 0.752 | , | 1.196 | ) | 0.654 | >0.999 |
| HA only | week 1-10 | 30/12/2019 | week 28-33 | 06/07/2020 | 2016-2017 | 0.998 | ( | 0.862 | , | 1.156 | ) | 0.978 | >0.999 |
|  |  | to |  | to | 2017-2018 | 1.063 | ( | 0.918 | , | 1.232 | ) | 0.415 | >0.999 |
|  |  | 08/03/2020 |  | 16/08/2020 | 2018-2019 | 1.026 | ( | 0.882 | , | 1.194 | ) | 0.736 | >0.999 |
| GP & ED only | week 1-10 | 30/12/2019 | week 14-18 | 30/03/2020 | 2016-2017 | 1.607 | ( | 0.990 | , | 2.608 | ) | 0.055 | 0.165 |
|  |  | to |  | to | 2017-2018 | 1.134 | ( | 0.717 | , | 1.795 | ) | 0.591 | >0.999 |
|  |  | 08/03/2020 |  | 03/05/2020 | 2018-2019 | 1.322 | ( | 0.822 | , | 2.125 | ) | 0.250 | 0.749 |
| GP & ED only | week 1-10 | 30/12/2019 | week 27-30 | 29/06/2020 | 2016-2017 | 1.423 | ( | 0.824 | , | 2.459 | ) | 0.206 | 0.617 |
|  |  | to |  | to | 2017-2018 | 1.490 | ( | 0.866 | , | 2.563 | ) | 0.150 | 0.449 |
|  |  | 08/03/2020 |  | 26/07/2020 | 2018-2019 | 1.116 | ( | 0.662 | , | 1.881 | ) | 0.680 | >0.999 |
| GP & ED only | week 1-10 | 30/12/2019 | week 35-36 | 24/08/2020 | 2016-2017 | 0.978 | ( | 0.619 | , | 1.546 | ) | 0.924 | >0.999 |
|  |  | to |  | to | 2017-2018 | 1.011 | ( | 0.626 | , | 1.634 | ) | 0.963 | >0.999 |
|  |  | 08/03/2020 |  | 06/09/2020 | 2018-2019 | 0.941 | ( | 0.580 | , | 1.525 | ) | 0.804 | >0.999 |
| GP & HA only | week 1-10 | 30/12/2019 | week 14-18 | 30/03/2020 | 2016-2017 | 0.776 | ( | 0.553 | , | 1.088 | ) | 0.141 | 0.424 |
|  |  | to |  | to | 2017-2018 | 0.706 | ( | 0.504 | , | 0.991 | ) | 0.044 | 0.132 |
|  |  | 08/03/2020 |  | 03/05/2020 | 2018-2019 | 0.961 | ( | 0.666 | , | 1.386 | ) | 0.829 | >0.999 |
| GP & HA only | week 1-10 | 30/12/2019 | week 28-33 | 06/07/2020 | 2016-2017 | 0.823 | ( | 0.603 | , | 1.123 | ) | 0.218 | 0.655 |
|  |  | to |  | to | 2017-2018 | 0.961 | ( | 0.708 | , | 1.304 | ) | 0.798 | >0.999 |
|  |  | 08/03/2020 |  | 16/08/2020 | 2018-2019 | 1.070 | ( | 0.771 | , | 1.484 | ) | 0.687 | >0.999 |
| ED & HA only | week 1-10 | 30/12/2019 | week 12-15 | 16/03/2020 | 2016-2017 | 1.264 | ( | 0.998 | , | 1.601 | ) | 0.052 | 0.156 |
|  |  | to |  | to | 2017-2018 | 1.104 | ( | 0.871 | , | 1.400 | ) | 0.413 | >0.999 |
|  |  | 08/03/2020 |  | 12/04/2020 | 2018-2019 | 1.170 | ( | 0.923 | , | 1.484 | ) | 0.194 | 0.583 |
| ED & HA only | week 1-10 | 30/12/2019 | week 28-33 | 06/07/2020 | 2016-2017 | 0.984 | ( | 0.809 | , | 1.197 | ) | 0.872 | >0.999 |
|  |  | to |  | to | 2017-2018 | 0.890 | ( | 0.742 | , | 1.069 | ) | 0.212 | 0.637 |
|  |  | 08/03/2020 |  | 16/08/2020 | 2018-2019 | 0.973 | ( | 0.802 | , | 1.179 | ) | 0.777 | >0.999 |
| ED & HA only | week 1-10 | 30/12/2019 | week 58-61 | 01/02/2021 | 2016-2017 | 1.314 | ( | 1.011 | , | 1.707 | ) | 0.041 | 0.124 |
|  |  | to |  | to | 2017-2018 | 0.940 | ( | 0.712 | , | 1.241 | ) | 0.664 | >0.999 |
|  |  | 08/03/2020 |  | 28/02/2021 | 2018-2019 | 1.251 | ( | 0.964 | , | 1.624 | ) | 0.092 | 0.275 |
| GP, ED & HA only | week 1-10 | 30/12/2019 | week 14-18 | 30/03/2020 | 2016-2017 | 0.893 | ( | 0.592 | , | 1.348 | ) | 0.591 | >0.999 |
|  |  | to |  | to | 2017-2018 | 0.967 | ( | 0.620 | , | 1.507 | ) | 0.881 | >0.999 |
|  |  | 08/03/2020 |  | 03/05/2020 | 2018-2019 | 0.961 | ( | 0.632 | , | 1.461 | ) | 0.852 | >0.999 |
| GP, ED & HA only | week 1-10 | 30/12/2019 | week 28-33 | 06/07/2020 | 2016-2017 | 1.030 | ( | 0.694 | , | 1.527 | ) | 0.885 | >0.999 |
|  |  | to |  | to | 2017-2018 | 1.001 | ( | 0.680 | , | 1.473 | ) | 0.997 | >0.999 |
|  |  | 08/03/2020 |  | 16/08/2020 | 2018-2019 | 1.194 | ( | 0.797 | , | 1.789 | ) | 0.391 | >0.999 |
| GP, ED & HA only | week 1-10 | 30/12/2019 | week 58-61 | 01/02/2021 | 2016-2017 | 1.152 | ( | 0.667 | , | 1.989 | ) | 0.612 | >0.999 |
|  |  | to |  | to | 2017-2018 | 1.044 | ( | 0.607 | , | 1.794 | ) | 0.877 | >0.999 |
|  |  | 08/03/2020 |  | 28/02/2021 | 2018-2019 | 0.953 | ( | 0.532 | , | 1.706 | ) | 0.870 | >0.999 |
| * Bonferroni corrected | | | | | | | | | | | | | |
| ^a^ Period > 1 week represented by the mean of the model coefficients within the period | | | | | | | | | | | | | |
| ^b^ RRR-ratio of rate ratios for prevalence/incidence outcomes; ROR-ratio of odds ratio for proportion outcomes | | | | | | | | | | | | | |

**Supplementary Table 10**. Characteristics of the COVID-19 infection cohorts for the main and sensitivity analyses.

|  |  |  | COVID-19 Definition 1 (main analysis) | | | | |  | COVID-19 Definition 2 (sensitivity analysis) | | | | |
| --- | --- | --- | --- | --- | --- | --- | --- | --- | --- | --- | --- | --- | --- |
|  |  |  | Infected | |  | Not infected | |  | Infected | |  | Not infected | |
| Characteristics |  |  | Numbers | % |  | Numbers | % |  | Numbers | % |  | Numbers | % |
|  |  | Total | 23,703 | 0.9 |  | 2,482,673 | 99.1 |  | 13,973 | 0.6 |  | 2,492,347 | 99.4 |
| Sex |  | Male | 8,582 | 36.2 |  | 1,238,561 | 49.9 |  | 4,926 | 35.3 |  | 1,242,082 | 49.8 |
|  |  | Female | 15,121 | 63.8 |  | 1,244,112 | 50.1 |  | 9,047 | 64.7 |  | 1,250,265 | 50.2 |
| Age* |  | 10-24 yr | 1,649 | 7.0 |  | 399,347 | 16.1 |  | 894 | 6.4 |  | 400,025 | 16.1 |
|  |  | 25-64 yr | 15,981 | 67.4 |  | 1,468,433 | 59.1 |  | 9,424 | 67.4 |  | 1,475,132 | 59.2 |
|  |  | 65 yr + | 6,073 | 25.6 |  | 614,893 | 24.8 |  | 3,655 | 26.2 |  | 617,190 | 24.8 |
| Ethnicity | White |  | 19,809 | 83.6 |  | 2,083,525 | 83.9 |  | 11,439 | 81.9 |  | 2,091,727 | 83.9 |
|  | non-White |  | 1,276 | 5.4 |  | 77,727 | 3.1 |  | 914 | 6.5 |  | 78,041 | 3.1 |
|  |  | Black | 125 | 0.5 |  | 10,225 | 0.4 |  | 82 | 0.6 |  | 10,257 | 0.4 |
|  |  | Asian | 902 | 3.8 |  | 38,859 | 1.6 |  | 687 | 4.9 |  | 39,058 | 1.6 |
|  |  | Others/Mixed | 249 | 1.1 |  | 28,643 | 1.2 |  | 145 | 1.0 |  | 28,726 | 1.2 |
|  | Unknown |  | 2,618 | 11.0 |  | 321,421 | 12.9 |  | 1,620 | 11.6 |  | 322,579 | 12.9 |
| WIMD quintile* |  | Q1 | 4,281 | 18.1 |  | 499,847 | 20.1 |  | 2,720 | 19.5 |  | 501,485 | 20.1 |
| (Q5: most deprived) |  | Q2 | 3,799 | 16.0 |  | 483,057 | 19.5 |  | 2,379 | 17.0 |  | 484,413 | 19.4 |
|  |  | Q3 | 4,864 | 20.5 |  | 503,605 | 20.3 |  | 2,674 | 19.1 |  | 505,823 | 20.3 |
|  |  | Q4 | 5,159 | 21.8 |  | 500,733 | 20.2 |  | 3,081 | 22.0 |  | 502,727 | 20.2 |
|  |  | Q5 | 5,600 | 23.6 |  | 495,431 | 20.0 |  | 3,119 | 22.3 |  | 497,899 | 20.0 |
| Urban/rural indicator* |  | Rural | 5,756 | 24.3 |  | 763,558 | 30.8 |  | 3,256 | 23.3 |  | 766,139 | 30.7 |
|  |  | Urban | 17,947 | 75.7 |  | 1,719,115 | 69.2 |  | 10,717 | 76.7 |  | 1,726,208 | 69.3 |
| Health Board* |  | Betsi Cadwaladr | 5,450 | 23.0 |  | 541,148 | 21.8 |  | 3,683 | 26.4 |  | 542,889 | 21.8 |
|  |  | Hywel Dda | 1,473 | 6.2 |  | 299,283 | 12.1 |  | 959 | 6.9 |  | 299,743 | 12.0 |
|  |  | Swansea Bay | 2,791 | 11.8 |  | 314,941 | 12.7 |  | 1,660 | 11.9 |  | 316,160 | 12.7 |
|  |  | Cardiff & Vale | 4,329 | 18.3 |  | 386,169 | 15.6 |  | 2,562 | 18.3 |  | 387,931 | 15.6 |
|  |  | Cwm Taf Morgannwg | 3,854 | 16.3 |  | 364,818 | 14.7 |  | 2,563 | 18.3 |  | 366,132 | 14.7 |
|  |  | Aneurin Bevan | 4,552 | 19.2 |  | 477,790 | 19.2 |  | 2,222 | 15.9 |  | 480,058 | 19.3 |
|  |  | Powys | 1,254 | 5.3 |  | 98,524 | 4.0 |  | 324 | 2.3 |  | 99,434 | 4.0 |
| Live in care homes* |  |  | 1,197 | 5.0 |  | 12,499 | 0.5 |  | 838 | 6.0 |  | 12,820 | 0.5 |
| In COVID-19 shielded list |  |  | 2,341 | 9.9 |  | 136,924 | 5.5 |  | 1,197 | 8.6 |  | 138,100 | 5.5 |
| Charlson Comorbidity Index** |  | 0 | 17,825 | 75.2 |  | 2,184,862 | 88.0 |  | 10,627 | 76.1 |  | 2,191,806 | 87.9 |
| (unweighted) |  | 1 | 3,482 | 14.7 |  | 230,413 | 9.3 |  | 1,786 | 12.8 |  | 232,367 | 9.3 |
|  |  | 2 | 1,313 | 5.5 |  | 47,578 | 1.9 |  | 828 | 5.9 |  | 47,958 | 1.9 |
|  |  | 3+ | 1,083 | 4.6 |  | 19,820 | 0.8 |  | 732 | 5.2 |  | 20,216 | 0.8 |
| Ever smoked*** |  |  | 14,307 | 60.4 |  | 1,238,552 | 49.9 |  | 7,601 | 54.4 |  | 1,245,202 | 50.0 |
| History of self-harm |  |  | 2,070 | 8.7 |  | 138,174 | 5.6 |  | 890 | 6.4 |  | 139,297 | 5.6 |
| History of mental health |  |  | 16,637 | 70.2 |  | 1,333,425 | 53.7 |  | 8,869 | 63.5 |  | 1,341,049 | 53.8 |
| Ever prescribed*** |  | Psychotropics | 14,258 | 60.2 |  | 1,024,121 | 41.3 |  | 7,362 | 52.7 |  | 1,031,056 | 41.4 |
|  |  | Opiates | 16,069 | 67.8 |  | 1,210,567 | 48.8 |  | 8,594 | 61.5 |  | 1,218,109 | 48.9 |
| % of length of residence |  | [0 - 20] | 9,753 | 41.1 |  | 890,150 | 35.9 |  | 5,669 | 40.6 |  | 894,209 | 35.9 |
| in Wales to age at index date |  | (20 - 40] | 7,402 | 31.2 |  | 706,810 | 28.5 |  | 4,466 | 32.0 |  | 709,740 | 28.5 |
|  |  | (40 - 60] | 3,738 | 15.8 |  | 417,283 | 16.8 |  | 2,179 | 15.6 |  | 418,910 | 16.8 |
|  |  | (60 - 80] | 1,078 | 4.5 |  | 146,207 | 5.9 |  | 639 | 4.6 |  | 146,658 | 5.9 |
|  |  | (80 - 100] | 1,732 | 7.3 |  | 322,223 | 13.0 |  | 1,020 | 7.3 |  | 322,830 | 13.0 |
| Number of moves*** |  | 0 | 18,268 | 77.1 |  | 1,975,146 | 79.6 |  | 10,898 | 78.0 |  | 1,982,508 | 79.5 |
|  |  | 1 | 4,171 | 17.6 |  | 393,169 | 15.8 |  | 2,376 | 17.0 |  | 394,992 | 15.8 |
|  |  | 2 | 906 | 3.8 |  | 82,918 | 3.3 |  | 507 | 3.6 |  | 83,254 | 3.3 |
|  |  | 3+ | 358 | 1.5 |  | 31,440 | 1.3 |  | 192 | 1.4 |  | 31,593 | 1.3 |
| % of valid GP data to length |  | [0 - 20] | 4,345 | 18.3 |  | 589,847 | 23.8 |  | 3,161 | 22.6 |  | 591,150 | 23.7 |
| of residence in Wales*** |  | (20 - 40] | 908 | 3.8 |  | 86,174 | 3.5 |  | 552 | 4.0 |  | 86,619 | 3.5 |
|  |  | (40 - 60] | 1,615 | 6.8 |  | 151,482 | 6.1 |  | 912 | 6.5 |  | 152,035 | 6.1 |
|  |  | (60 - 80] | 2,089 | 8.8 |  | 198,688 | 8.0 |  | 1,129 | 8.1 |  | 199,748 | 8.0 |
|  |  | (80 - 100] | 14,746 | 62.2 |  | 1,456,482 | 58.7 |  | 8,219 | 58.8 |  | 1,462,795 | 58.7 |
| * measured at index dates | | | | | | | | | | | | | |
| ** measured at a period from 1 year before to index dates | | | | | | | | | | | | | |
| *** measured from available record up to index dates | | | | | | | | | | | | | |

**Supplementary Table 11**. Summary of the univariable and multivariable logistic regression for the risk of COVID-19 infection for the main and sensitivity analyses.

|  |  |  | COVID-19 Definition 1 (main analysis) | | | | | | |  | COVID-19 Definition 2 (sensitivity analysis) | | | | | | |
| --- | --- | --- | --- | --- | --- | --- | --- | --- | --- | --- | --- | --- | --- | --- | --- | --- | --- |
|  |  |  | Unadjusted | | |  | Adjusted | | |  | Unadjusted | | |  | Adjusted | | |
| Characteristics | Reference | Level | OR | 95% CI | p-value |  | OR | 95% CI | p-value |  | OR | 95% CI | p-value |  | OR | 95% CI | p-value |
| History of self-harm | No | Yes | 2.1 | (1.8 - 2.4) | <0.001 |  | 1.4 | (1.2 - 1.6) | <0.001 |  | 1.4 | (1.2 - 1.8) | <0.001 |  | 1.2 | (1.0 - 1.5) | 0.126 |
| History of mental health | No | Yes | 2.3 | (2.3 - 2.4) | <0.001 |  | 1.3 | (1.3 - 1.4) | <0.001 |  | 2.0 | (1.9 - 2.1) | <0.001 |  | 1.2 | (1.2 - 1.3) | <0.001 |
| Sex | Male | Female | 1.8 | (1.7 - 1.8) | <0.001 |  | 1.6 | (1.5 - 1.6) | <0.001 |  | 1.8 | (1.8 - 1.9) | <0.001 |  | 1.7 | (1.7 - 1.8) | <0.001 |
| Age | 10-24 yr | 25-64 yr | 2.6 | (2.5 - 2.8) | <0.001 |  | 1.8 | (1.7 - 1.9) | <0.001 |  | 2.9 | (2.7 - 3.1) | <0.001 |  | 2.5 | (2.3 - 2.7) | <0.001 |
|  |  | 65 yr + | 2.4 | (2.3 - 2.5) | <0.001 |  | 1.1 | (1.1 - 1.2) | <0.001 |  | 2.6 | (2.5 - 2.9) | <0.001 |  | 1.6 | (1.4 - 1.7) | <0.001 |
| Ethnicity | White | non-White | 1.7 | (1.6 - 1.8) | <0.001 |  | 1.8 | (1.7 - 1.9) | <0.001 |  | 2.1 | (2.0 - 2.3) | <0.001 |  | 2.2 | (2.1 - 2.4) | <0.001 |
|  |  | Unknown | 0.9 | (0.8 - 0.9) | <0.001 |  | 0.9 | (0.9 - 1.0) | <0.001 |  | 0.9 | (0.9 - 1.0) | 0.001 |  | 0.9 | (0.9 - 1.0) | <0.001 |
| WIMD quintile | Q1 | Q2 | 0.9 | (0.9 - 1.0) | <0.001 |  | 1.0 | (0.9 - 1.0) | 0.089 |  | 0.9 | (0.9 - 1.0) | <0.001 |  | 1.0 | (0.9 - 1.1) | 0.845 |
| (Q5: most deprived) |  | Q3 | 1.1 | (1.1 - 1.2) | <0.001 |  | 1.2 | (1.1 - 1.2) | <0.001 |  | 1.0 | (0.9 - 1.0) | 0.347 |  | 1.1 | (1.0 - 1.1) | 0.022 |
|  |  | Q4 | 1.2 | (1.2 - 1.3) | <0.001 |  | 1.1 | (1.1 - 1.2) | <0.001 |  | 1.1 | (1.1 - 1.2) | <0.001 |  | 1.1 | (1.0 - 1.2) | <0.001 |
|  |  | Q5 | 1.3 | (1.3 - 1.4) | <0.001 |  | 1.1 | (1.1 - 1.2) | <0.001 |  | 1.2 | (1.1 - 1.2) | <0.001 |  | 1.1 | (1.0 - 1.1) | 0.054 |
| Urban/rural indicator | Rural | Urban | 1.4 | (1.3 - 1.4) | <0.001 |  | 1.3 | (1.2 - 1.3) | <0.001 |  | 1.5 | (1.4 - 1.5) | <0.001 |  | 1.3 | (1.3 - 1.4) | <0.001 |
| Health Board | Betsi Cadwaladr | Hywel Dda | 0.5 | (0.5 - 0.5) | <0.001 |  | 0.5 | (0.5 - 0.5) | <0.001 |  | 0.5 | (0.4 - 0.5) | <0.001 |  | 0.5 | (0.5 - 0.5) | <0.001 |
|  |  | Swansea Bay | 0.9 | (0.8 - 0.9) | <0.001 |  | 0.7 | (0.7 - 0.8) | <0.001 |  | 0.8 | (0.7 - 0.8) | <0.001 |  | 0.7 | (0.7 - 0.7) | <0.001 |
|  |  | Cardiff & Vale | 1.1 | (1.1 - 1.2) | <0.001 |  | 1.0 | (0.9 - 1.0) | 0.159 |  | 1.0 | (0.9 - 1.0) | 0.298 |  | 0.8 | (0.8 - 0.9) | <0.001 |
|  |  | Cwm Taf Morgannwg | 1.0 | (1.0 - 1.1) | 0.024 |  | 0.9 | (0.9 - 0.9) | <0.001 |  | 1.0 | (1.0 - 1.1) | 0.224 |  | 1.0 | (0.9 - 1.0) | 0.131 |
|  |  | Aneurin Bevan | 0.9 | (0.9 - 1.0) | 0.006 |  | 0.9 | (0.8 - 0.9) | <0.001 |  | 0.7 | (0.6 - 0.7) | <0.001 |  | 0.6 | (0.6 - 0.6) | <0.001 |
|  |  | Powys | 1.3 | (1.2 - 1.3) | <0.001 |  | 1.7 | (1.6 - 1.8) | <0.001 |  | 0.5 | (0.4 - 0.5) | <0.001 |  | 0.5 | (0.5 - 0.6) | <0.001 |
| Live in care homes | No | Yes | 10.5 | (9.9 - 11.2) | <0.001 |  | 6.7 | (6.2 - 7.2) | <0.001 |  | 12.3 | (11.5 - 13.3) | <0.001 |  | 8.1 | (7.4 - 8.8) | <0.001 |
| In COVID-19 shielded list | No | Yes | 1.9 | (1.8 - 2.0) | <0.001 |  | 1.2 | (1.1 - 1.2) | <0.001 |  | 1.6 | (1.5 - 1.7) | <0.001 |  | 1.0 | (0.9 - 1.0) | 0.541 |
| Charlson Comorbidity Index | 0 | 1 | 1.9 | (1.8 - 1.9) | <0.001 |  | 1.5 | (1.5 - 1.6) | <0.001 |  | 1.6 | (1.5 - 1.7) | <0.001 |  | 1.5 | (1.4 - 1.5) | <0.001 |
| (unweighted) |  | 2 | 3.4 | (3.2 - 3.6) | <0.001 |  | 2.5 | (2.4 - 2.7) | <0.001 |  | 3.6 | (3.3 - 3.8) | <0.001 |  | 3.1 | (2.9 - 3.4) | <0.001 |
|  |  | 3+ | 6.7 | (6.3 - 7.1) | <0.001 |  | 4.7 | (4.3 - 5.0) | <0.001 |  | 7.5 | (6.9 - 8.1) | <0.001 |  | 6.2 | (5.7 - 6.8) | <0.001 |
| Ever smoked | No | Yes | 1.5 | (1.5 - 1.6) | <0.001 |  | 1.0 | (0.9 - 1.0) | 0.108 |  | 1.2 | (1.2 - 1.2) | <0.001 |  | 0.8 | (0.8 - 0.9) | <0.001 |
| Ever prescribed psychotropics | No | Yes | 2.1 | (2.1 - 2.2) | <0.001 |  | 1.3 | (1.2 - 1.3) | <0.001 |  | 1.6 | (1.5 - 1.6) | <0.001 |  | 1.0 | (1.0 - 1.1) | 0.490 |
| Ever prescribed opiates | No | Yes | 2.2 | (2.2 - 2.3) | <0.001 |  | 1.5 | (1.4 - 1.5) | <0.001 |  | 1.7 | (1.6 - 1.7) | <0.001 |  | 1.3 | (1.2 - 1.3) | <0.001 |
| % of length of residence | [0 - 20] | (20 - 40] | 1.0 | (0.9 - 1.0) | 0.004 |  | 1.0 | (0.9 - 1.0) | 0.007 |  | 1.0 | (1.0 - 1.0) | 0.709 |  | 1.0 | (0.9 - 1.0) | 0.605 |
| in Wales to age at index date |  | (40 - 60] | 0.8 | (0.8 - 0.8) | <0.001 |  | 0.8 | (0.8 - 0.9) | <0.001 |  | 0.8 | (0.8 - 0.9) | <0.001 |  | 0.9 | (0.8 - 0.9) | <0.001 |
|  |  | (60 - 80] | 0.7 | (0.6 - 0.7) | <0.001 |  | 0.9 | (0.8 - 0.9) | <0.001 |  | 0.7 | (0.6 - 0.7) | <0.001 |  | 0.9 | (0.8 - 0.9) | <0.001 |
|  |  | (80 - 100] | 0.5 | (0.5 - 0.5) | <0.001 |  | 0.8 | (0.7 - 0.8) | <0.001 |  | 0.5 | (0.5 - 0.5) | <0.001 |  | 0.8 | (0.7 - 0.9) | <0.001 |
| Number of moves | 0 | 1 | 1.1 | (1.1 - 1.2) | <0.001 |  | 1.1 | (1.1 - 1.1) | <0.001 |  | 1.1 | (1.0 - 1.1) | <0.001 |  | 1.1 | (1.0 - 1.1) | <0.001 |
|  |  | 2 | 1.2 | (1.1 - 1.3) | <0.001 |  | 1.1 | (1.0 - 1.2) | 0.013 |  | 1.1 | (1.0 - 1.2) | 0.025 |  | 1.1 | (1.0 - 1.2) | 0.081 |
|  |  | 3+ | 1.2 | (1.1 - 1.4) | <0.001 |  | 1.1 | (1.0 - 1.2) | 0.069 |  | 1.1 | (1.0 - 1.3) | 0.169 |  | 1.1 | (0.9 - 1.3) | 0.287 |
| % of valid GP data to length | [0 - 20] | (20 - 40] | 1.4 | (1.3 - 1.5) | <0.001 |  | 1.1 | (1.1 - 1.2) | <0.001 |  | 1.2 | (1.1 - 1.3) | <0.001 |  | 1.0 | (0.9 - 1.1) | 0.498 |
| of residence in Wales |  | (40 - 60] | 1.4 | (1.4 - 1.5) | <0.001 |  | 1.1 | (1.1 - 1.2) | <0.001 |  | 1.1 | (1.0 - 1.2) | 0.002 |  | 0.9 | (0.9 - 1.0) | 0.154 |
|  |  | (60 - 80] | 1.4 | (1.4 - 1.5) | <0.001 |  | 1.1 | (1.1 - 1.2) | <0.001 |  | 1.1 | (1.0 - 1.1) | 0.111 |  | 0.9 | (0.8 - 1.0) | 0.005 |
|  |  | (80 - 100] | 1.4 | (1.3 - 1.4) | <0.001 |  | 1.1 | (1.0 - 1.1) | <0.001 |  | 1.1 | (1.0 - 1.1) | 0.018 |  | 0.9 | (0.9 - 1.0) | <0.001 |

**Supplementary Table 12**. Observed incidence of self-harm presentations during the pre- and post-COVID follow-up periods for the main and sensitivity analyses alongside robustness checks.

|  |  | COVID-19 | Follow-up period |  |  |  |  |
| --- | --- | --- | --- | --- | --- | --- | --- |
| DiD estimation | COVID-19 Definition | Infected? (Yes/No) | (pre-/post- COVID) | Numbers | PYAR (1000 x) | Incidence | 95% CI |
| Actual | 1 | Yes | pre-COVID | 87 | 10.2 | 8.5 | (6.9 - 10.5) |
| experiment |  |  | post-COVID | 92 | 10.2 | 9.0 | (7.3 - 11.0) |
|  |  | No | pre-COVID | 5,289 | 1119.5 | 4.7 | (4.6 - 4.9) |
|  |  |  | post-COVID | 4,811 | 1119.7 | 4.3 | (4.2 - 4.4) |
|  | 2 | Yes | pre-COVID | 28 | 5.9 | 4.8 | (3.2 - 6.8) |
|  |  |  | post-COVID | 32 | 5.9 | 5.4 | (3.8 - 7.6) |
|  |  | No | pre-COVID | 5,340 | 1123.9 | 4.8 | (4.6 - 4.9) |
|  |  |  | post-COVID | 4,879 | 1124.1 | 4.3 | (4.2 - 4.5) |
| Robustness check | 1 | Yes | pre-COVID | 80 | 11.4 | 7.0 | (5.6 - 8.7) |
| (Placebo period) |  |  | post-COVID | 104 | 11.4 | 9.2 | (7.5 - 11.1) |
|  |  | No | pre-COVID | 5,019 | 1123.5 | 4.5 | (4.3 - 4.6) |
|  |  |  | post-COVID | 5,570 | 1123.4 | 5.0 | (4.8 - 5.1) |
|  | 2 | Yes | pre-COVID | 30 | 6.8 | 4.4 | (3.0 - 6.2) |
|  |  |  | post-COVID | 44 | 6.8 | 6.5 | (4.8 - 8.6) |
|  |  | No | pre-COVID | 5,120 | 1128.1 | 4.5 | (4.4 - 4.7) |
|  |  |  | post-COVID | 5,608 | 1128.0 | 5.0 | (4.8 - 5.1) |
| Robustness check | 1 | Yes | pre-COVID | 54 | 11.3 | 4.8 | (3.6 - 6.2) |
| (Randomised |  |  | post-COVID | 56 | 11.3 | 5.0 | (3.8 - 6.4) |
| COVID + |  | No | pre-COVID | 5,045 | 1123.5 | 4.5 | (4.4 - 4.6) |
| Placebo period) |  |  | post-COVID | 5,618 | 1123.4 | 5.0 | (4.9 - 5.1) |
|  | 2 | Yes | pre-COVID | 25 | 6.8 | 3.7 | (2.4 - 5.4) |
|  |  |  | post-COVID | 31 | 6.8 | 4.6 | (3.2 - 6.4) |
|  |  | No | pre-COVID | 5,125 | 1128.1 | 4.5 | (4.4 - 4.7) |
|  |  |  | post-COVID | 5,621 | 1128.0 | 5.0 | (4.9 - 5.1) |

**Supplementary Table 13**. Summary of multivariable GEE models for the risk of self-harm following COVID-19 infection using DiD approach for the main and sensitivity analyses.

|  |  |  | COVID-19 Definition 1 | | |  | COVID-19 Definition 2 | | |
| --- | --- | --- | --- | --- | --- | --- | --- | --- | --- |
|  |  |  | (main analysis) | | |  | (sensitivity analysis) | | |
| Characteristics | Reference | Level | IRR | 95% CI | p-value |  | IRR | 95% CI | p-value |
| Period | Pre | Post | 0.9 | (0.9 - 0.9) | <0.001 |  | 0.9 | (0.9 - 0.9) | <0.001 |
| COVID-19 infected | No | Yes | 1.4 | (1.1 - 1.8) | 0.001 |  | 1.0 | (0.7 - 1.5) | 0.898 |
| Period x COVID-19 infected | - | - | 1.1 | (0.8 - 1.4) | 0.729 |  | 1.2 | (0.7 - 2.0) | 0.499 |
| History of self-harm | No | Yes | 11.6 | (10.9 - 12.3) | <0.001 |  | 11.7 | (11.0 - 12.4) | <0.001 |
| History of mental health | No | Yes | 4.5 | (4.3 - 4.8) | <0.001 |  | 4.7 | (4.4 - 4.9) | <0.001 |
| Sex | Male | Female | 1.0 | (1.0 - 1.1) | 0.621 |  | 1.0 | (1.0 - 1.1) | 0.251 |
| Age | 10-24 yr | 25-64 yr | 0.3 | (0.3 - 0.3) | <0.001 |  | 0.3 | (0.3 - 0.3) | <0.001 |
|  |  | 65 yr + | 0.1 | (0.1 - 0.1) | <0.001 |  | 0.1 | (0.1 - 0.1) | <0.001 |
| Ethnicity | White | non-White | 0.8 | (0.7 - 0.9) | 0.004 |  | 0.8 | (0.7 - 0.9) | 0.006 |
|  |  | Unknown | 0.7 | (0.7 - 0.8) | <0.001 |  | 0.7 | (0.7 - 0.8) | <0.001 |
| WIMD quintile | Q1 | Q2 | 1.1 | (1.1 - 1.2) | 0.002 |  | 1.1 | (1.0 - 1.2) | 0.007 |
| (Q5: most deprived) |  | Q3 | 1.3 | (1.2 - 1.4) | <0.001 |  | 1.3 | (1.2 - 1.4) | <0.001 |
|  |  | Q4 | 1.5 | (1.4 - 1.6) | <0.001 |  | 1.4 | (1.3 - 1.6) | <0.001 |
|  |  | Q5 | 1.7 | (1.6 - 1.8) | <0.001 |  | 1.7 | (1.5 - 1.8) | <0.001 |
| Urban/rural indicator | Rural | Urban | 1.1 | (1.1 - 1.2) | <0.001 |  | 1.1 | (1.1 - 1.2) | <0.001 |
| Health Board | Betsi Cadwaladr | Hywel Dda | 1.3 | (1.2 - 1.4) | <0.001 |  | 1.3 | (1.2 - 1.4) | <0.001 |
|  |  | Swansea Bay | 1.5 | (1.4 - 1.6) | <0.001 |  | 1.5 | (1.4 - 1.6) | <0.001 |
|  |  | Cardiff & Vale | 0.8 | (0.8 - 0.9) | <0.001 |  | 0.8 | (0.8 - 0.9) | <0.001 |
|  |  | Cwm Taf Morgannwg | 1.2 | (1.1 - 1.2) | <0.001 |  | 1.2 | (1.1 - 1.2) | <0.001 |
|  |  | Aneurin Bevan | 1.2 | (1.1 - 1.3) | <0.001 |  | 1.2 | (1.1 - 1.2) | <0.001 |
|  |  | Powys | 1.4 | (1.2 - 1.6) | <0.001 |  | 1.4 | (1.2 - 1.5) | <0.001 |
| Live in care homes | No | Yes | 0.6 | (0.4 - 0.8) | 0.003 |  | 0.6 | (0.5 - 0.9) | 0.011 |
| In COVID-19 shielded list | No | Yes | 1.1 | (1.0 - 1.2) | 0.100 |  | 1.1 | (1.0 - 1.2) | 0.009 |
| Charlson Comorbidity Index | 0 | 1 | 1.0 | (1.0 - 1.1) | 0.493 |  | 1.0 | (1.0 - 1.1) | 0.282 |
| (unweighted) |  | 2 | 1.1 | (0.9 - 1.2) | 0.410 |  | 1.1 | (0.9 - 1.2) | 0.334 |
|  |  | 3+ | 1.4 | (1.1 - 1.7) | 0.001 |  | 1.4 | (1.2 - 1.7) | <0.001 |
| Ever smoked | No | Yes | 1.3 | (1.3 - 1.4) | <0.001 |  | 1.3 | (1.2 - 1.4) | <0.001 |
| Ever prescribed psychotropics | No | Yes | 2.0 | (1.9 - 2.2) | <0.001 |  | 2.0 | (1.9 - 2.1) | <0.001 |
| Ever prescribed opiates | No | Yes | 1.1 | (1.0 - 1.2) | <0.001 |  | 1.1 | (1.0 - 1.2) | <0.001 |
| % of length of residence | [0 - 20] | (20 - 40] | 0.8 | (0.7 - 0.8) | <0.001 |  | 0.7 | (0.7 - 0.8) | <0.001 |
| in Wales to age at index date |  | (40 - 60] | 0.7 | (0.6 - 0.7) | <0.001 |  | 0.7 | (0.6 - 0.7) | <0.001 |
|  |  | (60 - 80] | 0.8 | (0.8 - 0.9) | <0.001 |  | 0.9 | (0.8 - 0.9) | <0.001 |
|  |  | (80 - 100] | 0.8 | (0.7 - 0.8) | <0.001 |  | 0.7 | (0.7 - 0.8) | <0.001 |
| Number of moves | 0 | 1 | 1.2 | (1.1 - 1.3) | <0.001 |  | 1.2 | (1.1 - 1.2) | <0.001 |
|  |  | 2 | 1.4 | (1.3 - 1.5) | <0.001 |  | 1.4 | (1.3 - 1.6) | <0.001 |
|  |  | 3+ | 1.5 | (1.4 - 1.7) | <0.001 |  | 1.5 | (1.3 - 1.7) | <0.001 |
| % of valid GP data to length | [0 - 20] | (20 - 40] | 0.7 | (0.6 - 0.8) | <0.001 |  | 0.7 | (0.7 - 0.8) | <0.001 |
| of residence in Wales |  | (40 - 60] | 0.7 | (0.6 - 0.8) | <0.001 |  | 0.7 | (0.6 - 0.8) | <0.001 |
|  |  | (60 - 80] | 0.7 | (0.7 - 0.8) | <0.001 |  | 0.7 | (0.7 - 0.8) | <0.001 |
|  |  | (80 - 100] | 0.8 | (0.7 - 0.8) | <0.001 |  | 0.8 | (0.7 - 0.8) | <0.001 |

**Supplementary Table 14**. Summary of the robustness check (placebo period) for the risk of self-harm following COVID-19 infection using DiD approach for the main and sensitivity analyses.

|  |  |  | COVID-19 Definition 1 | | |  | COVID-19 Definition 2 | | |
| --- | --- | --- | --- | --- | --- | --- | --- | --- | --- |
|  |  |  | (main analysis) | | |  | (sensitivity analysis) | | |
| Characteristics | Reference | Level | IRR | 95% CI | p-value |  | IRR | 95% CI | p-value |
| Period | Pre | Post | 1.1 | (1.0 - 1.1) | <0.001 |  | 1.1 | (1.0 - 1.1) | 0.007 |
| COVID-19 infected | No | Yes | 1.3 | (1.0 - 1.6) | 0.039 |  | 1.0 | (0.7 - 1.5) | 0.822 |
| Period x COVID-19 infected | - | - | 1.2 | (0.9 - 1.6) | 0.282 |  | 1.4 | (0.9 - 2.1) | 0.172 |
| History of self-harm | No | Yes | 11.8 | (11.1 - 12.5) | <0.001 |  | 11.7 | (11.0 - 12.5) | <0.001 |
| History of mental health | No | Yes | 4.5 | (4.3 - 4.8) | <0.001 |  | 4.5 | (4.3 - 4.7) | <0.001 |
| Sex | Male | Female | 1.0 | (0.9 - 1.0) | 0.119 |  | 1.0 | (1.0 - 1.0) | 0.705 |
| Age | 10-24 yr | 25-64 yr | 0.3 | (0.3 - 0.3) | <0.001 |  | 0.3 | (0.3 - 0.3) | <0.001 |
|  |  | 65 yr + | 0.1 | (0.1 - 0.1) | <0.001 |  | 0.1 | (0.1 - 0.1) | <0.001 |
| Ethnicity | White | non-White | 0.7 | (0.7 - 0.8) | <0.001 |  | 0.7 | (0.6 - 0.8) | <0.001 |
|  |  | Unknown | 0.8 | (0.7 - 0.8) | <0.001 |  | 0.8 | (0.7 - 0.8) | <0.001 |
| WIMD quintile | Q1 | Q2 | 1.1 | (1.0 - 1.2) | 0.066 |  | 1.1 | (1.0 - 1.2) | 0.044 |
| (Q5: most deprived) |  | Q3 | 1.2 | (1.1 - 1.3) | <0.001 |  | 1.2 | (1.2 - 1.3) | <0.001 |
|  |  | Q4 | 1.5 | (1.4 - 1.6) | <0.001 |  | 1.4 | (1.3 - 1.6) | <0.001 |
|  |  | Q5 | 1.6 | (1.5 - 1.7) | <0.001 |  | 1.6 | (1.5 - 1.7) | <0.001 |
| Urban/rural indicator | Rural | Urban | 1.1 | (1.1 - 1.2) | <0.001 |  | 1.1 | (1.0 - 1.2) | <0.001 |
| Health Board | Betsi Cadwaladr | Hywel Dda | 1.2 | (1.1 - 1.3) | <0.001 |  | 1.2 | (1.2 - 1.3) | <0.001 |
|  |  | Swansea Bay | 1.5 | (1.4 - 1.6) | <0.001 |  | 1.4 | (1.3 - 1.5) | <0.001 |
|  |  | Cardiff & Vale | 0.9 | (0.8 - 1.0) | 0.021 |  | 0.9 | (0.9 - 1.0) | 0.089 |
|  |  | Cwm Taf Morgannwg | 1.1 | (1.0 - 1.2) | 0.001 |  | 1.1 | (1.0 - 1.2) | 0.002 |
|  |  | Aneurin Bevan | 1.2 | (1.1 - 1.2) | <0.001 |  | 1.1 | (1.1 - 1.2) | <0.001 |
|  |  | Powys | 1.4 | (1.3 - 1.6) | <0.001 |  | 1.4 | (1.2 - 1.6) | <0.001 |
| Live in care homes | No | Yes | 0.8 | (0.6 - 1.1) | 0.270 |  | 1.0 | (0.7 - 1.3) | 0.895 |
| In COVID-19 shielded list | No | Yes | 1.1 | (1.0 - 1.2) | 0.110 |  | 1.1 | (1.0 - 1.2) | 0.043 |
| Charlson Comorbidity Index | 0 | 1 | 1.0 | (1.0 - 1.1) | 0.132 |  | 1.1 | (1.0 - 1.2) | 0.007 |
| (unweighted) |  | 2 | 1.1 | (1.0 - 1.2) | 0.198 |  | 1.0 | (0.9 - 1.2) | 0.954 |
|  |  | 3+ | 1.2 | (1.0 - 1.5) | 0.092 |  | 1.2 | (1.0 - 1.5) | 0.125 |
| Ever smoked | No | Yes | 1.3 | (1.2 - 1.3) | <0.001 |  | 1.3 | (1.2 - 1.3) | <0.001 |
| Ever prescribed psychotropics | No | Yes | 2.0 | (1.9 - 2.1) | <0.001 |  | 2.0 | (1.9 - 2.1) | <0.001 |
| Ever prescribed opiates | No | Yes | 1.2 | (1.1 - 1.2) | <0.001 |  | 1.1 | (1.1 - 1.2) | <0.001 |
| % of length of residence | [0 - 20] | (20 - 40] | 0.7 | (0.6 - 0.7) | <0.001 |  | 0.7 | (0.7 - 0.7) | <0.001 |
| in Wales to age at index date |  | (40 - 60] | 0.6 | (0.6 - 0.6) | <0.001 |  | 0.6 | (0.6 - 0.6) | <0.001 |
|  |  | (60 - 80] | 0.8 | (0.8 - 0.9) | <0.001 |  | 0.8 | (0.8 - 0.9) | <0.001 |
|  |  | (80 - 100] | 0.7 | (0.7 - 0.8) | <0.001 |  | 0.7 | (0.7 - 0.8) | <0.001 |
| Number of moves | 0 | 1 | 1.1 | (1.1 - 1.2) | <0.001 |  | 1.1 | (1.1 - 1.2) | <0.001 |
|  |  | 2 | 1.2 | (1.1 - 1.3) | <0.001 |  | 1.3 | (1.1 - 1.4) | <0.001 |
|  |  | 3+ | 1.3 | (1.1 - 1.6) | <0.001 |  | 1.3 | (1.1 - 1.6) | <0.001 |
| % of valid GP data to length | [0 - 20] | (20 - 40] | 0.7 | (0.6 - 0.8) | <0.001 |  | 0.8 | (0.7 - 0.9) | <0.001 |
| of residence in Wales |  | (40 - 60] | 0.7 | (0.7 - 0.8) | <0.001 |  | 0.7 | (0.7 - 0.8) | <0.001 |
|  |  | (60 - 80] | 0.7 | (0.6 - 0.7) | <0.001 |  | 0.7 | (0.6 - 0.7) | <0.001 |
|  |  | (80 - 100] | 0.8 | (0.7 - 0.8) | <0.001 |  | 0.8 | (0.7 - 0.8) | <0.001 |

**Supplementary Table 15**. Summary of the robustness check (randomised COVID + placebo period) for the risk of self-harm following COVID-19 infection using DiD approach for the main and sensitivity analyses.

|  |  |  | COVID-19 Definition 1 | | |  | COVID-19 Definition 2 | | |
| --- | --- | --- | --- | --- | --- | --- | --- | --- | --- |
|  |  |  | (main analysis) | | |  | (sensitivity analysis) | | |
| Characteristics | Reference | Level | IRR | 95% CI | p-value |  | IRR | 95% CI | p-value |
| Period | Pre | Post | 1.1 | (1.0 - 1.1) | <0.001 |  | 1.1 | (1.0 - 1.1) | 0.006 |
| COVID-19 infected | No | Yes | 1.1 | (0.8 - 1.4) | 0.571 |  | 0.8 | (0.5 - 1.2) | 0.221 |
| Period x COVID-19infected | - | - | 0.9 | (0.6 - 1.3) | 0.477 |  | 1.1 | (0.7 - 1.9) | 0.653 |
| History of self-harm | No | Yes | 11.8 | (11.1 - 12.5) | <0.001 |  | 11.7 | (11.0 - 12.5) | <0.001 |
| History of mental health | No | Yes | 4.5 | (4.3 - 4.8) | <0.001 |  | 4.5 | (4.3 - 4.7) | <0.001 |
| Sex | Male | Female | 1.0 | (0.9 - 1.0) | 0.144 |  | 1.0 | (1.0 - 1.0) | 0.679 |
| Age | 10-24 yr | 25-64 yr | 0.3 | (0.3 - 0.3) | <0.001 |  | 0.3 | (0.3 - 0.3) | <0.001 |
|  |  | 65 yr + | 0.1 | (0.1 - 0.1) | <0.001 |  | 0.1 | (0.1 - 0.1) | <0.001 |
| Ethnicity | White | non-White | 0.7 | (0.7 - 0.8) | <0.001 |  | 0.7 | (0.6 - 0.8) | <0.001 |
|  |  | Unknown | 0.8 | (0.7 - 0.8) | <0.001 |  | 0.8 | (0.7 - 0.8) | <0.001 |
| WIMD quintile | Q1 | Q2 | 1.1 | (1.0 - 1.2) | 0.065 |  | 1.1 | (1.0 - 1.2) | 0.044 |
| (Q5: most deprived) |  | Q3 | 1.2 | (1.1 - 1.3) | <0.001 |  | 1.2 | (1.2 - 1.3) | <0.001 |
|  |  | Q4 | 1.5 | (1.4 - 1.6) | <0.001 |  | 1.4 | (1.3 - 1.6) | <0.001 |
|  |  | Q5 | 1.6 | (1.5 - 1.7) | <0.001 |  | 1.6 | (1.5 - 1.7) | <0.001 |
| Urban/rural indicator | Rural | Urban | 1.1 | (1.1 - 1.2) | <0.001 |  | 1.1 | (1.0 - 1.2) | <0.001 |
| Health Board | Betsi Cadwaladr | Hywel Dda | 1.2 | (1.1 - 1.3) | <0.001 |  | 1.2 | (1.2 - 1.3) | <0.001 |
|  |  | Swansea Bay | 1.5 | (1.4 - 1.6) | <0.001 |  | 1.4 | (1.3 - 1.5) | <0.001 |
|  |  | Cardiff & Vale | 0.9 | (0.8 - 1.0) | 0.022 |  | 0.9 | (0.9 - 1.0) | 0.089 |
|  |  | Cwm Taf Morgannwg | 1.1 | (1.0 - 1.2) | 0.001 |  | 1.1 | (1.0 - 1.2) | 0.002 |
|  |  | Aneurin Bevan | 1.2 | (1.1 - 1.2) | <0.001 |  | 1.1 | (1.1 - 1.2) | <0.001 |
|  |  | Powys | 1.5 | (1.3 - 1.6) | <0.001 |  | 1.4 | (1.2 - 1.6) | <0.001 |
| Live in care homes | No | Yes | 0.9 | (0.6 - 1.2) | 0.328 |  | 1.0 | (0.7 - 1.3) | 0.942 |
| In COVID-19 shielded list | No | Yes | 1.1 | (1.0 - 1.2) | 0.099 |  | 1.1 | (1.0 - 1.2) | 0.042 |
| Charlson Comorbidity Index | 0 | 1 | 1.1 | (1.0 - 1.1) | 0.119 |  | 1.1 | (1.0 - 1.2) | 0.007 |
| (unweighted) |  | 2 | 1.1 | (1.0 - 1.3) | 0.176 |  | 1.0 | (0.9 - 1.2) | 0.937 |
|  |  | 3+ | 1.2 | (1.0 - 1.5) | 0.075 |  | 1.2 | (1.0 - 1.5) | 0.113 |
| Ever smoked | No | Yes | 1.3 | (1.2 - 1.3) | <0.001 |  | 1.3 | (1.2 - 1.3) | <0.001 |
| Ever prescribed psychotropics | No | Yes | 2.0 | (1.9 - 2.1) | <0.001 |  | 2.0 | (1.9 - 2.1) | <0.001 |
| Ever prescribed opiates | No | Yes | 1.2 | (1.1 - 1.2) | <0.001 |  | 1.1 | (1.1 - 1.2) | <0.001 |
| % of length of residence | [0 - 20] | (20 - 40] | 0.7 | (0.6 - 0.7) | <0.001 |  | 0.7 | (0.7 - 0.7) | <0.001 |
| in Wales to age at index date |  | (40 - 60] | 0.6 | (0.6 - 0.6) | <0.001 |  | 0.6 | (0.6 - 0.6) | <0.001 |
|  |  | (60 - 80] | 0.8 | (0.8 - 0.9) | <0.001 |  | 0.8 | (0.8 - 0.9) | <0.001 |
|  |  | (80 - 100] | 0.7 | (0.7 - 0.8) | <0.001 |  | 0.7 | (0.7 - 0.8) | <0.001 |
| Number of moves | 0 | 1 | 1.1 | (1.1 - 1.2) | <0.001 |  | 1.1 | (1.1 - 1.2) | <0.001 |
|  |  | 2 | 1.2 | (1.1 - 1.3) | <0.001 |  | 1.3 | (1.1 - 1.4) | <0.001 |
|  |  | 3+ | 1.3 | (1.1 - 1.6) | <0.001 |  | 1.3 | (1.1 - 1.6) | <0.001 |
| % of valid GP data to length | [0 - 20] | (20 - 40] | 0.7 | (0.6 - 0.8) | <0.001 |  | 0.8 | (0.7 - 0.9) | <0.001 |
| of residence in Wales |  | (40 - 60] | 0.7 | (0.7 - 0.8) | <0.001 |  | 0.7 | (0.7 - 0.8) | <0.001 |
|  |  | (60 - 80] | 0.7 | (0.6 - 0.7) | <0.001 |  | 0.7 | (0.6 - 0.7) | <0.001 |
|  |  | (80 - 100] | 0.8 | (0.7 - 0.8) | <0.001 |  | 0.8 | (0.7 - 0.8) | <0.001 |
