## Supplementary Methods for "Healthcare presentations with self-harm and the association with COVID-19: an e-cohort whole-population-based study using individual-level linked routine electronic health records in Wales, UK, 2016 - March 2021"

***Identification of COVID-19 infection***

COVID-19 infection was defined by ICD-10 code U07.3 or positive result from antibody test. Active and confirmed infection was defined by positive result from laboratory PCR test, or by ICD-10 code U07.1 or Read codes Y20d1, Y20ce, 4J3R1, A795. A7951 or 65PW1. Active and suspected infection was defined by ICD-10 code U07.2 or Read codes Y20cf or 1JX1..

In keeping with others (Lusignan & Williams, 2020; NHS Digital, 2020; Taquet et al., 2020) and considering that use of COVID-19 specific codes was not widespread during the early stage of the pandemic, we additionally included ICD-10 codes (B34.2, B97.2, J12.8 and U04.9) and Read codes (65PW. and 1JX..), originally used for general coronavirus infections (e.g., pneumonia due to Severe Acute Respiratory Syndrome), as active and suspected COVID-19 infection given the diagnoses made after 28/02/2020.

***Covariates included in the bidirectional associations between self-harm and COVID-19 infection***

For the association of COVID-19 infection following self-harm, all covariates were time-fixed and measured at the defined index date (see Methods for its definition). The same set of covariates were used for assessing the association of self-harm following COVID-19 infection. However, a longitudinal approach was adopted, and counterfactual index dates were defined as two years before the actual index dates (Suppl. Figure 9A). This allowed for the use of time-varying covariates corresponded to the actual and counterfactual index dates. We summarise as follows the covariates used (for COVID-19 infection, please refer to Methods) for both associations and the choice of time-fixed or time-varying covariate for the modelling of the risk of self-harm following COVID-19 infection:

| Variable | Time-fixed/  varying?* | Categories | Time of measurement for time-varying covariates* |
| --- | --- | --- | --- |
| Sex | Time-fixed | Male, Female | - |
| Age | Time-varying | 10-24, 25-64, 65+ | At index dates |
| Ethnicity | Time-fixed | White, Black, Asian, Mixed and others^1^ | - |
| Area deprivation (WIMD quintile) | Time-varying | Q1 to Q5 (see Methods) | At index dates |
| Urban/Rural indicator | Time-varying | Urban/Rural | At index dates |
| Health Board | Time-varying | Seven health boards in Wales | At index dates |
| Live in care homes^2^ | Time-varying | Yes, No | At index dates |
| In COVID-19 shielded list^3^ | Time-fixed | Yes, No | - |
| Unweighted Charlson Comorbidity Index^4^ | Time-varying | 0, 1, 2, 3+ | Between 1 year before index dates and index dates (see Supplementary Fig 13A for details) |
| Ever Smoked^5^ | Time-varying | Yes, No | Ever up to index dates |
| History of self-harm | Time-varying | Yes, No | Ever up to index dates |
| History of mental health^6^ | Time-varying | Yes, No | Ever up to index dates |
| Prescription of Psychotropic medications^7^ | Time-varying | Yes, No | Ever up to index dates |
| Prescription of opiates medications^7^ | Time-varying | Yes, No | Ever up to index dates |
| % of length of residence in Wales w. r. t. age | Time-varying | [0-20], (20-40], (40-60], (60-80], (80-100] | Ever up to index dates |
| Number of moves in residential address | Time-varying | 0, 1, 2, 3+ | Ever up to index dates |
| % of duration of valid GP data w. r. t. Wales residence^8^ | Time-varying | [0-20], (20-40], (40-60], (60-80], (80-100] | Ever up to index dates |

* Time-varying covariates only apply to the longitudinal modelling of the risk of self-harm following COVID-19 infection where changes in

categories of those covariates are allowed from the counterfactual to actual index dates (Suppl. Figure 9A).

^1^ Further collapse to White and non-White for modelling to ensure enough sample size.

^2^ Refer to Hollinghurst et al. (2021) for the detailed methodological descriptions.

^3^ Refer to <https://nwis.nhs.wales/news/latest-news/identifying-vulnerable-patient-lists/> and <https://nwis.nhs.wales/coronavirus/digital-support-updates-for-healthcare-professionals/identifying-shielding-patients/> for the identification of vulnerable patients.

^4^ Refer to Charlson et al. (1987) for its definition and Khan et al. (2010) and for the list of codes used.

^5^ Refer to Atkinson et al. (2017) for the detailed methodological descriptions.

^6^ Refer to John et al. (2020) for list of codes used.

^7^ Available from the WLGP only. Refer to John et al. (2020) for list of codes used.

^8^ Refer to Davies et al. (2018) and Thayer et al. (2020) for the description of defining duration of valid GP data.

***Statistical Analysis – Incidence and prevalence of self-harm contacts***

For each week, we measured incidence and prevalence of self-harm contacts. Incidence contact was defined as the first contact in a 12-month period with data available. During incidence and prevalence computation, only periods of known Welsh residency were considered. We measured ratio of rate ratios (RRRs) (and 95% confidence intervals, CIs) of 2020-March 2021 compared to each counterfactual period 2016-2019. Bonferroni adjustment was used to correct for multiple comparisons. Results can be found in Suppl. Figures 11-13.

***Statistical Analysis – modelling for weekly time trends and contrast of model coefficients using Difference-in-difference (DiD) approach***

We modelled weekly time trends via generalised estimating equations (GEE) using robust variance for parameter estimation (Agresti, 2007). We adopted the binomial distribution with logit link function for proportion outcomes and Poisson distribution with log link function for prevalence and incidence outcomes. The exchangeable within-subject correlation structure was chosen to circumvent correlation of outcomes over time based on the quasilikelihood under the independence model criterion as described previously (Cui & Qian, 2007; Pan, 2002). In all models, time was the key independent variable and as an indicator variable to estimate variation of outcomes each week within our observation period. We only included the weeks from the beginning of each year to the week corresponding to the end of the observation period in the modelling. Sex and age groups were represented by categorical variables and by WIMD quintile (1-5) as ranked categories. Only for modelling, we re-grouped age group as a two-level categorical variable (10-24 vs. >24 years) to circumvent a non-convergence issue due to small sample size in age group >64 years. We included three time-related 2^nd^ order interaction terms in four separate models, namely, time-by-sex, time-by-age and time-by-WIMD to disentangle overall and time-varying differential effects of age, sex and area deprivation on self-harm outcomes.

We reported ratios (and 95% confidence intervals, CIs) of the odds/rate ratios of 2020/2021 compared to each counterfactual period 2016-2018/2019, i.e., ratio of odds ratios (RORs) for proportion and ratios of rate ratios (RRRs) for prevalence and incidence. RRRs/RORs different from one reflect a difference in 2020/2021 trend compared to the previous four years. Whether RRRs/RORs >1 or <1 depends on outcome trends. Arithmetic means of the model coefficients were used for periods spanning more than one week (e.g., week 1 to 10). Comparisons across age groups, sex and deprivation level were also performed (i.e., triple differences) by examining whether the ratios of RRRs and RORs between subgroups are significantly different from unity. All ratios were compiled using ‘contrast’ command in Stata after modelling and Bonferroni adjustment was used to correct for multiple comparisons. We repeated these analyses stratifying by age, sex and WIMD quintile separately.

***Statistical Analysis – DiD estimator for the risk of self-harm following COVID-19 infection****.*

We adopted longitudinal two-groups-two-periods DiD approach (Wing et al., 2018) to assess the risk of self-harm following COVID-19 infection. The model specification is as follows:

$$logY-log\left( PYAR \right)=\beta_{0}+\beta_{1}COVID+\beta_{2}Period+\beta_{3}\left( COVID\times Period \right)+X^{T}\beta+\epsilon$$

Where *Y* = 0, 1 as the indicator of having self-harm contact; *PYAR* = person-years-at-risk as offset; *COVID* = 0, 1 as the COVID-19 infection indicator, *Period* = 1, 2 as the indicator for pre- and post-COVID period respectively; row vector *X* = list of the mentioned covariates; *β*_0_, *β*_1_, *β*_2_, *β*_3_ and row vector *β* = model coefficients, and *ϵ* = error term.

We ran GEE using exchangeable within-subject working correlation matrix and robust variance for parameter estimation. Poisson distribution with log link function was adopted for incidence. The parameter of interest, i.e., the DiD estimator, is the exponentiation of *β*_3_ and the corresponding confidence intervals.

Interpretation of the DiD estimator was the same as in the RRR. We performed a sensitivity analysis by repeating all modelling, but definition of infection was limited to those having any active confirmed infections. For the DiD estimation, we conducted two robustness checks against the common trend assumption as previously proposed (Lechner, 2011; Wing et al., 2018). The first was termed “placebo period” check: repeating the DiD estimation by shifting the index dates and follow-ups one year before, presuming COVID-19 outbreak occurred in the previous year (2019). The second was termed “randomised COVID + placebo period” check: repeating the first “placebo period” check but additionally randomising the COVID-19 infection group conditional on the actual infection proportion. The common trend assumption was deemed appropriate if DiD estimators from both checks approach unity.

**Sensitivity analyses**

To ascertain the effect data coverage may have had in our results, we replicated the main analysis (i.e., without stratification) using for each week the sub-population registered with a GP providing data to SAIL on that Monday. These results were similar to and yielded the same conclusions than the main results presented in the text.

For self-harm and COVID-19 infection, the main analyses were replicated (results shown in Suppl. Figure 10 and Suppl. Table 11-15) using more stringent COVID-19 infection ascertainment criteria. The results showed that history of self-harm was a significant risks factor of infection in the unadjusted but not adjusted model while sex, age, ethnicity, deprivation and living in care homes were still statistically robust in both models (Suppl, Table 11). Patterns of incidences of self-harm between pre-COVID and post-COVID periods and between infected and not infected groups, together with the DiD estimator were similar to the main analysis (Figure 4C, Suppl. Figure 10 and Suppl. Table 12-13).

*Robustness checks on DiD estimators (self-harm sequalae of COVID-19 infection)*

In the “placebo period” robustness check (pretending COVID-19 occurred in 2019 in Wales), incidence of self-harm did not decrease but slightly increase between pre-COVID and post-COVID periods in the not infected group (Suppl. Figure 10, Suppl. Table 12). However, increase in incidence was still larger in the infected group, resulting in the DiD estimator > 1 although not reaching level significance (Figure 4C, Suppl. Table 14). This between group difference was no longer discernible (DiD estimator approaching 1) by first randomising the COVID-19 infection group and repeating the “placebo period” check (Figure 4C, Suppl. Figure 10 and Suppl. Table 15).

**References for Supplementary Methods**

Agresti A. *An Introduction to Categorical Data Analysis*. 2nd ed. Hoboken, NJ: John Wiley & Sons Inc; 2007.

Atkinson, M. D., Kennedy, J. I., John, A., Lewis, K. E., Lyons, R. A., & Brophy, S. T. (2017). Development of an algorithm for determining smoking status and behaviour over the life course from UK electronic primary care records. *BMC medical informatics and decision making, 17*(1), 1-12.

Charlson, M. E., Pompei, P., Ales, K. L., & MacKenzie, C. R. (1987). A new method of classifying prognostic comorbidity in longitudinal studies: development and validation. *Journal of Chronic Diseases, 40*(5), 373–83. https://doi.org/10.1016/0021-9681(87)90171-8

Cui, J., & Qian, G. (2007). Selection of working correlation structure and best model in GEE analyses of longitudinal data. *Communications in statistics—Simulation and computation*, *36*(5), 987-996.

Davies, G., et al. (2018). Long term extension of a randomised controlled trial of probiotics using electronic health records. *Scientific reports, 8*(1), 1-8.

Hollinghurst, J. et al. (2021). The impact of COVID-19 on adjusted mortality risk in care homes for older adults in Wales, UK: a retrospective population-based cohort study for mortality in 2016–2020. *Age and ageing, 50*(1), 25-31

John, A., et al. (2020). Contacts with primary and secondary healthcare prior to suicide: case–control whole-population-based study using person-level linked routine data in Wales, UK, 2000–2017. *The British Journal of Psychiatry, 217*(6), 717-724.

Khan, N. F., Perera, R., Harper, S., & Rose, P. W. (2010). Adaptation and validation of the Charlson Index for Read/OXMIS coded databases. *BMC Family Practice, 11*, 1. <https://doi.org/10.1186/1471-2296-11-1>

Lechner M. The estimation of causal effects by difference-in-difference methods. Foundations and Trends in Econometrics.2011 Nov; 4(3):165-224.

Maxime Taquet, Sierra Luciano, John R Geddes, Paul J Harrison, Bidirectional associations between COVID-19 and psychiatric disorder: retrospective cohort studies of 62 354 COVID-19 cases in the USA, The Lancet Psychiatry, 2020, ISSN 2215-0366, https://doi.org/10.1016/S2215-0366(20)30462-4

NHS Digital, 2020, https://gov.wales/antibody-testing-coronavirus-covid-19.

Pan, W. (2002). Goodness‐of‐fit tests for GEE with correlated binary data. *Scandinavian Journal of Statistics*, *29*(1), 101-110.

Simon de Lusignan and John Williams, To monitor the COVID-19 pandemic we need better quality primary care data, BJGP Open 2020; 4 (2): bjgpopen20X101070. DOI: https://doi.org/10.3399/bjgpopen20X101070

Pan, W. (2002). Goodness‐of‐fit tests for GEE with correlated binary data. *Scandinavian Journal of Statistics*, *29*(1), 101-110.

Thayer, D. et al. (2020). Measuring follow-up time in routinely-collected health datasets: challenges and solutions. Plos one, 15(2), e0228545.

Wing, C., Simon, K., & Bello-Gomez, R. A. (2018). Designing difference in difference studies: best practices for public health policy research. *Annual review of public health, 39*.
